# Genetic landscape of rare autoinflammatory disease variants in Qatar and Middle Eastern populations through the integration of whole-genome and exome datasets

**DOI:** 10.1101/2020.08.10.20171363

**Authors:** Parul Sharma, Abhinav Jain, Vinod Scaria

## Abstract

Rare monogenic autoinflammatory diseases are a group of recurrent inflammatory genetic disorders caused due to genetic variants in over 37 genes. While a number of these disorders have been identified and reported from the Middle Eastern populations, the carrier frequency of these genetic variants in the Middle Eastern populations is not known. The availability of whole-genome and exome datasets of over a thousand individuals from Qatar persuaded us to explore the genetic epidemiology of rare autoinflammatory genetic variants. We have systematically analyzed genetic variants in genome-scale datasets from Qatar with a compendium of variants associated with autoinflammatory diseases. The variants were systematically reclassified according to the American College of Medical Genetics and Genomics guidelines for interpretation of variant pathogenicity. Our analysis identified 7 pathogenic and likely pathogenic variants with significant differences in their allele frequencies compared to the global population. The cumulative carrier frequency of these variants was found to be 2.58%. Furthermore, our analysis revealed that 5 genes implicated in rare autoinflammatory diseases were under natural selection. To our best knowledge, this is the first and comprehensive study on the population-scale analysis and genetic epidemiology for genetic variants causing rare autoinflammatory disease in Middle Eastern populations.

## Introduction

Autoinflammatory disorders are genetically heterogeneous groups of disorders caused due to the presence of disease-causing variants in the genes responsible for regulating the inflammatory response. However, monogenic autoinflammatory disorders are the group of Mendelian genetic disorders characterized by recurrent inflammatory episodes due to an abnormal innate immune system. Until recently, these disorders were only defined by phenotypic traits including recurrent attacks of fever, abdominal pain, arthritis, skin rashes or cutaneous signs and were known to exhibit overlapping symptoms with other diseases which leads to misdiagnosis in a large number of cases (Ben-Chetrit et al., 2018; Hausmann et al., 2018). Recent advancements in the understanding of the molecular basis of these disorders have resulted in an accurate classification that presently encompasses 20 disorders involving over 30 genes (Touitou et al., 2004; Ciccarelli et al., 2014; Martinez-Quiles y Goldbach-Mansky, 2018). The most common among all monogenic autoinflammatory diseases is Familial Mediterranean Fever (FMF). It is caused by the pathogenic variants in the *MEFV* gene and is highly prevalent in the Middle Eastern countries (The International FMF Consortium, 1997; Salehzadeh, 2015). A recent study from our group has estimated the population-specific frequencies of *MEFV* variants associated with FMF among 2000 Mediterranean individuals (Koshy et al., 2018).

There are multiple reports of autoinflammatory disorders from Middle Eastern countries including reports from a single hospital in Riyadh where thirty-four patients were admitted due to autoinflammatory disorders in 10 years (Alenazi et al., 2012). Also, there are multiple individuals from Arab countries affected by Majeed syndrome (Majeed et al., 1989; Al-Mosawi et al., 2007; Alenazi et al., 2012). Aladbe and group performed a retrospective study in Qatar for 5 years on the clinical and genetic profiling of ∼70 symptomatic and carrier individuals affected with autoinflammatory disorder and reported the expansion of the autoinflammatory disorder in Qatar (Aladbe et al., 2013). Also, in an web-based international registry of the autoinflammatory disorders i.e. Eurofever, composed of baseline and clinical information of 1,880 patients affected with autoinflammatory disease from 31 countries found that the second largest number of patients belonged to the eastern and southern Mediterranean region (Toplak et al., 2012). Incidence reports from Sweden on autoinflammatory disorders and hereditary amyloidosis found to be 98% from the Eastern Mediterranean region origin, mainly young Syrian descendants (Hemminki et al., 2013). Due to high consanguineous marriages in Arab countries, it provides an opportunity to find novel risk loci like *LACC1* and *LRBA* that were found to be associated with the monogenic juvenile idiopathic arthritis (JIA) and inflammatory bowel disease with combined immunodeficiency respectively in Arab population (Alangari et al., 2012; Patel et al., 2014; Martorana et al., 2017).

The recent availability of sequencing data from Middle East populations that were not previously covered in global sequencing projects motivates us to understand the genetic epidemiology of monogenic autoinflammatory diseases (Fakhro et al., 2016; Scott et al., 2016). In the present analysis, we have performed integrative analysis with extensive literature screening for classifying variants in autoinflammatory disorders. We have further attempted to understand the prevalence of these disorders by studying the allele and genotype frequencies of the variants in the Qatar population. Our analysis points to significant differences in the allele frequencies even for the small population studied.

## Materials and Methods

### Population-scale datasets of genetic variants

The analysis was performed on population-scale datasets which involve 1005 Qatar Genomes and Exomes (Fakhro et al., 2016) and 1,111 Greater Middle East exomes (Scott et al., 2016).

### Genomes and Exomes from Qatar

The Qatar dataset comprises a total of 88 whole genome and 917 whole exome sequences and a total of 20,937,965 single nucleotides variants and indels that were identified in alignment to the hg19/GRCh37 version of reference human genome. The sequenced individuals belong to seven different sub-populations within Qatar namely: European (EUR), South Asian (SA), Bedouin (BED), African Pygmy (AP), Arab (AB), Persian (PER), and Sub Saharan African (SAF) (Fakhro et al., 2016).

### Greater Middle East (GME) Variome Project

The GME Variome Project comprised of high quality whole exome data of 1,111 unrelated individuals from six different regions of GME which include Northwest Africa (NWA, n=85), Northeast Africa (NEA, n=423), Turkish Peninsula (TP, n=140), Syrian Desert (SD, n=81), Arabian Peninsula (AP, n=214), and Persia and Pakistan (PP, n=168). The dataset consists of a total of 689,297 SNVs and indels which were detected in alignment to the hg19/GRCh37 human reference genome (Scott et al., 2016).

### Datasets of disease-associated genetic variants

Three independent datasets of genetic variants were compiled for the analysis. This includes ClinVar, Infevers and the Human Gene Mutation Database (HGMD).

The ClinVar database [ClinVar version: 2018-02-25] variants which were annotated as pathogenic or likely pathogenic were retrieved (Landrum et al., 2018). These variants were further filtered based on a list of 37 rare autoinflammatory genes and then matched with the Qatar variants. The autoinflammatory genes and its associated disorders are tabulated in **Supplementary Table 1**.

**Table 1.**
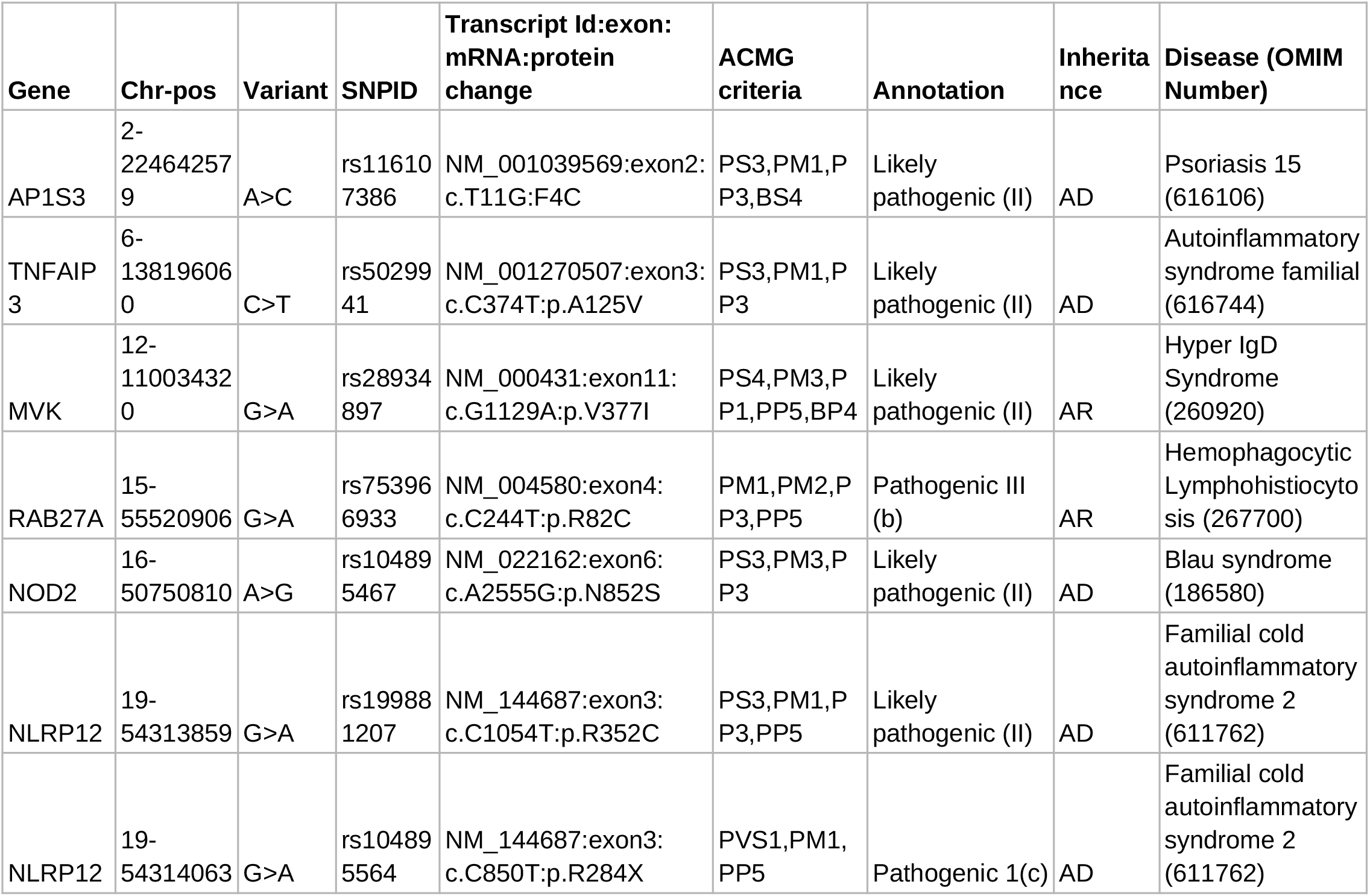
Pathogenic and Likely Pathogenic variants annotated as per ACMG guidelines for interpretation of variants.

Infevers (accessed on 2018-28-02) is a publicly available database of genetic variants implicated in autoinflammatory disorders. The database lists a total of 1,586 genetic variants for 30 genes associated with 31 autoinflammatory disorders (Milhavet et al., 2008) (Infevers). The rare autoinflammatory variants for 29 genes excluding *MEFV* as it has already been mentioned (Milhavet et al., 2008; Koshy et al., 2018) were downloaded and compiled. These variants were further filtered for the variants identified in the Qatar dataset.

The Human Gene Mutation Database (HGMD public version 2018) is a comprehensive resource and a collection of genetic variants in human genes (Milhavet et al., 2008; Stenson et al., 2014; Koshy et al., 2018). The data for all 37 autoinflammatory genes was manually compiled from the public version of the HGMD and filtered for the variants identified in the Qatar dataset.

### Computational Annotation protocol for variants

All the autoinflammatory variants from ClinVar, HGMD, and Infevers that were detected in the Qatar datasets were further annotated by using software ANNOVAR (v. date 2017-07-17) for the functional annotation which is composed of annotations from various databases (Wang et al., 2010). These databases include RefGene, dbnsfp33a, dbscsnv11, avsnp147, intervar_20170202, kaviar_20150923 and GWAS catalog for prediction of variants’ pathogenicity and other characteristics. There are databases for allele frequency across the global population which consist of esp6500si_all, exac03, and 1000g2015aug_all. We also annotated our variants using ClinVar (v. date 2018-02-25) which comprises human variations associated with a phenotype (Landrum et al., 2018).

#### Annotation of genetic variants according to ACMG and AMP guidelines

The expert panel from American College of Medical Genetics and Genomics (ACMG) and Association for Molecular Pathology (AMP) has put forward 28 criteria for variant classification into 5 broad categories i.e. pathogenic, likely pathogenic, benign, likely benign and VUS. Each criterion depending on the pathogenicity of the variant was weighted as very strong (PVS1), strong (PS1-4), moderate (PM1-6) or supporting (PP1-5). A similar case for benign characteristics was weighted as stand-alone (BA1), strong (BS1-4) or supporting (BP1-7). These criteria in combination classify the variant into the 5 broad categories (Richards et al., 2015). The detailed protocol for annotation of variants and assigning weighted criteria is described in Supplementary data 1.

Variants are assigned different criteria depending on the various evidences from literature and other databases. These criteria were put together in the Genetic Variant Interpretation tool (Genetic Variant Interpretation Tool) which classifies them into 5 categories viz pathogenic, likely pathogenic, benign, likely benign and VUS.

### Global Population Datasets

We employed four large datasets of global populations to compare allele frequencies that include:

The 1000 Genomes project (1000g2015aug_all) dataset encompasses genomes of 2504 individuals from 5 major populations i.e. Europe (EUR), South Asia (SAS), Africa (AFR), East Asia (EAS), and the America (AMR) (The 1000 Genomes Project Consortium, 2015).

The National Heart Lung and Blood Institute (NHLBI) Exome Sequencing Project (ESP) comprises unrelated 2203 African-American and 4300 European-American totaling to 6503 individuals’ exomes sequenced (Fu et al., 2013),

The Genome Aggregation Database (gnomAD V2) comprises data from 123,136 exomes and 15,496 whole genomes of unrelated individuals. These individuals belong to different ancestries i.e. African/African American, Latino, Ashkenazi Jewish, East Asian, Finnish, Non-Finish European, South Asian and other (Karczewski et al., 2019). We have used a combined allele frequency of the gnomAD V2 exomes and genomes dataset. There might be a possibility that the variants are genotyped in the genomes but absent in the exomes or vice-versa.

The updated version of the gnomAD V3.1 comprises genetic data from 76,156 whole genome sequences of unrelated individuals with >759 million short nuclear variants. These individuals belong to nine ancestral groups namely African/African-American, Amish, Latino/Admixed American, Ashkenazi Jewish, East Asian, Finnish, Non-Finnish European, Middle Eastern, South Asian and other (Karczewski et al., 2019).

### Statistical Significance of pathogenic variants

The differences in allele frequencies of variants, annotated as pathogenic or likely pathogenic by ACMG guidelines, were tested for statistically significant differences with the global population. We have compared the allele frequencies of the Qatar and GME populations and their subpopulation to the gnomAD all V2. We used gnomAD v2, since gnomAD V2 encompasses all the variants that are annotated as pathogenic or likely pathogenic in Qatar dataset. For statistical significance, we implemented Fisher’s exact test and data with a *p-*value less than 0.05 was considered significant.

### Natural Selection of autoinflammatory gene

Natural Selection of genes was predicted based on differences in the derived allele from the ancestral allele which was predicted using Integrated Haplotype Score (iHS) or by differences in allele frequency between populations to deduce selection pressure i.e. Wright’s fixation index (Fst) (Vitti et al., 2013; Karczewski et al., 2019). iHS for variants in the gene should be >2 or <-2 to be predicted as naturally selected (MerrimanLab / selectionTools). So we calculated iHS for all variants of the Qatar population using the selection tools pipeline (Cadzow et al., 2014) which is pubically available on GitHub. Out of which the top 1% of variants were filtered. Those autoinflammatory genes which fall in the top 1% criteria of iHS have been taken up for further analysis and their Fst scores were calculated using PLINK algorithm to infer selection pressure among populations (Purcell et al., 2007; Cadzow et al., 2014). We calculated the Fst score among populations i.e. Qatar population and its subpopulations with 1000 genome subpopulations (African, European and South Asian). The methodology adopted in this study is represented in **Supplementary Figure 1**.

**Figure 1:**
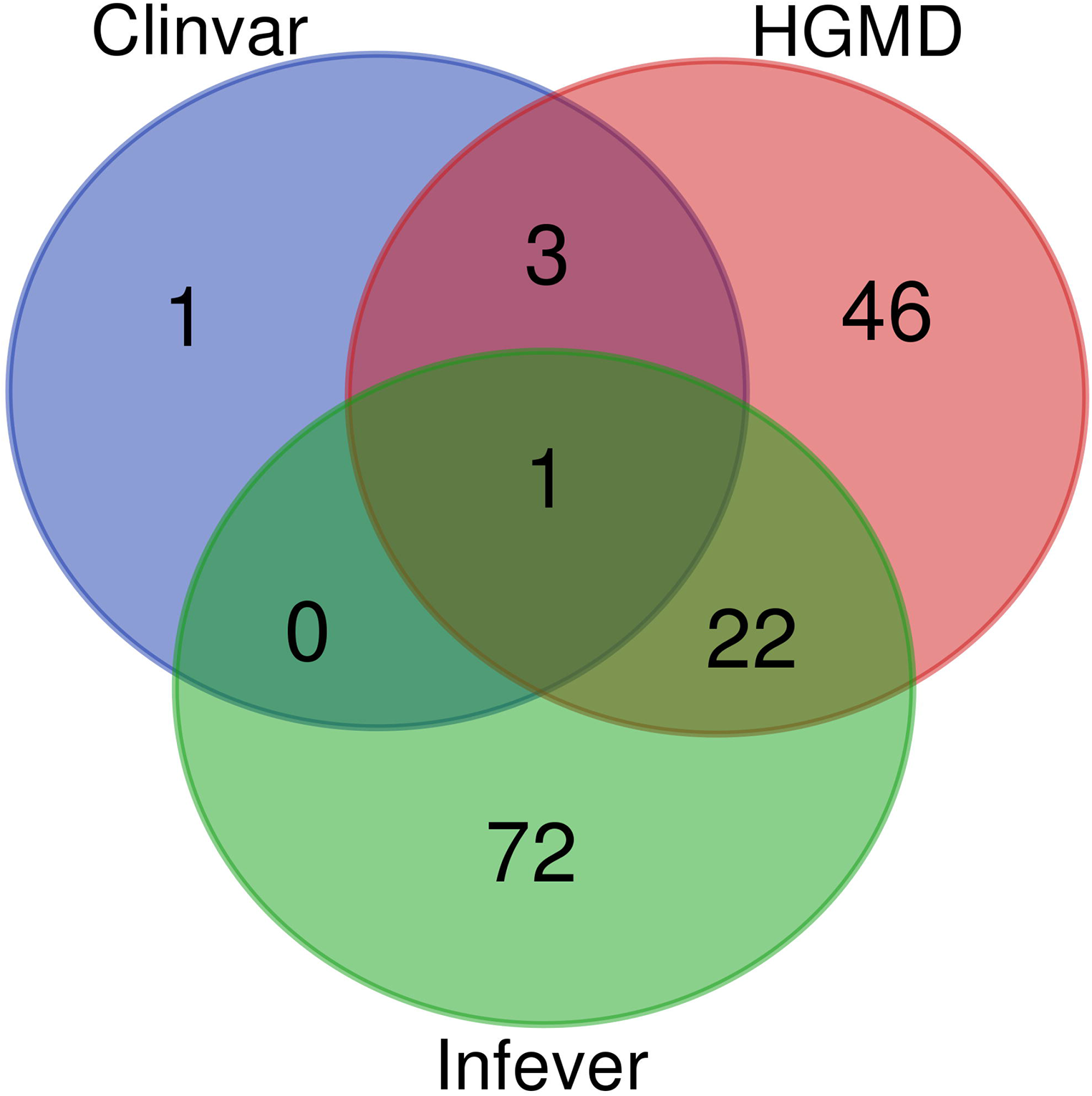
Venn Diagram for the variants present in HGMD, ClinVar and Infevers.

## Results

### Genetic variants in the autoinflammatory genes from different database

We retrieved pathogenic and likely pathogenic genetic variants that encompass a total of 72,331 variants from the ClinVar database. These variants were further filtered based on the list of 37 genes associated with monogenic autoinflammatory diseases resulting in 270 variants. These variants were further overlapped with the Qatar dataset and 5 variants were retrieved. In a similar way, the Infevers database had 1,254 genetic variants from 29 rare autoinflammatory genes excluding *MEFV* [Accessed: 15/11/2017]. These Infevers genetic variants were identified in the Qatar dataset that resulted in an overlap of 95 variants. Similarly, for HGMD, a total of 1,209 genetic variants exist in 37 rare autoinflammatory genes. These variants were overlapped with the Qatar dataset and a total of 72 genetic variants were retrieved.

ClinVar, Infevers, and HGMD had overlapping variants, while three variants were common to ClinVar and HGMD, 22 variants were common to HGMD and Infevers whereas there was no common variant to ClinVar and Infevers. There were also variants that were unique to the database i.e. one variant in ClinVar, 46 in HGMD and 72 in Infevers. So, we had a total of 145 unique variants in Qatar which overlapped with the databases. Venn diagrams for variant numbers and databases are represented in **Figure 1**. The information for the total number of variants from each database and genes has been tabulated in **Supplementary Table 2**.

### Variant Annotation based on ACMG and AMP classification

We retrieved 145 unique Qatari genetic variants which overlapped with three databases. These were classified as one pathogenic and six likely pathogenic variants, while 92 variants were benign and likely benign and 42 were VUS and VUS with conflicting evidence. The pathogenic and likely pathogenic variants are tabulated in **Table 1**. The detailed variant information is provided in **Supplementary Table 3**.

### Comparison of Allele Frequencies with the world population

We implemented Fisher’s exact test to compare the allele frequencies of Qatar and GME population and their subpopulations with the global dataset of gnomAD V2. We identified that five out of seven variants were significantly different (p-value < 0.05) in the Qatari population and its subpopulations and two variants in GME population and its subpopulations. A likely pathogenic variant p.A125V rs5029941 in *TNFAIP3* associated with the familial autoinflammatory, Behcet-like syndrome (OMIM: 616744) was found to be significantly high in Sub-Saharan African subpopulation of Qatar compared to the gnomAD V2. In Qatar Sub-Saharan African population, AF was found to be 1.6%, while in gnomAD V2 its AF was 0.16%. Another likely pathogenic variant p.V377I rs28934897 in the *MVK* gene associated with hyper-IgD syndrome (OMIM: 260920) was significantly enriched in Qatar subpopulation Arab with AF 0.8% compared to the global AF i.e. absent in 1000 genomes, and ∼0.15% in Esp6500, gnomAD V2, and gnomAD V3. Interestingly, this variant was also significantly enriched in GME subpopulation Arabian Peninsula (AP) with AF of 0.87% in comparison to the global frequency. A interesting pathogenic variant p.R82C rs753966933 in *RAB27A* gene associated with hemophagocytic lymphohistiocytosis (HLH) (OMIM number 267700) was found to be significantly enriched in Qatari population and its subpopulation Arab and Bedouin in comparison to the global population. The AF in Qatari population was found to be 0.5% and in Arab and Bedouin subpopulation its frequency is 2.7% and 0.1% respectively. However in the global population databases, this variant is absent in the 1000 Genome project, Esp6500 and gnomAD V3, while having a very low AF in gnomAD V2 i.e. 1.6 × 10^−3^ %. This variant is also absent in the GME population and its subpopulation. A likely pathogenic variant p.R352C in *NLRP12* rs199881207 was significantly enriched in two Qatar subpopulation i.e. European and South Asian whose AF was 12.5% and 0.8% respectively. However, it was absent in the 1000 genomes and had very low frequency in GME, Esp6500, gnomAD V2, and gnomAD V3 i.e. 0.05%, 0.07% 0.04% and 0.02% respectively. Another *NLRP12* pathogenic variant p.R284X was found to be significantly enriched in Qatar and its subpopulation Sub-Saharan African with AF 0.5% and 2.9% respectively in comparison to the global population. The global population had low AF i.e. 0.2%, 0.007%, 0.015%, and 0.04% in 1000 Genome project, Esp6500, gnomAD V2, and gnomAD V3 respectively. Another likely pathogenic variant p.N852S in *NOD2* gene associated with Blau syndrome (OMIM number 186580), Crohn’s disease (OMIM number 266600), and Yao syndrome (OMIM number 617321) was significantly enriched in the GME population and its subpopulation Northeast Africa (NEA) in comparison to the global control population. Its allele frequency in GME and its subpopulation NEA was 0.3% and 0.5% respectively whereas in 1000 genome, Esp6500, gnomAD V2 and gnomAD V3 it was 0.04%, 0.07%, 0.1%, and 0.07% respectively. Comparison of AF of Qatar population and its subpopulation with GME and global population dataset is well represented in **Figure 2**. The comparison of allele frequency of Qatar population among the global population (1000 genome, Esp6500, gnomAD V2 and gnomAD V3) and GME has been detailed in **Supplementary Table 4**.

**Figure 2:**
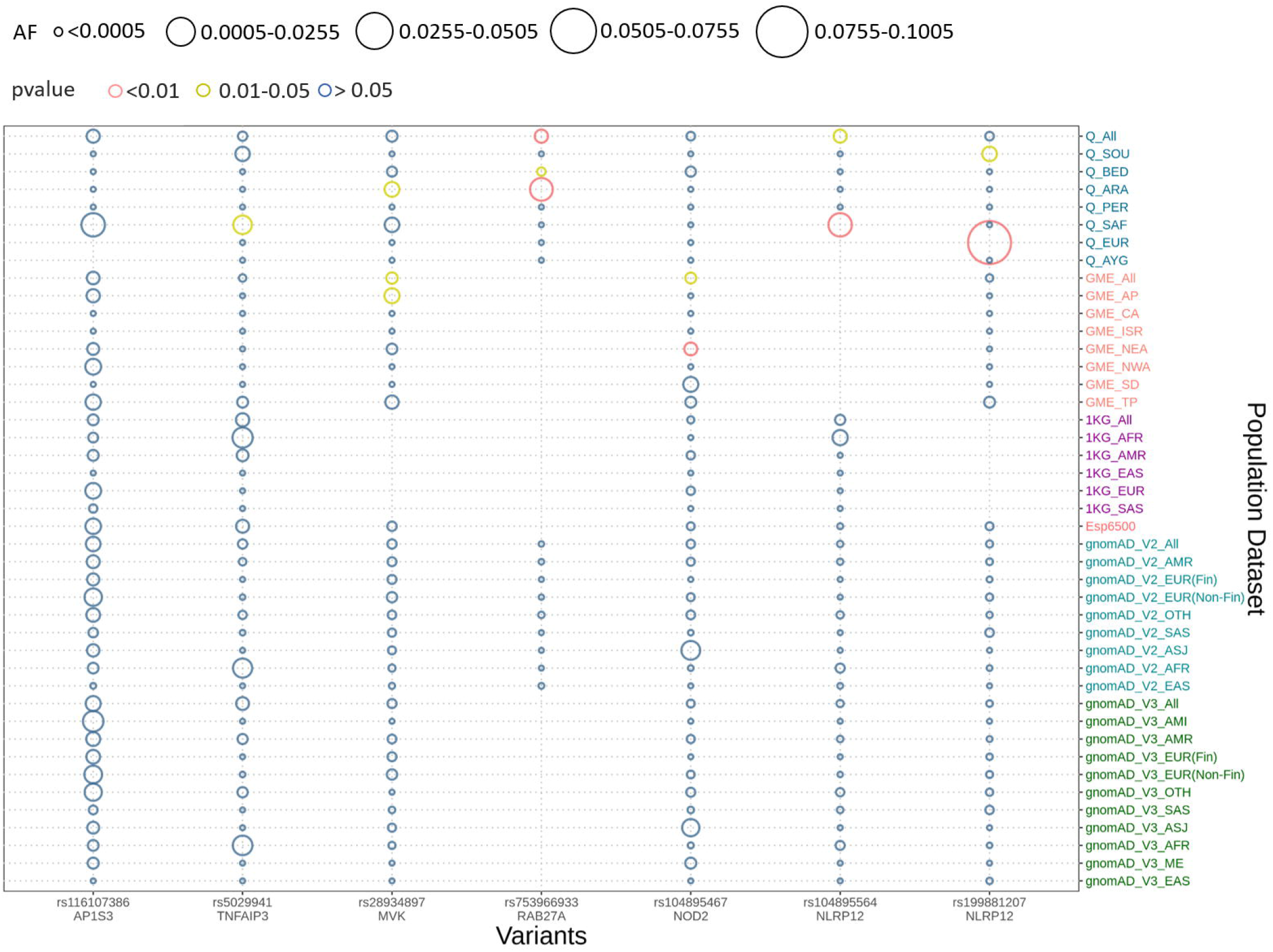
Comparison of allele frequency of pathogenic and likely pathogenic variants in the Qatari population with its subpopulation and GME and its subpopulation with 1000 genome, Esp6500, gnomAD V2 and gnomAD V3 and their subpopulations. Allele frequency highlighted with red for the p-values <0.01, yellow for p-value between 0.01-0.05 and blue for p-value >0.05.

### Genes under natural selection

We explored signals for natural selection in the rare autoinflammatory genes. We took 1% of top iHS scored variants and found that 5 rare autoinflammatory genes fall into top 1% naturally selected variants. These 5 rare autoinflammatory genes were *IL1RN, IL36RN, NLRP3, PSMB9*, and *RAB27A*. **Figure 3** represents iHS and Fst plot for a natural selection of the *RAB27A* gene, we chose *RAB27A* gene as one variant in *RAB27A that had* been annotated as likely pathogenic and had very high carrier allele frequency in comparison to global control population.

**Figure 3:**
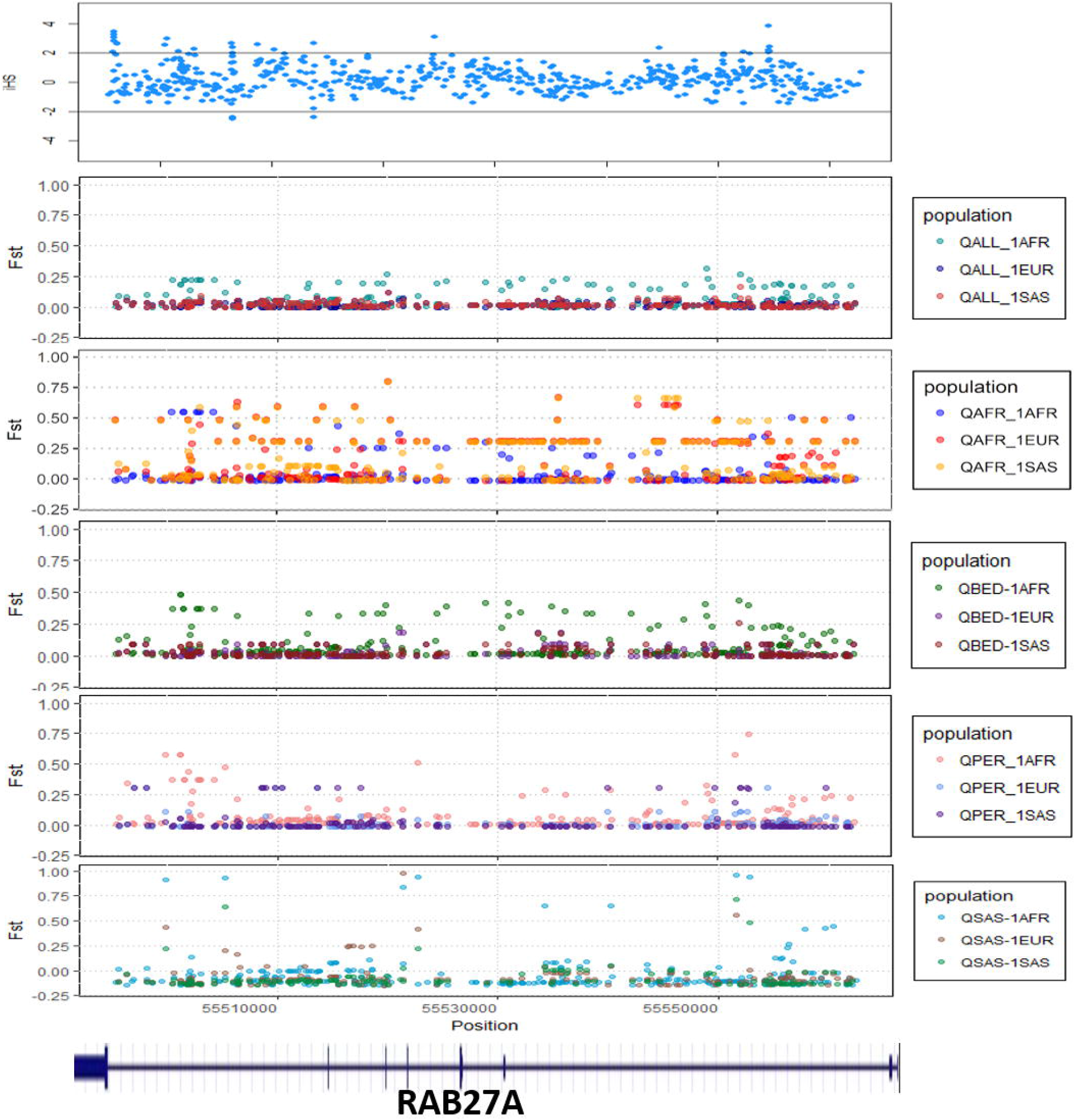
The Fst and iHS score depicted for RAB27A gene from the Qatari population (QALL: Qatar 1005 dataset; 1AFR,1000 Genome AFRICAN; 1EUR, 1000 Genome EUROPEAN; 1SAS, 1000 Genome South Asian; QPER: Qatar-Persian subset; QBED: Qatar-Bedouin subset; QARB: Qatar-Arab-subset; QAFR: Qatar-African-subset; QSAS: Qatar-South-Asian-subset).

## Discussion

The monogenic autoinflammatory disorders are a group of the Mendelian disorders that are caused due to abnormal development and maturation of innate immune cells resulting in the unprovoked recurrent fever with rashes, systemic inflammation, pain in the chest, arthritis, and serositis (McDermott et al., 1999; Cush, 2013). The spectrum of diseases encompasses over 30 distinct monogenic autoinflammatory diseases caused by pathogenic variants in 44 genes as characterized by an expert panel of Inborn Errors of Immunity Committee and Infevers (Touitou et al., 2004; Fakhro et al., 2016). A large number of cases of clinically and molecularly characterized autoinflammatory disorders have been reported in the literature, with the positivity of genetic confirmation of less than 10% that includes variants of uncertain significance (VUS) (Ter Haar et al., 2019). A significant number of autoinflammatory conditions have also been reported in the Middle Eastern population but genetic epidemiology of autoinflammatory disorders is not yet known (Tunca y Ozdogan, 2005; de Jesus et al., 2015; Manthiram et al., 2017; Ter Haar et al., 2019). The available whole genome and whole exome sequence datasets of Middle Eastern populations were used to understand the genetic epidemiology of autoinflammatory disorders in the population (Fakhro et al., 2016; Scott et al., 2016).

In the present study, we compiled a comprehensive list of genes and genetic variants from a number of resources. A total of 37 genes associated with 35 autoinflammatory conditions formed the basis of the analysis (Touitou et al., 2004). We did not consider Familial Mediterranean Fever and *MEFV* gene for the analysis, since an earlier work from our group comprehensively analyzed the genetic epidemiology of the genetic variants for this gene (Koshy et al., 2018). A total 145 unique variants in Qatar autoinflammatory variant, encompassing 917 exomes and 88 genomes were classified as per the ACMG/AMP guidelines for the interpretation of sequence variants. Systematic filtering and annotation by ACMG/AMP guidelines resulted in 7 pathogenic and likely pathogenic variants. The final list was composed of 6 nonsynonymous variants and 1 stop gain variant in 5 genes (one in *MVK*, one in *RAB27A*, one in *NOD2*, one in *AP1S3*, one in *TNFAIP3*, and two in *NLRP12*).

The nonsynonymous variant in *AP1S3* i.e. Phe4Cys (p.F4C) rs116107386 associated with Psoriasis 15 (OMIM number 616106) annotated as likely pathogenic by ACMG guidelines had been functionally validated using transfection studies in HEK293 cells and HaCaT keratinocytes that showed the decreased activity of the mutant protein (Setta-Kaffetzi et al., 2014). The F4C variant is located in the functionally important domain of AP1S3 protein and it has been predicted as deleterious computationally. Another nonsynonymous likely pathogenic variants in *TNFAIP3* i.e. Ala125Val (p.A125V) leads to familial autoinflammatory, Behcet-like syndrome (OMIM number 616744). Lodolce and group transfected HEK293 cell lines with wild type and mutant allele and found a decrease in enzymatic activity of *TNFAIP3* to deubiquitinase the target protein TRAF2 in mutant allele (Lodolce et al., 2010). This variant also falls in a functionally important domain, which affects the functionality of the protein. Third nonsynonymous likely pathogenic variant in the *MVK* i.e. V377I had been reported in various studies as a cause of Hyperimmunoglobulinemia D with Periodic Fever Syndrome (HIDS) (OMIM number 260920). Both of these diseases are rare and follow an autosomal recessive mode of inheritance. This variant was popularly known as the Dutch variant with a carrier frequency of 1:65 individuals (Houten et al., 2003). A number of studies have shown that this variant was present in homozygous or compound heterozygous states in multiple patients affected by HIDS with different ethnic backgrounds. Various in-vitro and in-vivo functional studies have proved p.V377I as disease-causing with very low allele frequency in control databases (Drenth et al., 1999; Houten et al., 1999, 2001; Cuisset et al., 2001; Levy et al., 2013; Govindaraj et al., 2020). Fourth nonsynonymous likely pathogenic variant has been found in *RAB27A* i.e. Arg82Cys (R82C) and had been predicted as a causal variant for hemophagocytic lymphohistiocytosis (HLH) (OMIM number 267700). This variant has very low allele frequency in the global population dataset and is located in a functional domain of the *RAB27A*. In 2016, Netter et al. also reported that a family has been severely affected due to homozygous p.R28C variant and less affected by heterozygous p.R82C (Netter et al., 2016). The functional assay also showed the significant effect of p.R82C on RAB27A binding activity to melanophilin and Munc13-4 (Cetica et al., 2015). Fifth nonsynonymous likely pathogenic variant was in *NOD2* i.e. Asn852Ser (p.N852S) which is associated with Blau syndrome (OMIM number 186580), Crohn’s disease (OMIM number 266600), and Yao syndrome (OMIM number 617321). These syndromes were associated with abnormal inflammation of the body. Horowitz *et al*. in 2017 identified compound heterozygous variants in two affected individuals. The first individual was compound heterozygous for p.G908R and p.N852S and the second was for novel variant p.S506Vfs*73 and p.G908R (Horowitz et al.). The functional assessment had shown that p.N852S led to impaired NOD2 ligand muramyl dipeptide(MDP)-induced NF-κB activation which could lead to Crohn’s disease (Rivas et al., 2011). p.N852S had been co-segregated within affected Ashkenazi Jewish family with Crohn’s disease. It has been reported that 15% of Crohn patients of Ashkenazi Jewish ancestry had the p.N852S variant (Tukel et al., 2004). Sixth nonsynonymous likely pathogenic variant from *NLRP12* i.e. Arg352Cys (p.R352C) is found to be associated with Familial cold autoinflammatory syndrome 2 (OMIM number 611762). Jeru and group in 2011 performed an in-vivo experiment in which they transfected the HEK293 cell line with wild type plasmid and mutant R352C plasmid. They showed that the variant enhanced the signaling activity of procaspase 1 which leads to abnormal inflammation and causes NLRP12-Associated Periodic Fever Syndrome (Jéru et al., 2011). The R352C variant also falls in the functionally important domain of NLRP12. The seventh stop-gain pathogenic variant which falls in *NLRP12* i.e. p.R248X causes Familial cold autoinflammatory syndrome 2 (OMIM number 611762). This stop gain variant leads to aberrant termination of protein synthesis which disrupts the integrity of protein structure and ultimately its function. NLRP12 was responsible to inhibit the inflammatory response but due to the stop-gain variant, it was not regulated which caused abnormal inflammation and periodic fever.

The allele frequencies of 5 out of 7 variants classified as pathogenic or likely pathogenic were significantly different in Qatari and GME population datasets in comparison with global populations. The allele frequencies of pathogenic or likely pathogenic variants showed concordance with another study performed previously in this region. In 2005, Mohammed Hammoudeh reported for the first time an Arab child being affected by HIDS, with the homozygous variant p.V377I in the *MVK* gene (Hammoudeh, 2005). Moussa et al. in 2015 also reported p.V377I segregation with HIDS in an Arabic family where 2 siblings were homozygous p.V377I and affected (Moussa et al., 2015). Also, there were various reports for a patient suffering from HIDS from the Middle East and nearby region which include Turkey, Armenia, Kuwait, Israel, Palestine (van der Hilst et al., 2008; Harel-Meir et al., 2009; Megged et al., 2011; Tas et al., 2012). In the Qatar population dataset, overall carrier allele frequency was 0.32% while the Arab subpopulation had a very high carrier allele frequency of 0.82%. Similarly, in the GME population dataset, carrier allele frequency was 0.35% while in the African Pygmy subpopulation, a high carrier allele frequency of 0.87% was observed. Another pathogenic variant p.R82C in *RAB27A* had a very high carrier allele frequency in Qatari Arab subpopulation i.e. 2.7% while the allele frequency was 0.5% in the Qatari population dataset. A large study was conducted by Tukel et al. in 2004 to find the frequency of Crohn disease-associated Ashkenazi, Sephardi, and Oriental Jewish Families in middle eastern countries which include Ashkenazi, Sephardi and Oriental Jewish from Israel, Morocco, Turkey, Tunisia, Iraq, Kurds, Iran, Yemen, and Syria. They found 15% of Ashkenazi Jews suffering from Crohn’s disease had the p.N852S variant (Tukel et al., 2004). Another study that involves p.N852S frequency in the Turkish population, but could not find the N852S association with Crohn’s disease (Budak Diler y Yaraş, 2018). In our study we found p.N852S had high carrier allele frequency in GME subpopulations as 0.8% in Syrian Desert, 0.5% in North-East Africa and 0.3% in Turkish Peninsula and Qatar allele frequency was 0.1% and its subpopulation Bedouin was 0.2%.

The distinct frequencies of many variants prompted us to explore whether any of these genes showed signals of natural selection. We have adopted two parameters i.e. Wright Fixation score (Fst) and integrated Haplotype (iHS) score to understand the signal of the selection in the rare autoinflammatory genes. Our analysis revealed the *RAB27A* gene has high values of the Fst score in the Qatari subpopulation Africa in comparison to the global population. This marks the genetic differences of the Qatari subpopulation Africa with the global population as well as other Qatari subpopulations. Also the positive iHS score for the multiple variants in the *RAB27A* gene represents the signal of selection of *RAB27A* gene in the Qatari population. RAB27A protein is mainly involved in the transportation of melanin for skin pigmentation. A previously reported study has shown a high correlation between *RAB27A* gene selection to the dark-pigmented melanocytes in comparison to the lightly-pigmented melanocytes. This makes *RAB27A* gene a possible candidate to be positively selected for individuals of African descent in comparison to the Caucasian (Yoshida-Amano et al. 2012). It also plays an important role in cytotoxic T lymphocytes for killing or neutralization of foreign invaders. *RAB27A* had been evolutionarily selected among different species, as editing levels of *RAB27A* showed the highest difference among humans and rhesus monkeys (Paz-Yaacov et al., 2010). Similarly, *IL1RN* was identified to be under selection in the European-American population (Akey et al., 2004; Jaffe et al., 2013) whereas *IL36RN* was found to be evolutionary conserved among different species (Lv et al., 2016). Other genes like NLRP3 and PSMB9 did not show any strong signals of natural selection or conservation.

## Conclusion

In summary, our analysis integrating population-scale genomic datasets provide the first comprehensive insights into the genetic epidemiology of monogenic autoinflammatory diseases in the Middle Eastern populations. Our analysis suggests distinct allele frequencies in subpopulations and also suggests that at least two genes show strong signals of natural selection in the middle eastern population. As more population-scale genomic datasets from the region become available, including from national initiatives like the Bahrain genome program and the Emirati genome initiative apart from the ongoing Qatar Genome project, would provide a higher resolution towards understanding frequencies of rarer genetic variants (Al-Ali et al., 2018).

## Data Availability

The data that support the findings of this study are available from the NCBI Sequence Read Archive (SRA accessions SRP060765, SRP061943 and SRP061463, accessible online at http://www.ncbi.nlm.nih.gov/Traces/study/?acc=SRP060765%2CSRP061943%2CSRP061463&go=go. (SRA accession SRP061943).

## Author contributions

Parul Sharma collected the data. Abhinav Jain performed the ACMG annotations, data analysis, and allele frequency comparison. Vinod Scaria oversaw the analysis. All authors contributed to writing the manuscript.

## Acknowledgments

Authors thank Disha Sharma, Mukta Poojary, Bani Jolly, and Vishu Gupta for suggestions that enriched the manuscript. The authors acknowledge funding from the Council of Scientific and Industrial Research (CSIR, India) through Grant MLP1809 (IndiGen). Abhinav Jain is a recipient of Senior Research Fellowship from Council of Scientific and Industrial Research (CSIR, India). The funders had no role in the preparation of the manuscript or decision to submit. We acknowledge the researchers at Weill Cornell Medicine for sharing the genome and exome data sets without which this analysis was not possible.

## Ethical standards

This article does not contain any studies with human participants or animals performed by any of the authors.

## Supplementary Material

**Supplementary Data 1:** The ACMG-AMP guidelines for each attribute

**Supplementary Figure 1:** Schematic summarising the data analysis pipeline utilized for this study.

**Supplementary Table 1**. Genes and associated autoinflammatory diseases considered in the present analysis and their respective inheritance patterns.

**Supplementary Table 2:** The total number of variants in the auto inflammatory genes from each database overlapped with the Qatar dataset

**Supplementary Table 3:** The detailed variant annotation and classification based on ACMG-AMP guidelines

**Supplementary Table 4:**. Allele frequency of pathogenic and likely pathogenic variants compared with control global population (1000 genome, gnomAD V2, gnomAD V3 and Esp6500) using Fisher exact test (p<0.05). BED Bedouin, SAF Sub-Saharan African, EUR European, SOU South Asian, APY African Pygmy, ARA ARAB, PER Persian, NA not applicable]. Significant values are marked with *

## Data Availability

The datasets for this study can be found in the NCBI Sequence Read Archive (SRA accessions SRP060765, SRP061943 and SRP061463, accessible online at http://www.ncbi.nlm.nih.gov/Traces/study/?acc=SRP060765%2CSRP061943%2CSRP061463&go=go. (SRA accession SRP061943).

