## Supplementary figures and images for "Genetic landscape of rare autoinflammatory disease variants in Qatar and Middle Eastern populations through the integration of whole-genome and exome datasets"

### Supplementary Figure 1

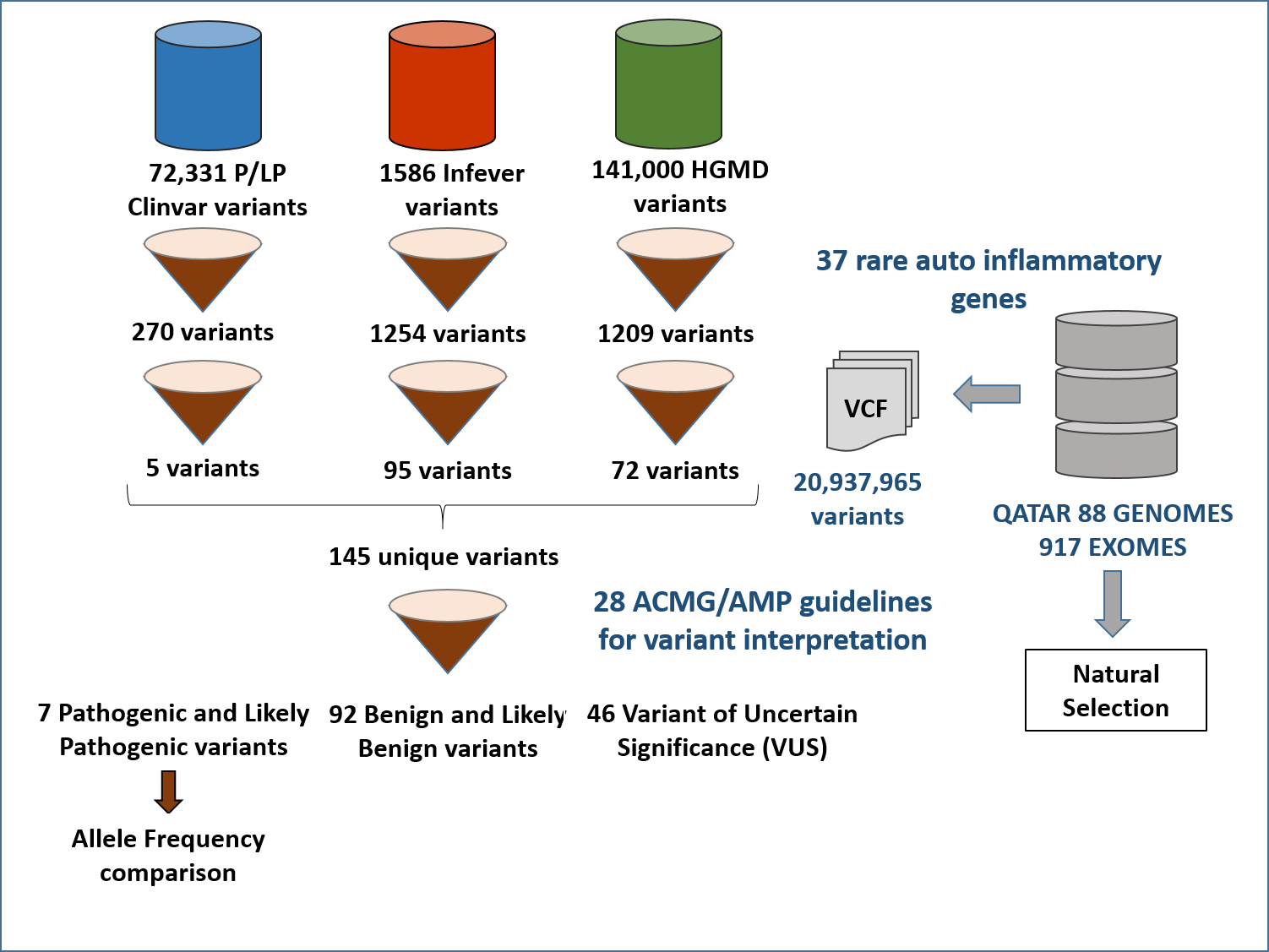
