## Supplementary Table 1 for "Genetic landscape of rare autoinflammatory disease variants in Qatar and Middle Eastern populations through the integration of whole-genome and exome datasets"

**Supplementary Table 1.** Genes and associated autoinflammatory diseases considered in the present analysis and their respective inheritance patterns.

| Disease Name | Gene | Inheritance |
|---|---|---|
| Familial Cold Autoinflammatory Syndrome (FCAS) | *NLRP3* | Autosomal Dominant |
| Muckle-Wells syndrome (MWS) | *NLRP3* | Autosomal Dominant |
| Neonatal onset multi-systemic inflammatory disorder/ Chronic Infantile Neurological Cutaneous Articular Syndrome (NOMID/CINCA) | *NLRP3* | Autosomal Dominant |
| TNF receptor-associated periodic syndrome (TRAPS) | *TNFRSF1A* | Autosomal Dominant |
| Hyperimmunoglobulinemia D with Periodic Fever Syndrome (HIDS) | *MVK* | Autosomal Recessive |
| Mevalonate Aciduria (MA) | *MVK* | Autosomal Recessive |
| Porokeratosis 3, multiple types | *MVK* | Autosomal Dominant |
| Deficiency of Interleukin-1ß (IL-1ß) Receptor Antagonist/ Osteomyelitis, Sterile Multifocal w/Periostitis Pustulosis (DIRA/OMPP) | *IL1RN* | Autosomal Recessive |
| MAJEED/ Chronic Recurrent Multifocal Osteomyelitis, Congenital Dyserythropoietic Anemia, & Neutrophilic Dermatosis | *LPIN2* | Autosomal Recessive |
| Deficiency of Interleukin-36-Receptor Antagonist/ Generalized Pustular Psoriasis (GPP)/ (DITRA/PSORP) | *IL36RN* | Autosomal Recessive |
| Familial Psoriasis/ CARD14-Mediated Pustular Psoriasis (CAMPS/PSORS2) | *CARD14* | Autosomal Dominant |
| Pityriasis rubra pilaris | *CARD14* | Autosomal Dominant |
| Pyogenic Sterile Arthritis, Pyoderma Gangrenosum and Acne Syndrome (PAPA) | *PSTPIP1* | Autosomal Dominant |
| Juvenile Systemic Granulomatosis– Blau syndrome, Pediatric Granulomatous Arthritis (PGA), Early Onset Sarcoidosis, or Jabs Syndrome (BLAU/PGA/EOS) | *NOD2* | Autosomal Dominant |
| NLRP12-Associated Periodic Fever Syndrome/ Familial Cold Autoinflammatory Syndrome 2, or Guadaloupe Periodic Fever (NLRP12/FCAS2) | *NLRP12* | Autosomal Dominant |

| | | |
|---|---|---|
| Chronic Atypical Neutrophilic Dermatosis w/ Lipodystrophy and Elevated Temperature- Nakajo Nishimura Syndrome (CANDLE/PRAAS) | *PSMB8; also PSMB4, PSMB9, PSMA3, POMP* | Autosomal Recessive |
| (Primary) Familial Hemophagocytic Lymphohistiocytosis/ a Familial Erythrophagocytic Lymphohistiocytosis (1°HLH/FHL) | *PRF1, STX11, STXBP2, MUNC13-4/UNC13D, RAB27A* | Autosomal recessive |
| (Primary) Familial Hemophagocytic Lymphohistiocytosis/ a Familial Erythrophagocytic Lymphohistiocytosis (1°HLH/FHL) | *SH2D1A,, BIRC4/XIAP* | X-linked recessive |
| PLCG2-associated Antibody Deficiency and Immune Dysregulation/ Familial Atypical Cold Urticaria (FACU) (PLAID/FCAS3) | *PLCG2* | Autosomal Dominant |
| Autoinflammation and PLCG2-associated Antibody Deficiency and Immune Dysregulation (APLAID) | *PLCG2* | Autosomal Dominant |
| SLC29A3 Spectrum Disorder (SLC29A3) | *SLC29A3* | Autosomal Recessive |
| Psoriasis 15, Pustular | *AP1S3* | Autosomal Dominant |
| Deficiency of Adenosine Deaminase 2 (DADA2) | *CECR1/ADA2* | Autosomal Recessive |
| Sneddon syndrome | *CECR1/ADA2* | Autosomal Recessive |
| Interleukin 10 deficiency (IL10D) | *IL10* | Autosomal Recessive |
| Inflammatory bowel disease 28 (IBD28)/ Interleukin 10 receptor A deficiency (IL10R1D) | *IL10RA* | Autosomal Recessive |
| Inflammatory bowel disease 25 (IBD25)/ Interleukin 10 receptor B deficiency (IL10R2D) | *IL10RB* | Autosomal Recessive |
| Autoinflammation with infantile enterocolitis (AIFEC) | *NLRC4* | Autosomal Dominant |
| Hydatidiform Mole, Recurrent, 1 (HYDM1) | *NLRP7* | Autosomal Recessive |
| Otulipenia | *OTULIN* | Autosomal Recessive |
| Polyglucosan Body Myopathy, Early-onset, with Or without Immunodeficiency (PBMEI) | *RBCK1* | Autosomal Recessive |
| *Cherubism* | *SH3BP2* | Autosomal Dominant |

| | | |
|---|---|---|
| Sting-Associated Vasculopathy, Infantile-Onset (SAVI) | *TMEM173* | Autosomal Dominant |
| Autoinflammatory syndrome, familial, Behcet-like (AISBL) | *TNFAIP3* | Autosomal Dominant |
| TNFRSF11A-associated hereditary fever disease (TRAPS11) | TNFRSF11A | Autosomal Dominant |
