## Supplementary Table 2 for "Genetic landscape of rare autoinflammatory disease variants in Qatar and Middle Eastern populations through the integration of whole-genome and exome datasets"

**Supplementary Table 2:** The total number of variants in the auto inflammatory genes from each database overlapped with the Qatar dataset

| Genes | Clinvar | Infever | HGMD |
|---|---|---|---|
| AP1S3 | 0 | 0 | 2 |
| NLRP3 | 0 | 33 | 4 |
| TNFRSF1A | 1 | 0 | 2 |
| MVK | 1 | 16 | 3 |
| IL1RN | 0 | 5 | 0 |
| LPIN2 | 0 | 0 | 6 |
| IL36RN | 0 | 1 | 1 |
| CARD14 | 0 | 8 | 1 |
| PSTPIP1 | 0 | 3 | 1 |
| NOD2 | 1 | 27 | 14 |
| NLRP12 | 1 | 0 | 5 |
| PSMB8 | 0 | 0 | 1 |
| PSMB4 | 0 | 0 | 0 |
| PSMB9 | 0 | 0 | 2 |
| PSMA3 | 0 | 0 | 0 |
| POMP | 0 | 0 | 0 |
| PRF1 | 0 | 0 | 7 |
| STX11 | 0 | 0 | 0 |
| STXBP2 | 0 | 0 | 3 |
| MUNC13 or UNC13D | 0 | 0 | 5 |
| RAB27A | 1 | 0 | 1 |
| SH2D1A | 0 | 0 | 0 |
| BIRC4 or XIAP | 0 | 0 | 0 |
| PLCG2 | 0 | 0 | 0 |
| SLC29A3 | 0 | 0 | 0 |
| CECR1 or ADA2 | 0 | 0 | 0 |
| IL10 | 0 | 0 | 0 |
| IL10RA | 0 | 1 | 1 |
| IL10RB | 0 | 0 | 0 |
| NLRC4 | 0 | 0 | 0 |
| NLRP7 | 0 | 0 | 9 |
| OTULIN | 0 | 0 | 0 |
| RBCK1 | 0 | 0 | 0 |
| SH3BP2 | 0 | 1 | 0 |
| TMEM173 | 0 | 0 | 2 |
| TNFAIP3 | 0 | 0 | 2 |
| TNFRSF11A | 0 | 0 | 0 |
| **Total** | **5** | **95** | **72** |
