## Supplementary Table 3 for "Genetic landscape of rare autoinflammatory disease variants in Qatar and Middle Eastern populations through the integration of whole-genome and exome datasets"

**Supplementary Table 3:** The detailed variant annotation and classification based on ACMG-AMP guidelines

| Chr | Start | End | Ref | Alt | Func. refGene | Gene. refGene | GeneDetail.refGene | CLINSIG 20180930 | CLNDBN | esp6500si all | ExAC ALL | 1000g2015aug all | gnomAD exome ALL | Qatar AF | SIFT pred | Polyphen2 HVAR pred | CADD phred | ExonicFunc refGene | AAChange refGene |
|---|---|---|---|---|---|---|---|---|---|---|---|---|---|---|---|---|---|---|---|
| 5 | 138856982 | 138856982 | C | T | exonic | TMEM173 | . | | . | 0.183454 | 0.2049 | 0.27496 | 0.2117 | 0.2149 | D | D | 28.4 | nonsynonymous SNV | TMEM173:NM_198282:exon7:c.G878A:p.R293Q |
| 5 | 138861078 | 138861078 | C | T | exonic | TMEM173 | . | | . | 0.104798 | 0.1834 | 0.203075 | 0.1946 | 0.1896 | T | B | 13.08 | nonsynonymous SNV | TMEM173:NM_001301738:exon2:c.G212A:p.R71H,TMEM173:NM_198282:exon3:c.G212A:p.R71H |
| 6 | 32811629 | 32811629 | G | T | exonic | PSMB8 | . | Benign/Likely_benign | Nakajo_syndrome\|not_specified | 0.127105 | 0.1453 | 0.151957 | 0.1438 | 0.06818 | T | B | 22.2 | nonsynonymous SNV | PSMB8:NM_148919:exon1:c.C145A:p.Q49K |
| 6 | 32825090 | 32825090 | G | A | exonic | PSMB9 | . | | . | 0.248045 | 0.2456 | 0.224441 | 0.2432 | 0.3129 | T | B | 14.26 | nonsynonymous SNV | PSMB9:NM_002800:exon3:c.G179A:p.R60H |
| 6 | 138196066 | 138196066 | T | G | exonic | TNFAIP3 | . | not_provided | not_specified | 0.141934 | 0.0613 | 0.139577 | 0.0534 | 0.05672 | T | B | 15.6 | nonsynonymous SNV | TNFAIP3:NM_001270507:exon3:c.T380G:p.F127C,TNFAIP3:NM_001270508:exon3:c.T380G:p.F127C,TNFAIP3:NM_006290:exon3:c.T380G:p.F127C |
| 11 | 117864063 | 117864063 | A | G | exonic | IL10RA | . | Benign/Likely_benign | not_specified\|Inflammatory_bowel_disease | 0.119458 | 0.1316 | 0.0816693 | 0.1309 | 0.08657 | T | B | 11.09 | nonsynonymous SNV | IL10RA:NM_001558:exon4:c.A475G:p.S159G |
| 11 | 117869670 | 117869670 | A | G | exonic | IL10RA | . | Benign | Inflammatory_bowel_disease\|not_specified | 0.728448 | 0.7381 | 0.811102 | 0.7416 | 0.2473 | T | B | 0.945 | nonsynonymous SNV | IL10RA:NM_001558:exon7:c.A1051G:p.R351G |
| 12 | 110013879 | 110013879 | G | A | exonic | MVK | . | Benign/Likely_benign | not_specified\|Mevalonic_aciduria\|Hyperimmunoglobulin_D_with_periodic_fever\|Methylmalonic_acidemia | 0.116177 | 0.1017 | 0.0758786 | 0.101 | 0.1124 | T | B | 0.002 | nonsynonymous SNV | MVK:NM_000431:exon3:c.G155A:p.S52N,MVK:NM_001114185:exon3:c.G155A:p.S52N,MVK:NM_001301182:exon3:c.G155A:p.S52N |
| 16 | 50744624 | 50744624 | C | T | exonic | NOD2 | . | Benign | Blau_syndrome\|Crohn_disease\|not_specified | 0.197676 | 0.1844 | 0.102037 | 0.1889 | 0.1867 | T | B | 0.001 | nonsynonymous SNV | NOD2:NM_001293557:exon3:c.C721T:p.P241S,NOD2:NM_022162:exon4:c.C802T:p.P268S |
| 16 | 50757276 | 50757276 | G | A | exonic | NOD2 | . | Benign | Sarcoidosis,_early-onset\|Crohn_disease\|Blau_syndrome | 0.069021 | 0.0611 | 0.0333466 | 0.0626 | 0.09801 | D | B | 1.382 | nonsynonymous SNV | NOD2:NM_001293557:exon8:c.G2782A:p.V928I,NOD2:NM_022162:exon9:c.G2863A:p.V955I |
| 17 | 78171944 | 78171944 | G | C | exonic | CARD14 | . | Benign | not_specified | 0.359142 | 0.3772 | 0.370208 | 0.372 | 0.4488 | T | B | 0.077 | nonsynonymous SNV | CARD14:NM_052819:exon9:c.G930C:p.R310S,CARD14:NM_001257970:exon12:c.G1641C:p.R547S,CARD14:NM_024110:exon12:c.G1641C:p.R547S |
| 17 | 78178893 | 78178893 | C | T | exonic | CARD14 | . | Benign | not_specified | 0.409322 | 0.4244 | 0.353035 | 0.4143 | 0.3941 | D | B | 25.8 | nonsynonymous SNV | CARD14:NM_024110:exon18:c.C2458T:p.R820W |
| 19 | 55450727 | 55450727 | C | T | exonic | NLRP7 | . | Benign/Likely_benign | Hydatidiform_mole\|not_specified | 0.141781 | 0.0586 | 0.125599 | 0.0523 | 0.09353 | T | B | 0.086 | nonsynonymous SNV | NLRP7:NM_001127255:exon4:c.G1460A:p.G487E,NLRP7:NM_139176:exon4:c.G1460A:p.G487E,NLRP7:NM_206828:exon4:c.G1460A:p.G487E |
| 19 | 55450746 | 55450746 | C | T | exonic | NLRP7 | . | Benign/Likely_benign | not_specified\|Hydatidiform_mole | 0.117484 | 0.0888 | 0.0908546 | 0.0869 | 0.1193 | T | B | 6.577 | nonsynonymous SNV | NLRP7:NM_001127255:exon4:c.G1441A:p.A481T,NLRP7:NM_139176:exon4:c.G1441A:p.A481T,NLRP7:NM_206828:exon4:c.G1441A:p.A481T |
| 1 | 247582063 | 247582063 | G | T | UTR5 | NLRP3 | NM_001079821:c.-34G>T;NM_001127461:c.-34G>T;NM_001127462:c.-34G>T;NM_183395:c.-34G>T;NM_001243133:c.-40G>T;NM_004895:c.-34G>T | Benign | Chronic_infantile_neurological,_cutaneous_and_articular_syndrome\|Familial_cold_autoinflammatory_syndrome\|Familial_amyloid_nephropathy_with_urticaria_AND_deafness | 0.041827 | 0.0367 | 0.0269569 | 0.0374 | 0.02841 | . | . | . | . | . |
| 1 | 247582309 | 247582309 | C | T | exonic | NLRP3 | . | Benign/Likely_benign | not_specified\|Chronic_infantile_neurological,_cutaneous_and_articular_syndrome\|Familial_cold_autoinflammatory_syndrome\|Familial_amyloid_nephropathy_with_urticaria_AND_deafness | 0.000077 | 0.0016 | 0.00319489 | 0.0016 | 0.002931 | . | . | . | synonymous SNV | NLRP3:NM_001127462:exon1:c.C213T:p.A71A,NLRP3:NM_001243133:exon1:c.C207T:p.A69A,NLRP3:NM_004895:exon1:c.C213T:p.A71A,NLRP3:NM_183395:exon1:c.C213T:p.A71A,NLRP3:NM_001127461:exon2:c.C213T:p.A71A,NLRP3:NM_001079821:exon3:c.C213T:p.A71A |
| 1 | 247586336 | 247586336 | T | C | intronic | NLRP3 | . | | . | 0 | 0 | 0.459665 | 0 | 0.4375 | . | . | . | . | . |
| 1 | 247586920 | 247586920 | T | G | intronic | NLRP3 | . | | . | 0 | 0 | 0.919728 | 0 | 0.04545 | . | . | . | . | . |
| 1 | 247587174 | 247587174 | C | T | exonic | NLRP3 | . | Benign/Likely_benign | Familial_cold_autoinflammatory_syndrome\|Chronic_infantile_neurological,_cutaneous_and_articular_syndrome\|Familial_amyloid_nephropathy_with_urticaria_AND_deafness | 0.006459 | 0.0021 | 0.00798722 | 0.0016 | 0.001601 | . | . | . | synonymous SNV | NLRP3:NM_001127462:exon3:c.C429T:p.Y143Y,NLRP3:NM_001243133:exon3:c.C423T:p.Y141Y,NLRP3:NM_004895:exon3:c.C429T:p.Y143Y,NLRP3:NM_183395:exon3:c.C429T:p.Y143Y,NLRP3:NM_001127461:exon4:c.C429T:p.Y143Y,NLRP3:NM_001079821:exon5:c.C429T:p.Y143Y |
| 1 | 247587408 | 247587408 | C | T | exonic | NLRP3 | . | Benign | not_specified\|Familial_amyloid_nephropathy_with_urticaria_AND_deafness\|Familial_cold_autoinflammatory_syndrome\|Chronic_infantile_neurological,_cutaneous_and_articular_syndrome | 0.115254 | 0.0817 | 0.116613 | 0.0797 | 0.09602 | . | . | . | synonymous SNV | NLRP3:NM_001127462:exon3:c.C663T:p.T221T,NLRP3:NM_001243133:exon3:c.C657T:p.T219T,NLRP3:NM_004895:exon3:c.C663T:p.T221T,NLRP3:NM_183395:exon3:c.C663T:p.T221T,NLRP3:NM_001127461:exon4:c.C663T:p.T221T,NLRP3:NM_001079821:exon5:c.C663T:p.T221T |
| 1 | 247587477 | 247587477 | G | A | exonic | NLRP3 | . | Benign | not_specified\|Chronic_infantile_neurological,_cutaneous_and_articular_syndrome\|Familial_cold_autoinflammatory_syndrome\|Familial_amyloid_nephropathy_with_urticaria_AND_deafness | 0.405966 | 0.4996 | 0.38738 | 0.5034 | 0.4881 | . | . | . | synonymous SNV | NLRP3:NM_001127462:exon3:c.G732A:p.A244A,NLRP3:NM_001243133:exon3:c.G726A:p.A242A,NLRP3:NM_004895:exon3:c.G732A:p.A244A,NLRP3:NM_183395:exon3:c.G732A:p.A244A,NLRP3:NM_001127461:exon4:c.G732A:p.A244A,NLRP3:NM_001079821:exon5:c.G732A:p.A244A |
| 1 | 247587681 | 247587681 | C | T | exonic | NLRP3 | . | Benign/Likely_benign | Familial_cold_urticaria\|not_specified | 0.001153 | 0.0012 | 0.000998403 | 0.0012 | 0.001172 | . | . | . | synonymous SNV | NLRP3:NM_001127462:exon3:c.C936T:p.D312D,NLRP3:NM_001243133:exon3:c.C930T:p.D310D,NLRP3:NM_004895:exon3:c.C936T:p.D312D,NLRP3:NM_183395:exon3:c.C936T:p.D312D,NLRP3:NM_001127461:exon4:c.C936T:p.D312D,NLRP3:NM_001079821:exon5:c.C936T:p.D312D |
| 1 | 247587771 | 247587771 | C | T | exonic | NLRP3 | . | Benign/Likely_benign | not_specified\|Familial_cold_autoinflammatory_syndrome | 0.006151 | 0.0049 | 0.00179712 | 0.0052 | 0.001761 | . | . | . | synonymous SNV | NLRP3:NM_001127462:exon3:c.C1026T:p.P342P,NLRP3:NM_001243133:exon3:c.C1020T:p.P340P,NLRP3:NM_004895:exon3:c.C1026T:p.P342P,NLRP3:NM_183395:exon3:c.C1026T:p.P342P,NLRP3:NM_001127461:exon4:c.C1026T:p.P342P,NLRP3:NM_001079821:exon5:c.C1026T:p.P342P |
| 1 | 247587783 | 247587783 | G | A | exonic | NLRP3 | . | Benign/Likely_benign | Familial_cold_urticaria\|Familial_amyloid_nephropathy_with_urticaria_AND_deafness\|Familial_cold_autoinflammatory_syndrome\|Chronic_infantile_neurological,_cutaneous_and_articular_syndrome | 0.000538 | 0.0015 | 0.00239617 | 0.0013 | 0.002137 | . | . | . | synonymous SNV | NLRP3:NM_001127462:exon3:c.G1038A:p.L346L,NLRP3:NM_001243133:exon3:c.G1032A:p.L344L,NLRP3:NM_004895:exon3:c.G1038A:p.L346L,NLRP3:NM_183395:exon3:c.G1038A:p.L346L,NLRP3:NM_001127461:exon4:c.G1038A:p.L346L,NLRP3:NM_001079821:exon5:c.G1038A:p.L346L |
| 1 | 247587795 | 247587795 | G | A | exonic | NLRP3 | . | Benign/Likely_benign | not_specified\|Chronic_infantile_neurological,_cutaneous_and_articular_syndrome\|Familial_amyloid_nephropathy_with_urticaria_AND_deafness\|Familial_cold_autoinflammatory_syndrome | 0.000077 | 0.0061 | 0.0115815 | 0.0055 | 0.004706 | . | . | . | synonymous SNV | NLRP3:NM_001127462:exon3:c.G1050A:p.T350T,NLRP3:NM_001243133:exon3:c.G1044A:p.T348T,NLRP3:NM_004895:exon3:c.G1050A:p.T350T,NLRP3:NM_183395:exon3:c.G1050A:p.T350T,NLRP3:NM_001127461:exon4:c.G1050A:p.T350T,NLRP3:NM_001079821:exon5:c.G1050A:p.T350T |
| 1 | 247587982 | 247587982 | C | T | exonic | NLRP3 | . | Benign/Likely_benign | Familial_cold_urticaria\|not_specified\|Chronic_infantile_neurological,_cutaneous_and_articular_syndrome\|Familial_cold_autoinflammatory_syndrome\|Familial_amyloid_nephropathy_with_urticaria_AND_deafness | 0.007304 | 0.0061 | 0.00379393 | 0.0062 | 0.001172 | . | . | . | synonymous SNV | NLRP3:NM_001127462:exon3:c.C1237T:p.L413L,NLRP3:NM_001243133:exon3:c.C1231T:p.L411L,NLRP3:NM_004895:exon3:c.C1237T:p.L413L,NLRP3:NM_183395:exon3:c.C1237T:p.L413L,NLRP3:NM_001127461:exon4:c.C1237T:p.L413L,NLRP3:NM_001079821:exon5:c.C1237T:p.L413L |
| 1 | 247587996 | 247587996 | C | T | exonic | NLRP3 | . | Likely_benign | Chronic_infantile_neurological,_cutaneous_and_articular_syndrome\|Familial_cold_autoinflammatory_syndrome\|Familial_amyloid_nephropathy_with_urticaria_AND_deafness | 0.000077 | 6.59E-05 | 0.000199681 | 8.13E-05 | 0.007812 | . | . | . | synonymous SNV | NLRP3:NM_001127462:exon3:c.C1251T:p.I417I,NLRP3:NM_001243133:exon3:c.C1245T:p.I415I,NLRP3:NM_004895:exon3:c.C1251T:p.I417I,NLRP3:NM_183395:exon3:c.C1251T:p.I417I,NLRP3:NM_001127461:exon4:c.C1251T:p.I417I,NLRP3:NM_001079821:exon5:c.C1251T:p.I417I |
| 1 | 247588053 | 247588053 | C | T | exonic | NLRP3 | . | Benign | not_specified\|Familial_cold_autoinflammatory_syndrome\|Familial_amyloid_nephropathy_with_urticaria_AND_deafness\|Chronic_infantile_neurological,_cutaneous_and_articular_syndrome | 0.088959 | 0.101 | 0.0656949 | 0.1013 | 0.1507 | . | . | . | synonymous SNV | NLRP3:NM_001127462:exon3:c.C1308T:p.S436S,NLRP3:NM_001243133:exon3:c.C1302T:p.S434S,NLRP3:NM_004895:exon3:c.C1308T:p.S436S,NLRP3:NM_183395:exon3:c.C1308T:p.S436S,NLRP3:NM_001127461:exon4:c.C1308T:p.S436S,NLRP3:NM_001079821:exon5:c.C1308T:p.S436S |
| 1 | 247588351 | 247588351 | C | T | exonic | NLRP3 | . | Benign/Likely_benign | Familial_cold_urticaria\|not_specified\|Chronic_infantile_neurological,_cutaneous_and_articular_syndrome\|Familial_amyloid_nephropathy_with_urticaria_AND_deafness\|Familial_cold_autoinflammatory_syndrome | 0.004768 | 0.0016 | 0.00678914 | 0.0013 | 0.001172 | . | . | . | synonymous SNV | NLRP3:NM_001127462:exon3:c.C1606T:p.L536L,NLRP3:NM_001243133:exon3:c.C1600T:p.L534L,NLRP3:NM_004895:exon3:c.C1606T:p.L536L,NLRP3:NM_183395:exon3:c.C1606T:p.L536L,NLRP3:NM_001127461:exon4:c.C1606T:p.L536L,NLRP3:NM_001079821:exon5:c.C1606T:p.L536L |
| 1 | 247588960 | 247588960 | G | A | intronic | NLRP3 | . | | . | 0 | 0 | 0.130591 | 0 | 0.1193 | . | . | . | . | . |
| 1 | 247593142 | 247593142 | A | G | intronic | NLRP3 | . | | . | 0 | 0 | 0.123003 | 0 | 0.1193 | . | . | . | . | . |
| 1 | 247597180 | 247597180 | T | C | intronic | NLRP3 | . | | . | 0 | 0 | 0 | 0 | 0.1529 | . | . | . | . | . |
| 1 | 247597181 | 247597181 | G | A | intronic | NLRP3 | . | | . | 0 | 0 | 0 | 0 | 0.1529 | . | . | . | . | . |
| 1 | 247597208 | 247597208 | T | G | intronic | NLRP3 | . | | . | 0 | 0 | 0 | 0 | 0.4059 | . | . | . | . | . |
| 1 | 247597211 | 247597211 | T | C | intronic | NLRP3 | . | | . | 0 | 0 | 0 | 0 | 0.4 | . | . | . | . | . |
| 1 | 247597329 | 247597329 | C | T | intronic | NLRP3 | . | | . | 0 | 0 | 0.917732 | 0 | 0.04545 | . | . | . | . | . |
| 1 | 247597365 | 247597365 | G | A | intronic | NLRP3 | . | | . | 0.907427 | 0 | 0.788738 | 0 | 0.05682 | . | . | . | . | . |
| 1 | 247599139 | 247599139 | G | C | intronic | NLRP3 | . | | . | 0 | 0 | 0.0421326 | 0 | 0.02841 | . | . | . | . | . |
| 1 | 247599232 | 247599232 | G | A | intronic | NLRP3 | . | | . | 0.496848 | 0.586 | 0.484026 | 0.5922 | 0.4716 | . | . | . | . | . |
| 1 | 247599513 | 247599513 | T | C | intronic | NLRP3 | . | | . | 0 | 0 | 0.0489217 | 0 | 0.02841 | . | . | . | . | . |
| 1 | 247600301 | 247600301 | A | C | intronic | NLRP3 | . | | . | 0 | 0 | 0.654353 | 0 | 0.4261 | . | . | . | . | . |
| 1 | 247608167 | 247608167 | C | A | intronic | NLRP3 | . | | . | 0.134861 | 0.1416 | 0.0972444 | 0.141 | 0.1648 | . | . | . | . | . |
| 2 | 113887207 | 113887207 | T | C | exonic | IL1RN | . | Benign | Osteomyelitis,_sterile_multifocal,_with_periostitis_and_pustulosis\|not_specified | 0.206136 | 0.2491 | 0.191693 | 0.2574 | 0.1791 | . | . | . | synonymous SNV | IL1RN:NM_173842:exon2:c.T171C:p.A57A,IL1RN:NM_000577:exon3:c.T117C:p.A39A,IL1RN:NM_173841:exon4:c.T180C:p.A60A,IL1RN:NM_173843:exon4:c.T69C:p.A23A,IL1RN:NM_001318914:exon5:c.T69C:p.A23A |

| Chr | Start | PVS1 | PS1 | PS2 | PS3 | PS4 | PM1 | PM2 | PM3 | PM4 | PM5 | PM6 | PP1 | PP2 | PP3 | PP4 | PP5 | BA1 | BS1 | BS2 | BS3 | BS4 | BP1 | BP2 | BP3 | BP4 | BP5 | BP6 | BP7 | Final Annotation | avsnp147 | ACMG classification | Other information |
|---|---|---|---|---|---|---|---|---|---|---|---|---|---|---|---|---|---|---|---|---|---|---|---|---|---|---|---|---|---|---|---|---|---|
| 5 | 138856982 | | | | | | PM1 | | | | | | | PP2 | PP3 | | | BA1 | | BS2 | | | | | | | | | | PM1,PP2,PP3,BA1,BS2 | rs7380824 | Benign (I) | |
| 5 | 138861078 | | | | | | PM1 | | | | | | | PP2 | | | | BA1 | | BS2 | | | | | | BP4 | | | | PM1,PP2,BA1,BS2,BP4 | rs11554776 | Benign (I) | |
| 6 | 32811629 | | | | | | | | | | | | | PP2 | | | | BA1 | | BS2 | | | | | | BP4 | | BP6 | | PP2,BA1,BS2,BP4,BP6 | rs2071543 | Benign (I) | |
| 6 | 32825090 | | | | | | PM1 | | | | | | | PP2 | | | | BA1 | | BS2 | | | | | | BP4 | | | | PM1,PP2,BA1,BS2,BP4 | rs17587 | Benign (I) | |
| 6 | 138196066 | | | | | | PM1 | | | | | | | | | | | BA1 | | BS2 | | | | | | BP4 | | | | PM1,BA1,BS2,BP4 | rs2230926 | Benign (I) | |
| 11 | 117864063 | | | | | | | | | | | | | PP2 | | | | BA1 | | BS2 | | | | | | BP4 | | BP6 | | PP2,BA1,BS2,BP4,BP6 | rs3135932 | Benign (I) | |
| 11 | 117869670 | | | | | | | | | | | | | PP2 | | | | BA1 | | BS2 | | | | | | BP4 | | BP6 | | PP2,BA1,BS2,BP4,BP6 | rs2229113 | Benign (I) | |
| 12 | 110013879 | | | | | | | | | | | | | | | | | BA1 | | BS2 | | | | | | BP4 | | BP6 | | BA1,BS2,BP4,BP6 | rs7957619 | Benign (I) | |
| 16 | 50744624 | | | | | | | | | | | | | PP2 | | | | BA1 | | BS2 | | | | | | BP4 | | BP6 | | PP2,BA1,BS2,BP4,BP6 | rs2066842 | Benign (I) | |
| 16 | 50757276 | | | | | | | | | | | | | PP2 | | | | BA1 | BS1 | BS2 | | | | | | BP4 | | BP6 | | PP2,BA1,BS1,BS2,BP4,BP6 | rs5743291 | Benign (I) | |
| 17 | 78171944 | | | | | | | | | | | | | | | | | BA1 | | BS2 | | | | | | BP4 | | BP6 | | BA1,BS2,BP4,BP6 | rs2066964 | Benign (I) | |
| 17 | 78178893 | | | | | | | | | | | | | | PP3 | | | BA1 | | BS2 | | | | | | | | BP6 | | PP3,BA1,BS2,BP6 | rs11652075 | Benign (I) | |
| 19 | 55450727 | | | | | | | | | | | | | | | | | BA1 | | BS2 | | | | | | BP4 | | BP6 | | BA1,BS2,BP4,BP6 | rs775881 | Benign (I) | |
| 19 | 55450746 | | | | | | | | | | | | | | | | | BA1 | | BS2 | | | | | | BP4 | | BP6 | | BA1,BS2,BP4,BP6 | rs61747414 | Benign (I) | |
| 1 | 247582063 | | | | | | | | | | | | | | | | | | BS1 | BS2 | | | | | | | | BP6 | | BS1,BS2,BP6 | rs73136263 | Benign (II) | |
| 1 | 247582309 | | | | | | PM1 | | | | | | | | | | | | | | | | | | | | | BP6 | BP7 | PM1,BP6,BP7 | rs200082602 | Likely Benign (II) | |
| 1 | 247586336 | | | | | | | | | | | | | | | | | BA1 | | BS2 | | | | | | | | | | BA1,BS2 | rs3806265 | Benign (I) | |
| 1 | 247586920 | | | | | | | | | | | | | | | | | BA1 | | BS2 | | | | | | | | | | BA1,BS2 | rs4925651 | Benign (I) | |
| 1 | 247587174 | | | | | | PM1 | | | | | | | | | | | | | | | | | | | | | BP6 | BP7 | PM1,BP6,BP7 | rs56710146 | Likely Benign (II) | |
| 1 | 247587408 | | | | | | PM1 | | | | | | | | | | | BA1 | | BS2 | | | | | | | | BP6 | BP7 | PM1,BA1,BS2,BP6,BP7 | rs7525979 | Benign (I) | |
| 1 | 247587477 | | | | | | PM1 | | | | | | | | | | | BA1 | | BS2 | | | | | | | | BP6 | BP7 | PM1,BA1,BS2,BP6,BP7 | rs3806268 | Benign (I) | |
| 1 | 247587681 | | | | | | PM1 | | | | | | | | | | | | | | | | | | | | | BP6 | BP7 | PM1,BP6,BP7 | rs143840033 | Likely Benign (II) | |
| 1 | 247587771 | | | | | | PM1 | | | | | | | | | | | | | | | | | | | | | BP6 | BP7 | PM1,BP6,BP7 | rs41311573 | Likely Benign (II) | |
| 1 | 247587783 | | | | | | PM1 | | | | | | | | | | | | | | | | | | | | | BP6 | BP7 | PM1,BP6,BP7 | rs180177471 | Likely Benign (II) | |
| 1 | 247587795 | | | | | | PM1 | | | | | | | | | | | | BS1 | | | | | | | | | BP6 | BP7 | PM1,BS1,BP6,BP7 | rs143140947 | Likely Benign (I) | |
| 1 | 247587982 | | | | | | | | | | | | | | | | | | | | | | | | | | | BP6 | BP7 | BP6,BP7 | rs148478875 | Likely Benign (II) | |
| 1 | 247587996 | | | | | | | | | | | | | | | | | | | | | | | | | | | BP6 | BP7 | BP6,BP7 | rs139852370 | Likely Benign (II) | |
| 1 | 247588053 | | | | | | | | | | | | | | | | | BA1 | | BS2 | | | | | | | | BP6 | BP7 | BA1,BS2,BP6,BP7 | rs34298354 | Benign (I) | |
| 1 | 247588351 | | | | | | | | | | | | | | | | | | | | | | | | | | | BP6 | BP7 | BP6,BP7 | rs116054301 | Likely Benign (II) | |
| 1 | 247588960 | | | | | | | | | | | | | | | | | BA1 | | BS2 | | | | | | | | | | BA1,BS2 | rs3738447 | Benign (I) | |
| 1 | 247593142 | | | | | | | | | | | | | | | | | BA1 | | BS2 | | | | | | | | | | BA1,BS2 | rs12060377 | Benign (I) | |
| 1 | 247597180 | | | | | | | PM2 | | | | | | | | | | | | BS2 | | | | | | | | | | PM2,BS2 | rs76911796 | VUS | |
| 1 | 247597181 | | | | | | | PM2 | | | | | | | | | | | | BS2 | | | | | | | | | | PM2,BS2 | rs148560736 | VUS | |
| 1 | 247597208 | | | | | | | PM2 | | | | | | | | | | | | BS2 | | | | | | | | | | PM2,BS2 | rs113966299 | VUS | |
| 1 | 247597211 | | | | | | | PM2 | | | | | | | | | | | | BS2 | | | | | | | | | | PM2,BS2 | rs112695411 | VUS | |
| 1 | 247597329 | | | | | | | | | | | | | | | | | BA1 | | BS2 | | | | | | | | | | BA1,BS2 | rs12124866 | Benign (I) | |
| 1 | 247597365 | | | | | | | | | | | | | | | | | BA1 | | BS2 | | | | | | | | | | BA1,BS2 | rs10925022 | Benign (I) | |
| 1 | 247599139 | | | | | | | | | | | | | | | | | | BS1 | BS2 | | | | | | | | | | BS1,BS2 | rs34777555 | Benign (II) | |
| 1 | 247599232 | | | | | | | | | | | | | | | | | BA1 | | BS2 | | | | | | | | | | BA1,BS2 | rs10754557 | Benign (I) | |
| 1 | 247599513 | | | | | | | | | | | | | | | | | | BS1 | BS2 | | | | | | | | | | BS1,BS2 | rs76225363 | Benign (II) | |
| 1 | 247600301 | | | | | | | | | | | | | | | | | BA1 | | BS2 | | | | | | | | | | BA1,BS2 | rs1539019 | Benign (I) | |
| 1 | 247608167 | | | | | | | | | | | | | | | | | BA1 | | BS2 | | | | | | | | | | BA1,BS2 | rs45624634 | Benign (I) | |
| 2 | 113887207 | | | | | | | | | | | | | | | | | BA1 | | BS2 | | | | | | | | BP6 | BP7 | BA1,BS2,BP6,BP7 | rs419598 | Benign (I) | |

<table>
<tr><td>2</td><td>113890304</td><td>113890304</td><td>T</td><td>C</td><td>exonic</td><td>IL1RN</td><td>.</td><td>Benign</td><td>Osteomyelitis,_sterile_multifocal,_with_periostitis_and_pustulosis|not_specified</td><td>0.333154</td><td>0.3082</td><td>0.35004</td><td>0.3044</td><td>0.198</td><td>.</td><td>.</td><td>.</td><td>synonymous SNV</td><td>IL1RN:NM_173842:exon4:c.T390C:p.S130S,IL1RN:NM_000577:exon5:c.T336C:p.S112S,IL1RN:NM_173841:exon6:c.T399C:p.S133S,IL1RN:NM_173843:exon6:c.T288C:p.S96S,IL1RN:NM_001318914:exon7:c.T288C:p.S96S</td><td></td><td></td><td></td><td></td><td></td><td>PM1</td><td></td><td></td><td></td><td></td><td></td><td></td><td></td><td></td><td></td><td></td><td>BA1</td><td></td><td>BS2</td><td></td><td></td><td></td><td></td><td></td><td></td><td></td><td>BP6</td><td>BP7</td><td>PM1,BA1,BS2,BP6,BP7</td><td>rs315952</td><td>Benign (I)</td><td></td></tr>
<tr><td>2</td><td>113890586</td><td>113890586</td><td>C</td><td>G</td><td>UTR3</td><td>IL1RN</td><td>NM_173841:c.*138C>G; NM_001318914:c.*138C>G; NM_173843:c.*138C>G; NM_000577:c.*138C>G; NM_173842:c.*138C>G</td><td>Benign</td><td>Osteomyelitis,_sterile_multifocal,_with_periostitis_and_pustulosis</td><td>0</td><td>0</td><td>0.355032</td><td>0</td><td>0.2102</td><td>.</td><td>.</td><td>.</td><td>.</td><td>.</td><td></td><td></td><td></td><td></td><td></td><td></td><td></td><td></td><td></td><td></td><td></td><td></td><td></td><td></td><td></td><td></td><td>BA1</td><td></td><td>BS2</td><td></td><td></td><td></td><td></td><td></td><td></td><td></td><td>BP6</td><td></td><td>BA1,BS2,BP6</td><td>rs315951</td><td>Benign (I)</td><td></td></tr>
<tr><td>2</td><td>113890610</td><td>113890610</td><td>C</td><td>T</td><td>UTR3</td><td>IL1RN</td><td>NM_173841:c.*162C>T; NM_001318914:c.*162C>T; NM_173843:c.*162C>T; NM_000577:c.*162C>T; NM_173842:c.*162C>T</td><td>Likely_benign</td><td>Osteomyelitis,_sterile_multifocal,_with_periostitis_and_pustulosis</td><td>0</td><td>0</td><td>0.01877</td><td>0</td><td>0.1705</td><td>.</td><td>.</td><td>.</td><td>.</td><td>.</td><td></td><td></td><td></td><td></td><td></td><td></td><td></td><td></td><td></td><td></td><td></td><td></td><td></td><td></td><td></td><td></td><td></td><td>BS1</td><td>BS2</td><td></td><td></td><td></td><td></td><td></td><td></td><td></td><td>BP6</td><td></td><td>BS1,BS2,BP6</td><td>rs4252041</td><td>Benign (II)</td><td></td></tr>
<tr><td>12</td><td>110012766</td><td>110012766</td><td>A</td><td>G</td><td>intronic</td><td>MVK</td><td>.</td><td>Benign</td><td>.</td><td>0.121989</td><td>0</td><td>0.142173</td><td>0</td><td>0.1023</td><td>.</td><td>.</td><td>.</td><td>.</td><td>.</td><td></td><td></td><td></td><td></td><td></td><td></td><td></td><td></td><td></td><td></td><td></td><td></td><td></td><td></td><td></td><td></td><td>BA1</td><td></td><td>BS2</td><td></td><td></td><td></td><td></td><td></td><td></td><td></td><td>BP6</td><td></td><td>BA1,BS2,BP6</td><td>rs61940512</td><td>Benign (I)</td><td></td></tr>
<tr><td>12</td><td>110017759</td><td>110017759</td><td>C</td><td>T</td><td>intronic</td><td>MVK</td><td>.</td><td>Benign/Likely_benign</td><td>not_specified|Hyperimmunoglobulin_D_with_periodic_fever|Mevalonic_aciduria</td><td>0.058435</td><td>0.0605</td><td>0.0267572</td><td>0.0589</td><td>0.07955</td><td>.</td><td>.</td><td>.</td><td>.</td><td>.</td><td></td><td></td><td></td><td></td><td></td><td>PM1</td><td></td><td></td><td></td><td></td><td></td><td></td><td></td><td></td><td></td><td></td><td>BA1</td><td>BS1</td><td>BS2</td><td></td><td></td><td></td><td></td><td></td><td></td><td></td><td>BP6</td><td></td><td>PM1,BA1,BS1,BS2,BP6</td><td>rs67886029</td><td>Benign (I)</td><td></td></tr>
<tr><td>12</td><td>110019233</td><td>110019233</td><td>G</td><td>A</td><td>exonic</td><td>MVK</td><td>.</td><td>Benign/Likely_benign</td><td>not_specified|Hyperimmunoglobulin_D_with_periodic_fever|Mevalonic_aciduria</td><td>0.051515</td><td>0.047</td><td>0.0321486</td><td>0.0453</td><td>0.05867</td><td>.</td><td>.</td><td>.</td><td>synonymous SNV</td><td>MVK:NM_000431:exon5:c.G405A:p.S135S,MVK:NM_001114185:exon5:c.G405A:p.S135S</td><td></td><td></td><td></td><td></td><td></td><td>PM1</td><td></td><td></td><td></td><td></td><td></td><td></td><td></td><td></td><td></td><td></td><td>BA1</td><td>BS1</td><td>BS2</td><td></td><td></td><td></td><td></td><td></td><td></td><td></td><td>BP6</td><td>BP7</td><td>PM1,BA1,BS1,BS2,BP6,BP7</td><td>rs34368092</td><td>Benign (I)</td><td></td></tr>
<tr><td>12</td><td>110019338</td><td>110019338</td><td>C</td><td>T</td><td>exonic</td><td>MVK</td><td>.</td><td>Benign/Likely_benign</td><td>not_specified|Hyperimmunoglobulin_D_with_periodic_fever|Mevalonic_aciduria</td><td>0.17761</td><td>0.1618</td><td>0.163339</td><td>0.1605</td><td>0.2323</td><td>.</td><td>.</td><td>.</td><td>synonymous SNV</td><td>MVK:NM_000431:exon5:c.C510T:p.D170D,MVK:NM_001114185:exon5:c.C510T:p.D170D</td><td></td><td></td><td></td><td></td><td></td><td>PM1</td><td></td><td></td><td></td><td></td><td></td><td></td><td></td><td></td><td></td><td></td><td>BA1</td><td></td><td>BS2</td><td></td><td></td><td></td><td></td><td></td><td></td><td></td><td>BP6</td><td>BP7</td><td>PM1,BA1,BS2,BP6,BP7</td><td>rs2287218</td><td>Benign (I)</td><td></td></tr>
<tr><td>12</td><td>110024541</td><td>110024541</td><td>A</td><td>G</td><td>intronic</td><td>MVK</td><td>.</td><td>Benign</td><td>not_specified</td><td>0.271951</td><td>0.2339</td><td>0.228634</td><td>0.2293</td><td>0.3068</td><td>.</td><td>.</td><td>.</td><td>.</td><td>.</td><td></td><td></td><td></td><td></td><td></td><td>PM1</td><td></td><td></td><td></td><td></td><td></td><td></td><td></td><td></td><td></td><td></td><td>BA1</td><td></td><td>BS2</td><td></td><td></td><td></td><td></td><td></td><td></td><td></td><td>BP6</td><td></td><td>PM1,BA1,BS2,BP6</td><td>rs2270375</td><td>Benign (I)</td><td></td></tr>
<tr><td>12</td><td>110028689</td><td>110028689</td><td>G</td><td>C</td><td>intronic</td><td>MVK</td><td>.</td><td>not_provided</td><td>Hyperimmunoglobulin_D_with_periodic_fever</td><td>0.003229</td><td>0.0011</td><td>0.00539137</td><td>0.0008</td><td>0.005682</td><td>.</td><td>.</td><td>.</td><td>.</td><td>.</td><td></td><td></td><td></td><td></td><td></td><td></td><td></td><td></td><td></td><td></td><td></td><td></td><td></td><td></td><td></td><td></td><td></td><td></td><td></td><td></td><td></td><td></td><td></td><td></td><td></td><td></td><td></td><td></td><td></td><td>rs104895347</td><td>VUS</td><td></td></tr>
<tr><td>12</td><td>110029008</td><td>110029008</td><td>C</td><td>T</td><td>intronic</td><td>MVK</td><td>.</td><td>Benign</td><td>.</td><td>0.172305</td><td>0.1601</td><td>0.154752</td><td>0.1593</td><td>0.2159</td><td>.</td><td>.</td><td>.</td><td>.</td><td>.</td><td></td><td></td><td></td><td></td><td></td><td></td><td></td><td></td><td></td><td></td><td></td><td></td><td></td><td></td><td></td><td></td><td>BA1</td><td></td><td>BS2</td><td></td><td></td><td></td><td></td><td></td><td></td><td></td><td>BP6</td><td></td><td>BA1,BS2,BP6</td><td>rs35191208</td><td>Benign (I)</td><td></td></tr>
<tr><td>12</td><td>110029039</td><td>110029039</td><td>T</td><td>G</td><td>intronic</td><td>MVK</td><td>.</td><td>Benign/Likely_benign</td><td>Hyperimmunoglobulin_D_with_periodic_fever|not_provided</td><td>0.00346</td><td>0.0065</td><td>0.00678914</td><td>0.0066</td><td>0.01136</td><td>.</td><td>.</td><td>.</td><td>.</td><td>.</td><td></td><td></td><td></td><td></td><td></td><td></td><td></td><td></td><td></td><td></td><td></td><td></td><td></td><td></td><td></td><td></td><td></td><td></td><td>BS2</td><td></td><td></td><td></td><td></td><td></td><td></td><td></td><td>BP6</td><td></td><td>BS2,BP6</td><td>rs104895331</td><td>Likely Benign (I)</td><td></td></tr>
<tr><td>12</td><td>110029057</td><td>110029057</td><td>C</td><td>A</td><td>exonic</td><td>MVK</td><td>.</td><td>Conflicting_interpretations_of_pathogenicity</td><td>Hyperimmunoglobulin_D_with_periodic_fever|Mevalonic_aciduria</td><td>0.010457</td><td>0.0025</td><td>0.00738818</td><td>0.002</td><td>0.00249</td><td>.</td><td>.</td><td>.</td><td>synonymous SNV</td><td>MVK:NM_001301182:exon8:c.C624A:p.I208I,MVK:NM_000431:exon9:c.C780A:p.I260I,MVK:NM_001114185:exon9:c.C780A:p.I260I</td><td></td><td></td><td></td><td></td><td></td><td></td><td></td><td></td><td></td><td></td><td></td><td></td><td></td><td></td><td></td><td></td><td></td><td>BS1</td><td></td><td></td><td></td><td></td><td></td><td></td><td></td><td></td><td></td><td>BP7</td><td>BS1,BP7</td><td>rs34975996</td><td>Likely Benign (I)</td><td></td></tr>
<tr><td>12</td><td>110029108</td><td>110029108</td><td>C</td><td>T</td><td>exonic</td><td>MVK</td><td>.</td><td>Benign/Likely_benign</td><td>Hyperimmunoglobulin_D_with_periodic_fever|not_specified|Mevalonic_aciduria</td><td>0.010149</td><td>0.0058</td><td>0.0169728</td><td>0.0053</td><td>0.003188</td><td>.</td><td>.</td><td>.</td><td>synonymous SNV</td><td>MVK:NM_001301182:exon8:c.C675T:p.R225R,MVK:NM_000431:exon9:c.C831T:p.R277R,MVK:NM_001114185:exon9:c.C831T:p.R277R</td><td></td><td></td><td></td><td></td><td></td><td></td><td></td><td></td><td></td><td></td><td></td><td></td><td></td><td></td><td></td><td></td><td></td><td>BS1</td><td></td><td></td><td></td><td></td><td></td><td></td><td></td><td></td><td>BP6</td><td>BP7</td><td>BS1,BP6,BP7</td><td>rs104895353</td><td>Likely Benign (I)</td><td></td></tr>
<tr><td>12</td><td>110029186</td><td>110029186</td><td>G</td><td>A</td><td>intronic</td><td>MVK</td><td>.</td><td>Benign</td><td>.</td><td>0.128864</td><td>0.149</td><td>0.111422</td><td>0.1496</td><td>0.1761</td><td>.</td><td>.</td><td>.</td><td>.</td><td>.</td><td></td><td></td><td></td><td></td><td></td><td>PM1</td><td></td><td></td><td></td><td></td><td></td><td></td><td></td><td></td><td></td><td></td><td>BA1</td><td></td><td>BS2</td><td></td><td></td><td></td><td></td><td></td><td></td><td></td><td>BP6</td><td></td><td>PM1,BA1,BS2,BP6</td><td>rs2270374</td><td>Benign (I)</td><td></td></tr>
<tr><td>12</td><td>110029279</td><td>110029279</td><td>C</td><td>A</td><td>intronic</td><td>MVK</td><td>.</td><td></td><td>.</td><td>0</td><td>0</td><td>0.0267572</td><td>0</td><td>0.07955</td><td>.</td><td>.</td><td>.</td><td>.</td><td>.</td><td></td><td></td><td></td><td></td><td></td><td>PM1</td><td></td><td></td><td></td><td></td><td></td><td></td><td></td><td></td><td></td><td></td><td></td><td>BS1</td><td>BS2</td><td></td><td></td><td></td><td></td><td></td><td></td><td></td><td></td><td></td><td>PM1,BS1,BS2</td><td>rs72648031</td><td>Benign (II)</td><td></td></tr>
<tr><td>15</td><td>77310636</td><td>77310636</td><td>G</td><td>C</td><td>intronic</td><td>PSTPIP1</td><td>.</td><td></td><td>.</td><td>0.730242</td><td>0.7481</td><td>0.60603</td><td>0.7212</td><td>0.2273</td><td>.</td><td>.</td><td>.</td><td>.</td><td>.</td><td></td><td></td><td></td><td></td><td></td><td>PM1</td><td></td><td></td><td></td><td></td><td></td><td></td><td></td><td></td><td></td><td></td><td>BA1</td><td></td><td>BS2</td><td></td><td></td><td></td><td></td><td></td><td></td><td></td><td></td><td></td><td>PM1,BA1,BS2</td><td>rs3812911</td><td>Benign (I)</td><td></td></tr>
<tr><td>15</td><td>77325279</td><td>77325279</td><td>C</td><td>T</td><td>exonic</td><td>PSTPIP1</td><td>.</td><td>Benign</td><td>not_specified|Pyogenic_arthritis,_pyoderma_gangrenosum_and_acne</td><td>0.03735</td><td>0.0495</td><td>0.057508</td><td>0.0281</td><td>0.02273</td><td>.</td><td>.</td><td>.</td><td>synonymous SNV</td><td>PSTPIP1:NM_003978:exon12:c.C915T:p.C305C,PSTPIP1:NM_001321136:exon13:c.C888T:p.C296C,PSTPIP1:NM_001321137:exon13:c.C1110T:p.C370C</td><td></td><td></td><td></td><td></td><td></td><td></td><td></td><td></td><td></td><td></td><td></td><td></td><td></td><td></td><td></td><td></td><td>BA1</td><td>BS1</td><td>BS2</td><td></td><td></td><td></td><td></td><td></td><td></td><td></td><td>BP6</td><td>BP7</td><td>BA1,BS1,BS2,BP6,BP7</td><td>rs11858480</td><td>Benign (I)</td><td></td></tr>
<tr><td>16</td><td>50731102</td><td>50731102</td><td>C</td><td>T</td><td>UTR5</td><td>NOD2</td><td>NM_022162:c.-53C>T</td><td>Likely_benign</td><td>Crohn_disease|Blau_syndrome</td><td>0.000438</td><td>0</td><td>0.000199681</td><td>0</td><td>0.005682</td><td>.</td><td>.</td><td>.</td><td>.</td><td>.</td><td></td><td></td><td></td><td></td><td></td><td></td><td></td><td></td><td></td><td></td><td></td><td></td><td></td><td></td><td></td><td></td><td></td><td></td><td></td><td></td><td></td><td></td><td></td><td></td><td></td><td></td><td>BP6</td><td></td><td>BP6</td><td>rs188341692</td><td>VUS</td><td></td></tr>
<tr><td>16</td><td>50733374</td><td>50733374</td><td>G</td><td>T</td><td>UTR5</td><td>NOD2</td><td>NM_001293557:c.-33G>T</td><td></td><td>.</td><td>0.211977</td><td>0.1982</td><td>0.115615</td><td>0.2018</td><td>0.1932</td><td>.</td><td>.</td><td>.</td><td>.</td><td>.</td><td></td><td></td><td></td><td></td><td></td><td></td><td></td><td></td><td></td><td></td><td></td><td></td><td></td><td></td><td></td><td></td><td>BA1</td><td></td><td>BS2</td><td></td><td></td><td></td><td></td><td></td><td></td><td></td><td></td><td></td><td>BA1,BS2</td><td>rs2076753</td><td>Benign (I)</td><td></td></tr>
<tr><td>16</td><td>50733392</td><td>50733392</td><td>T</td><td>A</td><td>UTR5</td><td>NOD2</td><td>NM_001293557:c.-15T>A</td><td>Conflicting_interpretations_of_pathogenicity</td><td>Sarcoidosis,_early-onset|not_specified|Blau_syndrome|Crohn_disease</td><td>0.001308</td><td>0.0018</td><td>0.000599042</td><td>0.0023</td><td>0.01705</td><td>.</td><td>.</td><td>.</td><td>.</td><td>.</td><td></td><td></td><td></td><td></td><td></td><td></td><td></td><td></td><td></td><td></td><td></td><td></td><td></td><td></td><td></td><td></td><td></td><td></td><td>BS2</td><td></td><td></td><td></td><td></td><td></td><td></td><td></td><td></td><td></td><td>BS2</td><td>rs104895421</td><td>VUS</td><td></td></tr>
<tr><td>16</td><td>50733859</td><td>50733859</td><td>C</td><td>G</td><td>exonic</td><td>NOD2</td><td>.</td><td>Benign</td><td>Blau_syndrome|Crohn_disease|not_specified</td><td>0.370883</td><td>0.336</td><td>0.246605</td><td>0.332</td><td>0.38</td><td>.</td><td>.</td><td>.</td><td>synonymous SNV</td><td>NOD2:NM_001293557:exon1:c.C453G:p.S151S,NOD2:NM_022162:exon2:c.C534G:p.S178S</td><td></td><td></td><td></td><td></td><td></td><td></td><td></td><td></td><td></td><td></td><td></td><td></td><td></td><td></td><td></td><td></td><td>BA1</td><td></td><td>BS2</td><td></td><td></td><td></td><td></td><td></td><td></td><td></td><td>BP6</td><td>BP7</td><td>BA1,BS2,BP6,BP7</td><td>rs2067085</td><td>Benign (I)</td><td></td></tr>
<tr><td>16</td><td>50745199</td><td>50745199</td><td>C</td><td>T</td><td>exonic</td><td>NOD2</td><td>.</td><td>Benign</td><td>Blau_syndrome|Crohn_disease|not_specified</td><td>0.201939</td><td>0.1864</td><td>0.107628</td><td>0.1906</td><td>0.193</td><td>.</td><td>.</td><td>.</td><td>synonymous SNV</td><td>NOD2:NM_001293557:exon3:c.C1296T:p.R432R,NOD2:NM_022162:exon4:c.C1377T:p.R459R</td><td></td><td></td><td></td><td></td><td></td><td>PM1</td><td></td><td></td><td></td><td></td><td></td><td></td><td></td><td></td><td></td><td></td><td>BA1</td><td></td><td>BS2</td><td></td><td></td><td></td><td></td><td></td><td></td><td></td><td>BP6</td><td>BP7</td><td>PM1,BA1,BS2,BP6,BP7</td><td>rs2066843</td><td>Benign (I)</td><td></td></tr>
<tr><td>16</td><td>50745583</td><td>50745583</td><td>T</td><td>G</td><td>exonic</td><td>NOD2</td><td>.</td><td>Benign</td><td>Blau_syndrome|Crohn_disease|not_specified</td><td>0.335796</td><td>0.3302</td><td>0.216054</td><td>0.3303</td><td>0.3649</td><td>.</td><td>.</td><td>.</td><td>synonymous SNV</td><td>NOD2:NM_001293557:exon3:c.T1680G:p.R560R,NOD2:NM_022162:exon4:c.T1761G:p.R587R</td><td></td><td></td><td></td><td></td><td></td><td></td><td></td><td></td><td></td><td></td><td></td><td></td><td></td><td></td><td></td><td></td><td>BA1</td><td></td><td>BS2</td><td></td><td></td><td></td><td></td><td></td><td></td><td></td><td>BP6</td><td>BP7</td><td>BA1,BS2,BP6,BP7</td><td>rs1861759</td><td>Benign (I)</td><td></td></tr>
<tr><td>16</td><td>50745610</td><td>50745610</td><td>G</td><td>A</td><td>exonic</td><td>NOD2</td><td>.</td><td>Benign</td><td>Sarcoidosis,_early-onset</td><td>0.000846</td><td>0.0004</td><td>0.000599042</td><td>0.0004</td><td>0.001063</td><td>.</td><td>.</td><td>.</td><td>synonymous SNV</td><td>NOD2:NM_001293557:exon3:c.G1707A:p.T569T,NOD2:NM_022162:exon4:c.G1788A:p.T596T</td><td></td><td></td><td></td><td></td><td></td><td></td><td></td><td></td><td></td><td></td><td></td><td></td><td></td><td></td><td></td><td></td><td></td><td></td><td></td><td></td><td></td><td></td><td></td><td></td><td></td><td></td><td>BP6</td><td>BP7</td><td>BP6,BP7</td><td>rs104895437</td><td>Likely Benign (II)</td><td></td></tr>
<tr><td>16</td><td>50745655</td><td>50745655</td><td>C</td><td>T</td><td>exonic</td><td>NOD2</td><td>.</td><td>Benign/Likely_benign</td><td>Crohn_disease|Blau_syndrome</td><td>0.007541</td><td>0.0098</td><td>0.00259585</td><td>0.0098</td><td>0.004782</td><td>.</td><td>.</td><td>.</td><td>synonymous SNV</td><td>NOD2:NM_001293557:exon3:c.C1752T:p.A584A,NOD2:NM_022162:exon4:c.C1833T:p.A611A</td><td></td><td></td><td></td><td></td><td></td><td></td><td></td><td></td><td></td><td></td><td></td><td></td><td></td><td></td><td></td><td></td><td></td><td></td><td></td><td></td><td></td><td></td><td></td><td></td><td></td><td></td><td>BP6</td><td>BP7</td><td>BP6,BP7</td><td>rs61736932</td><td>Likely Benign (II)</td><td></td></tr>
<tr><td>16</td><td>50750791</td><td>50750791</td><td>G</td><td>T</td><td>intronic</td><td>NOD2</td><td>.</td><td>Likely_benign</td><td>Crohn_disease|Blau_syndrome</td><td>0.003924</td><td>0.0012</td><td>0.00559105</td><td>0.0009</td><td>0.005682</td><td>.</td><td>.</td><td>.</td><td>.</td><td>.</td><td></td><td></td><td></td><td></td><td></td><td></td><td></td><td></td><td></td><td></td><td></td><td></td><td></td><td></td><td></td><td></td><td></td><td></td><td></td><td></td><td></td><td></td><td></td><td></td><td></td><td></td><td>BP6</td><td></td><td>BP6</td><td>rs5743285</td><td>VUS</td><td></td></tr>
<tr><td>16</td><td>50756774</td><td>50756774</td><td>C</td><td>T</td><td>intronic</td><td>NOD2</td><td>.</td><td>Pathogenic,_risk_factor</td><td>INFLAMMATORY_BOWEL_DISEASE_1_(CROHN_DISEASE),_SUSCEPTIBILITY_TO|Blau_syndrome|YAO_SYNDROME,_SUSCEPTIBILITY_TO</td><td>0</td><td>0</td><td>0.0517173</td><td>0</td><td>0.04545</td><td>.</td><td>.</td><td>.</td><td>.</td><td>.</td><td></td><td></td><td></td><td></td><td></td><td>PM1</td><td></td><td></td><td></td><td></td><td></td><td></td><td></td><td></td><td></td><td>PP5</td><td>BA1</td><td></td><td>BS2</td><td></td><td></td><td></td><td></td><td></td><td></td><td></td><td></td><td></td><td>PM1,PP5,BA1,BS2</td><td>rs5743289</td><td>Benign (I)</td><td></td></tr>
<tr><td>16</td><td>50766603</td><td>50766603</td><td>C</td><td>T</td><td>UTR3</td><td>NOD2</td><td>NM_022162:c.*873C>T; NM_001293557:c.*873C>T</td><td>Uncertain_significance</td><td>Crohn_disease|Blau_syndrome</td><td>0</td><td>0</td><td>0.00119808</td><td>0</td><td>0.07386</td><td>.</td><td>.</td><td>.</td><td>.</td><td>.</td><td></td><td></td><td></td><td></td><td></td><td></td><td></td><td></td><td></td><td></td><td></td><td></td><td></td><td></td><td></td><td></td><td></td><td></td><td>BS2</td><td></td><td></td><td></td><td></td><td></td><td></td><td></td><td></td><td></td><td>BS2</td><td>rs373812846</td><td>VUS</td><td></td></tr>
<tr><td>17</td><td>78178830</td><td>78178830</td><td>A</td><td>G</td><td>intronic</td><td>CARD14</td><td>.</td><td>Benign</td><td>not_specified</td><td>0.410148</td><td>0.4904</td><td>0.358626</td><td>0.4125</td><td>0.3182</td><td>.</td><td>.</td><td>.</td><td>.</td><td>.</td><td></td><td></td><td></td><td></td><td></td><td></td><td></td><td></td><td></td><td></td><td></td><td></td><td></td><td></td><td></td><td></td><td>BA1</td><td></td><td>BS2</td><td></td><td></td><td></td><td></td><td></td><td></td><td></td><td>BP6</td><td></td><td>BA1,BS2,BP6</td><td>rs11653893</td><td>Benign (I)</td><td></td></tr>
<tr><td>19</td><td>54313707</td><td>54313707</td><td>G</td><td>C</td><td>exonic</td><td>NLRP12</td><td>.</td><td>Benign</td><td>not_specified|Familial_cold_autoinflammatory_syndrome</td><td>0.05113</td><td>0.052</td><td>0.0269569</td><td>0.051</td><td>0.01642</td><td>T</td><td>D</td><td>23.1</td><td>nonsynonymous SNV</td><td>NLRP12:NM_001277126:exon3:c.C1206G:p.F402L,NLRP12:NM_001277129:exon3:c.C1206G:p.F402L,NLRP12:NM_144687:exon3:c.C1206G:p.F402L</td><td></td><td></td><td></td><td></td><td></td><td></td><td></td><td></td><td></td><td></td><td></td><td></td><td></td><td>PP3</td><td></td><td></td><td>BA1</td><td>BS1</td><td>BS2</td><td></td><td></td><td></td><td></td><td></td><td></td><td></td><td>BP6</td><td></td><td>PP3,BA1,BS1,BS2,BP6</td><td>rs34971363</td><td>Benign (I)</td><td></td></tr>
<tr><td>1</td><td>247582310</td><td>247582310</td><td>G</td><td>A</td><td>exonic</td><td>NLRP3</td><td>.</td><td>Benign/Likely_benign</td><td>not_specified|Chronic_infantile_neurological,_cutaneous_and_articular_syndrome|Familial_amyloid_nephropathy_with_urticaria_AND_deafness|Familial_cold_autoinflammatory_syndrome</td><td>0</td><td>0.0006</td><td>0.00219649</td><td>0.0007</td><td>0.000586</td><td>D</td><td>P</td><td>18.61</td><td>nonsynonymous SNV</td><td>NLRP3:NM_001127462:exon1:c.G214A:p.V72M,NLRP3:NM_001243133:exon1:c.G208A:p.V70M,NLRP3:NM_004895:exon1:c.G214A:p.V72M,NLRP3:NM_183395:exon1:c.G214A:p.V72M,NLRP3:NM_001127461:exon2:c.G214A:p.V72M,NLRP3:NM_001079821:exon3:c.G214A:p.V72M</td><td></td><td></td><td></td><td></td><td></td><td>PM1</td><td></td><td></td><td></td><td></td><td></td><td></td><td>PP2</td><td>PP3</td><td></td><td></td><td></td><td></td><td></td><td></td><td></td><td></td><td></td><td></td><td></td><td></td><td>BP6</td><td></td><td>PM1,PP2,PP3,BP6</td><td>rs117287351</td><td>VUS</td><td></td></tr>
<tr><td>1</td><td>247582326</td><td>247582326</td><td>C</td><td>T</td><td>exonic</td><td>NLRP3</td><td>.</td><td>Uncertain_significance</td><td>not_specified</td><td>0</td><td>6.67E-05</td><td>0</td><td>0.0001</td><td>0.01046</td><td>D</td><td>B</td><td>23.9</td><td>nonsynonymous SNV</td><td>NLRP3:NM_001127462:exon1:c.C230T:p.A77V,NLRP3:NM_001243133:exon1:c.C224T:p.A75V,NLRP3:NM_004895:exon1:c.C230T:p.A77V,NLRP3:NM_183395:exon1:c.C230T:p.A77V,NLRP3:NM_001127461:exon2:c.C230T:p.A77V,NLRP3:NM_001079821:exon3:c.C230T:p.A77V</td><td></td><td></td><td></td><td></td><td></td><td>PM1</td><td></td><td></td><td></td><td></td><td></td><td></td><td>PP2</td><td>PP3</td><td></td><td></td><td></td><td></td><td>BS2</td><td></td><td></td><td></td><td></td><td></td><td></td><td></td><td></td><td></td><td>PM1,PP2,PP3,BS2</td><td>rs200288250</td><td>VUS</td><td></td></tr>
<tr><td>1</td><td>247587343</td><td>247587343</td><td>G</td><td>A</td><td>exonic</td><td>NLRP3</td><td>.</td><td>Conflicting_interpretations_of_pathogenicity</td><td>Familial_cold_urticaria|not_provided|not_specified|Familial_cold_autoinflammatory_syndrome</td><td>0.006843</td><td>0.0083</td><td>0.00399361</td><td>0.0085</td><td>0.005453</td><td>T</td><td>B</td><td>0.002</td><td>nonsynonymous SNV</td><td>NLRP3:NM_001127462:exon3:c.G598A:p.V200M,NLRP3:NM_001243133:exon3:c.G592A:p.V198M,NLRP3:NM_004895:exon3:c.G598A:p.V200M,NLRP3:NM_183395:exon3:c.G598A:p.V200M,NLRP3:NM_001127461:exon4:c.G598A:p.V200M,NLRP3:NM_001079821:exon5:c.G598A:p.V200M</td><td></td><td></td><td></td><td></td><td></td><td>PM1</td><td></td><td></td><td></td><td></td><td></td><td>PP1</td><td>PP2</td><td></td><td>PP4</td><td></td><td></td><td></td><td></td><td></td><td></td><td></td><td></td><td></td><td>BP4</td><td></td><td></td><td></td><td>PM1,PP1,PP2,PP4,BP4</td><td>rs121908147</td><td>VUS</td><td>PubMed: 49161, PubMed: 11687797: PP1-M, : In a family with familial cold autoinflammatory syndrome (FCAS1; 120100) previously reported by Vlagopoulos et al. (1975), Hoffman et al. (2001) identified a G-to-A transition at nucleotide 592 in exon 3 of the CIAS1 gene, resulting in a valine-to-methionine substitution at codon 198 (V198M). PMCID: PMC4321998 We investigated the allele frequency for one of the earliest reported CIAS1 mutations, V198M, in a large collection of DNA samples from Caucasian subjects. This mutation was originally identified in an FCAS family that was later found to also have an E525K mutation (5), and was subsequently reported in Spanish (42) and French (4) FCAS families, in a British family with MWS and 2 British patients with unchQ(ARA)cterized periodic fevers (3), and in 3 German patients with atypical periodic fevers (43). V198M has been considered to be a reduced-penetrance CIAS1 mutation because it was found in asymptomatic family members and in control DNA samples. PMID:24986970, PMCID: PMC3672677 PP4 pmid24123366: Related to Qatari individual</td></tr>
<tr><td>1</td><td>247587688</td><td>247587688</td><td>A</td><td>G</td><td>exonic</td><td>NLRP3</td><td>.</td><td>Uncertain_significance</td><td>Familial_cold_urticaria</td><td>0.000231</td><td>0.0002</td><td>0</td><td>0.0002</td><td>0.001172</td><td>T</td><td>B</td><td>0.001</td><td>nonsynonymous SNV</td><td>NLRP3:NM_001127462:exon3:c.A943G:p.I315V,NLRP3:NM_001243133:exon3:c.A937G:p.I313V,NLRP3:NM_004895:exon3:c.A943G:p.I315V,NLRP3:NM_183395:exon3:c.A943G:p.I315V,NLRP3:NM_001127461:exon4:c.A943G:p.I315V,NLRP3:NM_001079821:exon5:c.A943G:p.I315V</td><td></td><td></td><td></td><td></td><td></td><td>PM1</td><td></td><td></td><td></td><td></td><td></td><td>PP1-M</td><td>PP2</td><td></td><td></td><td></td><td></td><td></td><td></td><td></td><td></td><td></td><td></td><td></td><td>BP4</td><td></td><td></td><td></td><td>PM1,PP1-M,PP2,BP4</td><td>rs180177501</td><td>VUS</td><td>PMC3194675:PP1-M:Both case #1 (M.T) and #2 (V.C) displayed a mild clinical phenotype (episodes of urticarial rash and arthralgia associated with elevation of acute phase reactants), compatible with FCAS and Muckle-Wells syndrome, respectively. Both patients displayed good response to NSAID and/or steroid on demand.</td></tr>
<tr><td>1</td><td>247588858</td><td>247588858</td><td>C</td><td>A</td><td>exonic</td><td>NLRP3</td><td>.</td><td>Conflicting_interpretations_of_pathogenicity</td><td>not_specified|Familial_cold_autoinflammatory_syndrome|Chronic_infantile_neurological,_cutaneous_and_articular_syndrome|Familial_amyloid_nephropathy_with_urticaria_AND_deafness|not_provided</td><td>0.03483</td><td>0.0409</td><td>0.0223642</td><td>0.0394</td><td>0.04027</td><td>T</td><td>B</td><td>0.01</td><td>nonsynonymous SNV</td><td>NLRP3:NM_001127462:exon3:c.C2113A:p.Q705K,NLRP3:NM_001243133:exon3:c.C2107A:p.Q703K,NLRP3:NM_004895:exon3:c.C2113A:p.Q705K,NLRP3:NM_183395:exon3:c.C2113A:p.Q705K,NLRP3:NM_001127461:exon4:c.C2113A:p.Q705K,NLRP3:NM_001079821:exon5:c.C2113A:p.Q705K</td><td></td><td></td><td></td><td>PS3</td><td>PS4</td><td></td><td></td><td></td><td></td><td></td><td></td><td></td><td>PP2</td><td></td><td></td><td></td><td></td><td>BS1</td><td>BS2</td><td></td><td></td><td></td><td></td><td></td><td>BP4</td><td></td><td></td><td></td><td>PS3,PS4,PP2,BS1,BS2,BP4</td><td>rs35829419</td><td>VUS - confliciting evidence</td><td>PS3:PMID:22529966: NLRP3-WT and NLRP3-Q705K were retrovirally transduced into the human monocytic cell line THP-1, followed by the assessment of IL-1β and IL-18 levels in the cell culture supernatant. hese findings show that the Q705K polymorphism in NLRP3 is a gain-of-function alteration leading to an overactive NLRP3 inflammasome. PMID:27819323, PMID:22128899 No association.This study suggests that variations in the innate immunity genes CARD8 (p.C10X) and NLRP3 (p.Q705K) have no effect on RA susceptibility either in the Tunisian or in the French population. PS4: PMID:26535712 Very high odds ratio PMID: 26273672 We observed an association between NLRP3 rs35829419 polymorphism and the risk for macrovascular complications (OR = 3.93; 95% CI = 1.54–10.0; P = 0.004). rs35829419 with macrovascular complications was observed only in males (OR = 4.44; 95% CI = 1.39–14.25; P = 0.012) (Table 2). Additionally, significant association with PAOD was observed among males (OR = 5.93; 95% CI = 1.17–30.06; P = 0.032). This association could not be tested in females as only 3 female patients had PAOD and all were NLRP3 CC homozygous. PMID:24142982 In stratified analysis, carriage of at least one NLRP3 rs35829419 variant allele conferred a 2.5-fold increased risk for pathological interstitial lung fibrosis (OR 2.44, 95% CI 0.97 to 6.14), although this association was only borderline significant.</td></tr>
<tr><td>1</td><td>247607973</td><td>247607973</td><td>C</td><td>T</td><td>exonic</td><td>NLRP3</td><td>.</td><td>Conflicting_interpretations_of_pathogenicity</td><td>not_specified|Familial_cold_autoinflammatory_syndrome|Familial_amyloid_nephropathy_with_urticaria_AND_deafness|Chronic_infantile_neurological,_cutaneous_and_articular_syndrome</td><td>0.000692</td><td>0.0012</td><td>0.00179712</td><td>0.0013</td><td>0.002787</td><td>D</td><td>D</td><td>33</td><td>nonsynonymous SNV</td><td>NLRP3:NM_183395:exon6:c.C2519T:p.T840M,NLRP3:NM_001127462:exon7:c.C2690T:p.T897M,NLRP3:NM_001127461:exon8:c.C2690T:p.T897M,NLRP3:NM_001243133:exon8:c.C2855T:p.T952M,NLRP3:NM_004895:exon8:c.C2861T:p.T954M,NLRP3:NM_001079821:exon10:c.C2861T:p.T954M</td><td></td><td></td><td></td><td></td><td></td><td></td><td></td><td></td><td></td><td></td><td></td><td></td><td>PP2</td><td>PP3</td><td></td><td></td><td></td><td></td><td></td><td></td><td></td><td></td><td></td><td></td><td></td><td></td><td></td><td></td><td>PP2,PP3</td><td>rs139814109</td><td>VUS</td><td></td></tr>
<tr><td>2</td><td>113819812</td><td>113819812</td><td>C</td><td>T</td><td>exonic</td><td>IL36RN</td><td>.</td><td></td><td>.</td><td>0.000154</td><td>0.0003</td><td>0.000798722</td><td>0.0003</td><td>0.001758</td><td>T</td><td>D</td><td>26.3</td><td>nonsynonymous SNV</td><td>IL36RN:NM_012275:exon4:c.C227T:p.P76L,IL36RN:NM_173170:exon4:c.C227T:p.P76L</td><td></td><td></td><td></td><td></td><td></td><td>PM1</td><td></td><td></td><td></td><td></td><td></td><td></td><td>PP2</td><td>PP3</td><td></td><td></td><td></td><td></td><td></td><td></td><td></td><td></td><td></td><td></td><td></td><td></td><td></td><td></td><td>PM1,PP2,PP3</td><td>rs139497891</td><td>VUS</td><td>PMID: 25468355</td></tr>
<tr><td>2</td><td>113890284</td><td>113890284</td><td>G</td><td>A</td><td>exonic</td><td>IL1RN</td><td>.</td><td>Benign</td><td>.</td><td>0.000923</td><td>0.003</td><td>0.00479233</td><td>0.003</td><td>0.01791</td><td>T</td><td>B</td><td>4.647</td><td>nonsynonymous SNV</td><td>IL1RN:NM_173842:exon4:c.G370A:p.A124T,IL1RN:NM_000577:exon5:c.G316A:p.A106T,IL1RN:NM_173841:exon6:c.G379A:p.A127T,IL1RN:NM_173843:exon6:c.G268A:p.A90T,IL1RN:NM_001318914:exon7:c.G268A:p.A90T</td><td></td><td></td><td></td><td></td><td></td><td>PM1</td><td></td><td></td><td></td><td></td><td></td><td></td><td></td><td></td><td></td><td></td><td></td><td></td><td>BS2</td><td></td><td></td><td>BP1</td><td></td><td></td><td>BP4</td><td></td><td>BP6</td><td></td><td>PM1,BS2,BP1,BP4,BP6</td><td>rs45507693</td><td>Likely Benign (I)</td><td></td></tr>
<tr><td>2</td><td>224642493</td><td>224642493</td><td>G</td><td>A</td><td>exonic</td><td>AP1S3</td><td>.</td><td>Likely_benign</td><td>Psoriasis_15,_pustular,_susceptibility_to|not_specified</td><td>0.008593</td><td>0.008</td><td>0.00239617</td><td>0.0075</td><td>0.001636</td><td>D</td><td>D</td><td>34</td><td>nonsynonymous SNV</td><td>AP1S3:NM_001039569:exon2:c.C97T:p.R33W</td><td></td><td></td><td></td><td>PS3</td><td></td><td>PM1</td><td></td><td></td><td></td><td></td><td></td><td></td><td></td><td>PP3</td><td></td><td></td><td></td><td></td><td></td><td></td><td>BS4</td><td></td><td></td><td></td><td></td><td></td><td>BP6</td><td></td><td>PS3,PM1,PP3,BS4,BP6</td><td>rs138292988</td><td>VUS</td><td>PubMed: 24791904: PS3 Transfection studies in HEK293 cells and HaCaT keratinocytes showed decreased levels of F4C mutant protein compared to wildtype. The c.11T-G allele frequency was significantly higher in affected individuals (2.3%) than in 1,695 unrelated controls (0.6%; p = 0.01). BS4 Segregation analysis in a representative pedigree revealed that the proband had inherited the c.11T-G disease allele from an unaffected parent.</td></tr>
<tr><td>2</td><td>224642579</td><td>224642579</td><td>A</td><td>C</td><td>exonic</td><td>AP1S3</td><td>.</td><td>risk_factor</td><td>Psoriasis_15,_pustular,_susceptibility_to</td><td>0.009587</td><td>0.0078</td><td>0.00299521</td><td>0.0077</td><td>0.005682</td><td>D</td><td>D</td><td>27.2</td><td>nonsynonymous SNV</td><td>AP1S3:NM_001039569:exon2:c.T11G:p.F4C</td><td></td><td></td><td></td><td>PS3</td><td></td><td>PM1</td><td></td><td></td><td></td><td></td><td></td><td></td><td></td><td>PP3</td><td></td><td></td><td></td><td></td><td></td><td></td><td>BS4</td><td></td><td></td><td></td><td></td><td></td><td></td><td></td><td>PS3,PM1,PP3,BS4</td><td>rs116107386</td><td>Likely pathogenic (II)</td><td>PubMed: 24791904: PS3 Transfection studies in HEK293 cells and HaCaT keratinocytes showed decreased levels of F4C mutant protein compared to wildtype. The c.11T-G allele frequency was significantly higher in affected individuals (2.3%) than in 1,695 unrelated controls (0.6%; p = 0.01). BS4 Segregation analysis in a representative pedigree revealed that the proband had inherited the c.11T-G disease allele from an unaffected parent.</td></tr>
<tr><td>4</td><td>2828992</td><td>2828992</td><td>C</td><td>T</td><td>exonic</td><td>SH3BP2</td><td>.</td><td>Benign/Likely_benign</td><td>Fibrous_dysplasia_of_jaw</td><td>0.005613</td><td>0.0056</td><td>0.00399361</td><td>0.0053</td><td>0.01692</td><td>D</td><td>B</td><td>22.6</td><td>nonsynonymous SNV</td><td>SH3BP2:NM_001122681:exon6:c.C464T:p.A155V,SH3BP2:NM_001145855:exon6:c.C548T:p.A183V,SH3BP2:NM_001145856:exon6:c.C635T:p.A212V,SH3BP2:NM_003023:exon6:c.C464T:p.A155V</td><td></td><td></td><td></td><td></td><td></td><td></td><td></td><td></td><td></td><td></td><td></td><td></td><td>PP2</td><td>PP3</td><td></td><td></td><td></td><td></td><td>BS2</td><td></td><td></td><td></td><td></td><td></td><td></td><td></td><td>BP6</td><td></td><td>PP2,PP3,BS2,BP6</td><td>rs35313240</td><td>Likely Benign (I)</td><td></td></tr>
<tr><td>6</td><td>32823948</td><td>32823948</td><td>G</td><td>A</td><td>exonic</td><td>PSMB9</td><td>.</td><td></td><td>.</td><td>0.028436</td><td>0.0156</td><td>0.0217652</td><td>0.0161</td><td>0.007996</td><td>D</td><td>P</td><td>23.2</td><td>nonsynonymous SNV</td><td>PSMB9:NM_002800:exon2:c.G94A:p.V32I</td><td></td><td></td><td></td><td></td><td></td><td>PM1</td><td></td><td></td><td></td><td></td><td></td><td></td><td>PP2</td><td>PP3</td><td></td><td></td><td></td><td>BS1</td><td></td><td></td><td></td><td></td><td></td><td></td><td></td><td></td><td></td><td></td><td>PM1,PP2,PP3,BS1</td><td>rs241419</td><td>VUS</td><td></td></tr>
</table>

| | | | | | | | | | | | | | | | | | | | | | | | | | | | | | | | | | | | | | | | | | | | | | | | | | | | |
|---|---|---|---|---|---|---|---|---|---|---|---|---|---|---|---|---|---|---|---|---|---|---|---|---|---|---|---|---|---|---|---|---|---|---|---|---|---|---|---|---|---|---|---|---|---|---|---|---|---|---|---|
| 6 | 138196060 | 138196060 | C | T | exonic | TNFAIP3 | . | Uncertain_significance | not_specified | 0.005536 | 0.0017 | 0.00599042 | 0.0012 | 0.001594 | D | B | 15.11 | nonsynonymous SNV | TNFAIP3:NM_001270507:exon3:c.C374T:p.A125V,TNFAIP3:NM_001270508:exon3:c.C374T:p.A125V,TNFAIP3:NM_006290:exon3:c.C374T:p.c | | | | PS3 | | PM1 | | | | | | | | PP3 | | | | | | | | | | | | | | | PS3,PM1,PP3 | rs5029941 | Likely pathogenic (II) | PubMed: 20483768:PS3:Lodolce et al. (2010) noted that there are 2 nonsynonymous coding polymorphisms in the deubiquitinating (DUB) domain of TNFAIP3: phe127 to cys (F127C ; rs2230926), which is in high linkage disequilibrium with SLE risk variants in people of European descent, and ala125 to val (A125V; rs5029941). They performed a case-control study with 217 African-American SLE patients using these coding variants, along with 6 tagging SNPs in TNFAIP3, and identified a novel African-derived risk haplotype. Although F127C was associated with SLE risk in this African American population, the most significant SLE-associated risk variant was an African-derived tagging SNP, rs5029953 (p = 0.008; odds ratio = 1.6). The rare minor allele of A125V was associated with protection from SLE in African Americans (p = 0.05; odds ratio = 0.38) and with susceptibility to inflammatory bowel disease (IBD; 266600) (p = 0.027; odds ratio = 3.73). Enzymatic functional analysis showed that A125V was associated with diminished TNFAIP3 deubiquitinating activity, and computer modeling suggested that A125V altered the 3-dimensional structure of the DUB domain to a greater extent than F127C. PMID: 24489017 |
| 10 | 72358215 | 72358215 | A | C | exonic | PRF1 | . | | . | 0.000308 | 0.0004 | 0.000199681 | 0.0003 | 0.001639 | T | B | 0.005 | nonsynonymous SNV | PRF1:NM_001083116:exon3:c.T1262G:p.F421C,PRF1:NM_005041:exon3:c.T1262G:p.F421C | | | | | | PM1 | | | | | | | | | | | | | | BS3 | | | | | BP4 | | | | PM1,BS3,BP4 | rs200534908 | Likely Benign (I) | PMID 20536554:BS3:Consistent with the above hypothesis, two mutations (the common polymorphism Ala91Val and Arg232His), complemented GME(AP)proximately 50% of wildtype PRF function in mouse CTLs, while the F421C mutation had essentially normal activity compared with wildtype PRF. However, none of the other cancer-associated mutants had any detectable activity in either RBL cells or primary CTLs |
| 10 | 72358722 | 72358722 | T | C | exonic | PRF1 | . | Conflicting_interpretations_of_pathogenicity | Hemophagocytic_lymphohistiocytosis,_familial,_2\|Malignant_lymphoma,_non-Hodgkin\|not_specified | 0.008458 | 0.0052 | 0.00758786 | 0.005 | 0.001758 | T | B | 0.037 | nonsynonymous SNV | PRF1:NM_001083116:exon3:c.A755G:p.N252S,PRF1:NM_005041:exon3:c.A755G:p.N252S | | | | | PS4 | PM1 | | | | | | | | | | | | | | BS3 | | | | | BP4 | | | | PS4,PM1,BS3,BP4 | rs28933375 | VUS | PubMed: 10583959: In a patient with familial hemophagocytic lymphohistiocytosis (FHL2; 603553), Stepp et al. (1999) identified a 755A-G transition in the PRF1 gene, resulting in an asn252-to-ser (N252S) substitution. PP1 PubMed: 15459303, PubMed: 15659737 In a 27-year-old man with autoimmune lymphoproliferative syndrome (601859) who later developed a T-cell-rich, histiocyte-rich, diffuse large B-cell lymphoma (605027), Clementi et al. (2004) identified a heterozygous N252S mutation in the PRF1 gene and a heterozygous mutation in the FAS gene (134637). The FAS mutation was inherited from his healthy father and was also carried by his healthy brother, whereas the PRF1 mutation was inherited from his healthy mother. Clementi et al. (2004) noted that the N252S substitution occurs within the membrane attack complex of the perforin protein, a region involved in the pore-forming activity of the molecule. However, both the patient and his mother had normal levels of perforin and normal NK cell activity. The authors suggested that the combined effect of the 2 mutant genes contributed to the development of ALPS and lymphoma in this patient. Rieux-Laucat et al. (2005) cast doubt on the pathogenicity of the N252S mutation, and the primary authors provided a rebuttal (Clementi et al., 2005). PubMed: 15728124 Clementi et al. (2005) identified the N252S mutation in 1 of 660 (0.02%) control alleles, suggesting that it is a benign polymorphism. PMID: 16720836:PS4:N252S conferred an OR = 62.7 (P = .0016) for ALPS 18198357:PS4 :In both cohorts, allelic frequency of N252S was significantly higher in patients than in control subjects (combined cohorts: 1.5 vs. 0.4%; odds ratio 6.68 [95% CI 1.83–7.48]). PMC4184267 :PP4 One patient died of MAS: she carried the N252S PRF1 variant and showed reduced perforin expression. PMID 15755897:BS3 In contrast, expression and function of N252S-substituted perforin were normal.Taken together, our data and epidemiologic studies9 indicate that the N252S substitution alone could not have been causative of disease but, rather, that an additional genetic defect(s) might have been responsible. We therefore concluded that N252S probably represents a neutral PRF1 polymorphism. note: THIS VARIANT IS RESPONSIBLE FOR CAUSING DISEASE IN COMPOUND HETEROZYGOUS STATE ONLY. OTHERWISE IT IS BENIGN POLYMORPHISM. |
| 10 | 72358845 | 72358845 | G | A | exonic | PRF1 | . | Benign | . | 0 | 0.0024 | 0.00299521 | 0.0022 | 0.002657 | T | B | 0.122 | nonsynonymous SNV | PRF1:NM_001083116:exon3:c.C632T:p.A211V,PRF1:NM_005041:exon3:c.C632T:p.A211V | | | | | | PM1 | | | | | | PP1 | | | | | | | | BS3 | BS4 | | | | BP4 | | BP6 | | PM1,PP1,BS3,BS4,BP4,BP6 | rs368524364 | Benign (II) | PMID: 21936944 BS3: Cr-release cytotoxicity assays were performed using p.Ala211Val perforin transfected RBL cells. The p. Ala211Val substitution showed normal WT perforin and lytic activity (Figure 2).BS4 and PP1: second cousin sister has mutation and affected by disease but mother and other members of family had mutation but they are free of disease |
| 10 | 72360291 | 72360291 | C | T | exonic | PRF1 | . | | . | 0.000923 | 0.0005 | 0.000199681 | 0.0005 | 0.003529 | D | B | 18.12 | nonsynonymous SNV | PRF1:NM_001083116:exon2:c.G368A:p.R123H,PRF1:NM_005041:exon2:c.G368A:p.R123H | | | | | | | | | | | | | | PP3 | | | | | | | | | | | | | | | PP3 | rs139336186 | VUS | |
| 10 | 72360387 | 72360387 | G | A | exonic | PRF1 | . | Conflicting_interpretations_of_pathogenicity | Hemophagocytic_lymphohistiocytosis,_familial,_2,_susceptibility_to\|not_provided\|Familial_hemophagocytic_lymphohistiocytosis\|not_specified | 0.033754 | 0.0311 | 0.0131789 | 0.0294 | 0.005034 | D | D | 26 | nonsynonymous SNV | PRF1:NM_001083116:exon2:c.C272T:p.A91V,PRF1:NM_005041:exon2:c.C272T:p.A91V | | | | | | | | | | | | | | PP3 | | | | BS1 | | | | | | | | | | | PP3,BS1 | rs35947132 | VUS | |
| 10 | 72360577 | 72360577 | G | A | exonic | PRF1 | . | | . | 0.00054 | 0.0013 | 0.000199681 | 0.0012 | 0.001779 | D | P | 21.1 | nonsynonymous SNV | PRF1:NM_001083116:exon2:c.C82T:p.R28C,PRF1:NM_005041:exon2:c.C82T:p.R28C | | | | PS3 | | | | | | | | | | PP3 | | | | | | | | | | | | | | | PS3,PP3 | rs141660796 | VUS | PMID:20019066:PS3:Repeated flow cytometry for perforin in cytotoxic lymphocytes and assessment of lytic NK cell function in Patient P54 showed intermediately decreased levels of perforin and lytic NK cell function (~20%).PMID: 23734337: Both PRF1A91V- and PRF1R28C-expressing lymphocytes exhibited severely impaired but measurable cytotoxic function. |
| 10 | 72360648 | 72360648 | C | T | exonic | PRF1 | . | Conflicting_interpretations_of_pathogenicity | Aplastic_anemia\|not_specified\|Familial_hemophagocytic_lymphohistiocytosis\|not_provided | 0.026793 | 0.0116 | 0.0219649 | 0.0059 | 0.01705 | D | B | 19.06 | nonsynonymous SNV | PRF1:NM_001083116:exon2:c.G11A:p.R4H,PRF1:NM_005041:exon2:c.G11A:p.R4H | | | | | | | | | | | | | | PP3 | | | | BS1 | BS2 | | | | | | | | | | PP3,BS1,BS2 | rs35418374 | Benign (II) | |
| 12 | 6442643 | 6442643 | C | T | exonic | TNFRSF1A | . | Conflicting_interpretations_of_pathogenicity | TNF_receptor-associated_periodic_fever_syndrome_(TRAPS)\|not_specified\|not_provided | 0.01384 | 0.0141 | 0.00599042 | 0.0132 | 0.009815 | T | B | 15.3 | nonsynonymous SNV | TNFRSF1A:NM_001346091:exon3:c.G38A:p.R13Q,TNFRSF1A:NM_001065:exon4:c.G362A:p.R121Q | | | | | | PM1 | | | | | | | PP2 | | | | | BS1 | | | | | | | BP4 | | | | PM1,PP2,BS1,BP4 | rs4149584 | Likely Benign (I) | |
| 12 | 6443001 | 6443001 | G | A | exonic | TNFRSF1A | . | Benign/Likely_benign | not_specified\|Familial_Periodic_Fever | 0.025373 | 0.0127 | 0.028155 | 0.0055 | 0.01361 | D | D | 22.2 | nonsynonymous SNV | TNFRSF1A:NM_001065:exon3:c.C224T:p.P75L | | | | | | PM1 | | | | | | | PP2 | PP3 | | | | BS1 | BS2 | | | | | | | | BP6 | | PM1,PP2,PP3,BS1,BS2,BP6 | rs4149637 | Benign (II) | |
| 12 | 110017618 | 110017618 | G | A | exonic | MVK | . | Conflicting_interpretations_of_pathogenicity | Hyperimmunoglobulin_D_with_periodic_fever\|not_specified | 0.002537 | 0.0007 | 0.00199681 | 0.0007 | 0.001063 | T | B | 0.002 | nonsynonymous SNV | MVK:NM_000431:exon4:c.G238A:p.V80I,MVK:NM_001114185:exon4:c.G238A:p.V80I,MVK:NM_001301182:exon4:c.G238A:p.V80I | | | | | | | | | | | | | | | | | | | | | | | | | BP4 | | | | BP4 | rs76914224 | VUS | |
| 12 | 110019222 | 110019222 | G | A | exonic | MVK | . | not_provided | Hyperimmunoglobulin_D_with_periodic_fever | 0 | 2.47E-05 | 0 | 3.66E-05 | 0.001794 | T | B | 0.003 | nonsynonymous SNV | MVK:NM_000431:exon5:c.G394A:p.V132I,MVK:NM_001114185:exon5:c.G394A:p.V132I | | | | | | PM1 | PM2 | | | | | | | | | | | | | | | | | | BP4 | | | | PM1,PM2,BP4 | rs104895336 | VUS | PMID:15536479, PMID:25897835, PMID:19877056, PMID26935981 |
| 12 | 110034320 | 110034320 | G | A | exonic | MVK | . | Pathogenic | Hyperimmunoglobulin_D_with_periodic_fever\|Hyperimmunoglobulin_D_with_periodic_fever\|Mevalonic_aciduria\|not_provided | 0.001692 | 0.0014 | 0 | 0.0016 | 0.003212 | T | B | 20.3 | nonsynonymous SNV | MVK:NM_001301182:exon10:c.G973A:p.V325I,MVK:NM_000431:exon11:c.G1129A:p.V377I,MVK:NM_001114185:exon11:c.G1129A:p.V377I | | | | | PS4 | | | PM3 | | | | PP1 | | | | PP5 | | | | | | | | | BP4 | | | | PS4,PM3,PP1,PP5,BP4 | rs28934897 | Likely pathogenic (II) | PS4 :PubMed: 10369261: In 4 patients with the hyper-IgD syndrome (HIDS; 260920), Houten et al. (1999) found a 1129G-A transition, resulting in a val377-to-ile amino acid substitution. In 3 of the 4 patients, V377I was present in compound heterozygous state; in the fourth patient, V377I GME(AP)peared to be present in homozygous state. The V377I mutation was found in homozygous state in a family with HIDS studied by Drenth et al. (1999).PS4 PubMed: 11313768 In 16 unrelated patients with HIDS, Houten et al. (2001) detected the V377I mutation, including one sibship with homozygosity. PS4 PubMed: 11313769 Cuisset et al. (2001) detected the V377I mutation in 20 of 25 unrelated patients with HIDS, the majority of whom were of Dutch origin. PS4 PubMed: 15536479 D'Osualdo et al. (2005) identified the V377I mutation in 50% of mutant alleles in 15 unrelated Italian patients with HIDS. In 2 patients the V377I mutation was found in homozygous state. PMID: 24470648 : PP1, PM3 Mutation analysis of the MVK gene revealed that the girl was compound heterozygote for 2 mutations, c.803T>C (p.Ile268Thr) and c.1129G>A (p.Val377Ile), known to be associated with the HIDS phenotype. Genotyping of both parents revealed that both parents carried a heterozygous mutation of the MVK gene: mother, c.803T>C (p.Ile268Thr); father, c.1129G>A (p.Val377Ile).(As it is taken from mother and father then it is in trans). PMID: 23979089 PP1, PM3 MVK enzyme activity in lymphocytes was below 1% and genomic sequencing of the MVK gene revealed 2 compound heterozygous mutations: p.[Val377Ile]+[Tyr116His]. The patient's mother carried the heterozygous p.Tyr116His mutation, whereas the father had heterozygous p.Val377Ile mutation. |
| 15 | 55520906 | 55520906 | G | A | exonic | RAB27A | . | Likely_pathogenic | not_provided | 0 | 8.30E-06 | 0 | 1.63E-05 | 0.005845 | D | D | 34 | nonsynonymous SNV | RAB27A:NM_004580:exon4:c.C244T:p.R82C,RAB27A:NM_183234:exon5:c.C244T:p.R82C,RAB27A:NM_183235:exon5:c.C244T:p.R82C,RAB27A:NM_183236:exon5:c.C244T:p.R82C | | | | PS3 | | PM1 | PM2 | | | | | PP1 | | PP3 | | PP5 | | | | | | | | | | | | | PS3,PM1,PM2,PP1,PP3,PP5 | rs753966933 | Pathogenic III(b) | **PMID:27016801 PS3: Rab27a R82C bound melanophilin, although to a lesser extent than wild-type Rab27a (Fig 2, A). In contrast, no Munc13-4 was detectable in the Rab27a R82C precipitate; PP1: Both homozygous patients (II.4 and II.5) have normal NK cell numbers but reduced CD107a mobilization, which could explain the severely decreased cytotoxic NK cell function. The heterozygous siblings (II.3 and II.6) are less affected. The different cytotoxic responses observed for II.3 and II.6 could be caused by different percentages of NK cells detected for these patient. PMID:29357941 in chinese patient** |
| 15 | 77324670 | 77324670 | G | C | exonic | PSTPIP1 | . | Benign/Likely_benign | not_specified\|Pyogenic_arthritis,_pyoderma_gangrenosum_and_acne | 0.011028 | 0.0078 | 0.0147764 | 0.0081 | 0.05114 | T | P | 3.648 | nonsynonymous SNV | PSTPIP1:NM_001321135:exon11:c.G773C:p.G258A,PSTPIP1:NM_003978:exon11:c.G773C:p.G258A,PSTPIP1:NM_001321136:exon12:c.G746C:p.G249A,PSTPIP1:NM_001321137:exon12:c.G968C:p.G323A | | | | | | | | | | | | | | | | | | BS1 | BS2 | | | | | | BP4 | | BP6 | | BS1,BS2,BP4,BP6 | rs34240327 | Benign (II) | |
| 16 | 50733743 | 50733743 | G | A | exonic | NOD2 | . | Benign | . | 0.007156 | 0.002 | 0.00698882 | 0.0017 | 0.005682 | T | B | 14.02 | nonsynonymous SNV | NOD2:NM_001293557:exon1:c.G337A:p.A113T,NOD2:NM_022162:exon2:c.G418A:p.A140T | | | | | | | | | | | | | PP2 | | | | | | | | | | | | BP4 | | BP6 | | PP2,BP4,BP6 | rs34684955 | Likely Benign (II) | |
| 16 | 50744688 | 50744688 | A | G | exonic | NOD2 | . | Benign/Likely_benign | Crohn_disease | 0.00554 | 0.0043 | 0.00399361 | 0.0044 | 0.001172 | T | B | 0.026 | nonsynonymous SNV | NOD2:NM_001293557:exon3:c.A785G:p.N262S,NOD2:NM_022162:exon4:c.A866G:p.N289S | | | | | | | | | | | | | PP2 | | | | | | | | | | | | BP4 | | BP6 | | PP2,BP4,BP6 | rs5743271 | Likely Benign (II) | |
| 16 | 50744877 | 50744877 | A | G | exonic | NOD2 | . | Benign | . | 0.003386 | 0.0011 | 0.00279553 | 0.0009 | 0.002345 | D | B | 15.93 | nonsynonymous SNV | NOD2:NM_001293557:exon3:c.A974G:p.H325R,NOD2:NM_022162:exon4:c.A1055G:p.H352R | | | | | | PM1 | | | | | | | PP2 | PP3 | | | | | | | | | | | | | BP6 | | PM1,PP2,PP3,BP6 | rs5743272 | VUS | |
| 16 | 50744939 | 50744939 | C | T | exonic | NOD2 | . | not_provided | Sarcoidosis,_early-onset | 0.000154 | 0.0002 | 0 | 0.0002 | 0.001758 | T | B | 16.54 | nonsynonymous SNV | NOD2:NM_001293557:exon3:c.C1036T:p.R346C,NOD2:NM_022162:exon4:c.C1117T:p.R373C | | | | | | PM1 | | | | | | | PP2 | | | | | | | | | | | | BP4 | | | | PM1,PP2,BP4 | rs145293873 | VUS | |
| 16 | 50745868 | 50745868 | G | T | exonic | NOD2 | . | Likely_benign | Crohn_disease\|Blau_syndrome | 0.000154 | 0.0005 | 0.000199681 | 0.0004 | 0.002822 | D | D | 24.4 | nonsynonymous SNV | NOD2:NM_001293557:exon3:c.G1965T:p.L655F,NOD2:NM_022162:exon4:c.G2046T:p.L682F | | | | | | | | | | | | | PP2 | PP3 | | | | | | | | | | | | | BP6 | | PP2,PP3,BP6 | rs149002807 | VUS | |
| 16 | 50745872 | 50745872 | C | T | exonic | NOD2 | . | Conflicting_interpretations_of_pathogenicity | Crohn_disease\|Blau_syndrome | 0.000385 | 0.0004 | 0.000199681 | 0.0004 | 0.005682 | D | B | 23.2 | nonsynonymous SNV | NOD2:NM_001293557:exon3:c.C1969T:p.R657W,NOD2:NM_022162:exon4:c.C2050T:p.R684W | | | | | | | | | | | | | PP2 | PP3 | | | | | | | | | | | | | | | PP2,PP3 | rs5743276 | VUS | |
| 16 | 50745873 | 50745873 | G | A | exonic | NOD2 | . | | . | 0.000924 | 0.0003 | 0.00259585 | 0.0003 | 0.003612 | T | B | 7.749 | nonsynonymous SNV | NOD2:NM_001293557:exon3:c.G1970A:p.R657Q,NOD2:NM_022162:exon4:c.G2051A:p.R684Q | | | | | | | | | | | | | PP2 | | | | | | | | | | | | BP4 | | | | PP2,BP4 | rs114664276 | VUS | |
| 16 | 50745926 | 50745926 | C | T | exonic | NOD2 | . | Conflicting_interpretations_of_pathogenicity,_risk_factor | INFLAMMATORY_BOWEL_DISEASE_1_(CROHN_DISEASE),_SUSCEPTIBILITY_TO\|not_specified\|Crohn_disease\|Blau_syndrome\|YAO_SYNDROME,_SUSCEPTIBILITY_TO | 0.031558 | 0.0227 | 0.014377 | 0.0259 | 0.002934 | D | P | 24.1 | nonsynonymous SNV | NOD2:NM_001293557:exon3:c.C2023T:p.R675W,NOD2:NM_022162:exon4:c.C2104T:p.R702W | | | | | | | | | | | | | PP2 | PP3 | | | | BS1 | | | | | | | | | | | PP2,PP3,BS1 | rs2066844 | VUS | |
| 16 | 50745996 | 50745996 | C | G | exonic | NOD2 | . | Benign | Crohn_disease\|Blau_syndrome | 0.012927 | 0.0041 | 0.0183706 | 0.0031 | 0.00763 | T | B | 3.629 | nonsynonymous SNV | NOD2:NM_001293557:exon3:c.C2093G:p.A698G,NOD2:NM_022162:exon4:c.C2174G:p.A725G | | | | | | | | | | | | | PP2 | | | | | BS1 | | | | | | | BP4 | | BP6 | | PP2,BS1,BP4,BP6 | rs5743278 | Likely Benign (I) | |
| 16 | 50746194 | 50746194 | G | A | exonic | NOD2 | . | Benign/Likely_benign | Sarcoidosis,_early-onset\|Crohn_disease\|Blau_syndrome | 0.00177 | 0.0016 | 0.000199681 | 0.0016 | 0.001758 | T | B | 0.074 | nonsynonymous SNV | NOD2:NM_001293557:exon3:c.G2291A:p.R764Q,NOD2:NM_022162:exon4:c.G2372A:p.R791Q | | | | | | | | | | | | | PP2 | | | | | | | | | | | | BP4 | | BP6 | | PP2,BP4,BP6 | rs104895464 | Likely Benign (II) | |
| 16 | 50746199 | 50746199 | G | A | exonic | NOD2 | . | Likely_benign | Sarcoidosis,_early-onset\|Crohn_disease | 0.001231 | 0.0011 | 0.000199681 | 0.0011 | 0.000586 | D | B | 22 | nonsynonymous SNV | NOD2:NM_001293557:exon3:c.G2296A:p.V766M,NOD2:NM_022162:exon4:c.G2377A:p.V793M | | | | | | | | | | | | | PP2 | PP3 | | | | | | BS3 | | | | | | | BP6 | | PP2,PP3,BS3,BP6 | rs104895444 | Likely Benign (I) | PMCID: PMC3378381; PMID: 12626759:BS3 Sixteen variants were indistinguishable from WT: R235C, T294S, R311W, H352R, N414S, S431L, A432V, E441K, R684W, R703C, A725G, V793M, R790Q, M836V, V955I, and V972I. |
| 16 | 50750810 | 50750810 | A | G | exonic | NOD2 | . | Conflicting_interpretations_of_pathogenicity,_risk_factor | Sarcoidosis,_early-onset\|not_specified\|Crohn_disease\|Blau_syndrome | 0.000769 | 0.0012 | 0.000399361 | 0.0012 | 0.001172 | D | D | 23.6 | nonsynonymous SNV | NOD2:NM_001293557:exon5:c.A2474G:p.N825S,NOD2:NM_022162:exon6:c.A2555G:p.N852S | | | | PS3 | | | | PM3 | | | | PP1 | PP2 | PP3 | | | | | | | | | | | | | | | PS3,PM3,PP1,PP2,PP3 | rs104895467 | Likely pathogenic (II) | PM3:Mutation spectrum of NOD2 reveals recessive inheritance as a main driver of<br>2 Early Onset Crohn's Disease :We identified two individuals who are compound heterozygous for the p. G908R risk<br>129 allele in trans with a less common NOD2 CD risk variant (p.N852S) in one case and a novel<br>130 truncating indel (p.S506Vfs*73) in the second case PMID: 21983784 :PS3: Western blot analysis showed that the point mutations did not affect expression compared with the wild-type protein (Fig. 4). As published earlier28,29, the fs1007insC mutant did not induce NF-κB activation after MDP stimulation. The MDP-induced NF-κB activation was also impaired in the presence of S431L and N852S (Fig. 4). |
| 16 | 50756540 | 50756540 | G | C | exonic | NOD2 | . | Conflicting_interpretations_of_pathogenicity,_risk_factor | INFLAMMATORY_BOWEL_DISEASE_1_(CROHN_DISEASE),_SUSCEPTIBILITY_TO\|not_specified\|Blau_syndrome\|Crohn_disease\|YAO_SYNDROME,_SUSCEPTIBILITY_TO | 0.010157 | 0.0099 | 0.00459265 | 0.0113 | 0.04885 | D | D | 31 | nonsynonymous SNV | NOD2:NM_001293557:exon7:c.G2641C:p.G881R,NOD2:NM_022162:exon8:c.G2722C:p.G908R | | | | | | PM1 | | | | | | | PP2 | PP3 | | | | BS1 | BS2 | | | | | | | | | | PM1,PP2,PP3,BS1,BS2 | rs2066845 | Benign (II) | |
| 16 | 50756571 | 50756571 | C | A | exonic | NOD2 | . | Likely_benign | Sarcoidosis,_early-onset\|Crohn_disease\|Blau_syndrome | 0.000539 | 0.0004 | 0.000599042 | 0.0006 | 0.002732 | D | D | 33 | nonsynonymous SNV | NOD2:NM_001293557:exon7:c.C2672A:p.A891D,NOD2:NM_022162:exon8:c.C2753A:p.A918D | | | | | | PM1 | | | | | | | PP2 | PP3 | | | | | | | | | | | | | BP6 | | PM1,PP2,PP3,BP6 | rs104895452 | VUS | |
| 16 | 50759450 | 50759450 | G | A | exonic | NOD2 | . | not_provided | Sarcoidosis,_early-onset | 0 | 3.31E-05 | 0 | 4.47E-05 | 0.009033 | T | D | 22.6 | nonsynonymous SNV | NOD2:NM_001293557:exon9:c.G2852A:p.G951E,NOD2:NM_022162:exon10:c.G2933A:p.G978E | | | | | | | PM2 | | | | | | PP2 | PP3 | | | | | | BS3 | | | | | | | | | PM2,PP2,PP3,BS3 | rs104895457 | VUS | PMID: 15044951:BS3:In contrast, multiple LRR mutants including A759V, G761S, L762M, V793M, I805N, P812T, D824N, A860G, L876I, Q889P, L891M, A892V, L904M, D913V, A920T, A944V, L947M, L949M, M956T, Q968H, G978E, S984T, K986I, A1002S, T1012I/S, G1032C and L1038F exhibited a wild-type phenotype. several of the rare CARD15/NOD2 variants found in CD patients, including A140T, R373C, V793M and G978E, exhibited wild-type function, suggesting that they likely represent neutral polymorphisms without clinical significance |
| 17 | 73826491 | 73826491 | G | A | exonic | UNC13D | . | Benign/Likely_benign | not_specified\|Familial_hemophagocytic_lymphohistiocytosis | 0.015866 | 0.0299 | 0.0103834 | 0.0192 | 0.02709 | T | B | 15.21 | nonsynonymous SNV | UNC13D:NM_199242:exon29:c.C2782T:p.R928C | | | | | | PM1 | | | | | | | | | | | | BS1 | BS2 | | | | | | BP4 | | BP6 | | PM1,BS1,BS2,BP4,BP6 | rs35037984 | Benign (II) | |
| 17 | 73831953 | 73831953 | G | A | exonic | UNC13D | . | Benign/Likely_benign | not_provided\|not_specified\|Familial_hemophagocytic_lymphohistiocytosis | 0.017915 | 0.0052 | 0.0191693 | 0.0038 | 0.002022 | D | P | 24 | nonsynonymous SNV | UNC13D:NM_199242:exon18:c.C1579T:p.R527W | | | | | | | | | | | | | | PP3 | | | | BS1 | | | | | | | | | BP6 | | PP3,BS1,BP6 | rs75366116 | Likely Benign (I) | |
| 17 | 73836317 | 73836317 | T | C | exonic | UNC13D | . | Benign | not_specified | 0.009073 | 0.0025 | 0.00938498 | 0.0019 | 0.000586 | D | B | 24.4 | nonsynonymous SNV | UNC13D:NM_199242:exon10:c.A847G:p.I283V | | | | | | | | | | | | | | PP3 | | | | | | | | | | | | | BP6 | | PP3,BP6 | rs61754871 | VUS | |
| 17 | 73837042 | 73837042 | T | C | exonic | UNC13D | . | | . | 0.001 | 0.0006 | 0.000399361 | 0.0007 | 0.001594 | D | B | 22.6 | nonsynonymous SNV | UNC13D:NM_199242:exon7:c.A610G:p.M204V | | | | | | PM1 | | | | | | | | PP3 | | | | | | | | | | | | | | | PM1,PP3 | rs144722609 | VUS | |
| 17 | 73839326 | 73839326 | C | T | exonic | UNC13D | . | Benign | not_specified | 0.016146 | 0.0159 | 0.0155751 | 0.0158 | 0.1881 | D | B | 21.6 | nonsynonymous SNV | UNC13D:NM_199242:exon3:c.G175A:p.A59T | | | | | | | | | | | | | | PP3 | | | | BS1 | BS2 | | BS4 | | | | | | BP6 | | PP3,BS1,BS2,BS4,BP6 | rs9904366 | Benign (II) | BS4:https://onlinelibrary.wiley.com/doi/pdf/10.1111/j.1365-2141.2009.07729.x: See pedigree |
| 17 | 78155422 | 78155422 | G | A | exonic | CARD14 | . | Benign | not_provided | 0.0044 | 0.0026 | 0.00539137 | 0.0013 | 0.005682 | T | B | 4.483 | nonsynonymous SNV | CARD14:NM_001257970:exon2:c.G185A:p.R62Q,CARD14:NM_024110:exon2:c.G185A:p.R62Q | | | | | | PM1 | | | | | | | | | | | | | | | | | | | BP4 | | BP6 | | PM1,BP4,BP6 | rs115582620 | Likely Benign (II) | |

| | | | | | | | | | | | | | | | | | | | | | | | | | | | | | | | | | | |
|---|---|---|---|---|---|---|---|---|---|---|---|---|---|---|---|---|---|---|---|---|---|---|---|---|---|---|---|---|---|---|---|---|---|---|
| 17 | 78157888 | 78157888 | G | C | exonic | CARD14 | . | Benign | . | 0.000231 | 0.005 | 0.00459265 | 0.0029 | 0.001175 | D | P | 26.7 | nonsynonymous SNV | CARD14:NM_001257970:exon4:c.G526C:p.D176H,CARD14:NM_024110:exon4:c.G526C:p.D176H | | PS3 | | PP3 | | | | | | | BP6 | PS3,PP3,BP6 | rs144475004 | VUS | PMCID: PMC3376540: PS3: Several rare variants (p.Glu142Lys, p.Glu142Gly, and p.Asp176His [rs144475004 (CARD14)]) in the coiled-coiled domain increased NF-kB activation two to five times more than did wild-type CARD14sh. Two variants clustered together: c.526G>C (p.Asp176His) (rs144475004 (CARD14)) and c.536G>A (p.Arg179His). Both amino acid substitutions were predicted to be damaging by PolyPhen2,27 and both lead to upregulation of NF-kB activation (2.78- and 1.38-fold increases, respectively).Compared with wild-type CARD14sh, a third variant, p.Asp176His (rs144475004 (CARD14)), leads to enhanced NF-kB activation. However, its frequency was similar in cases and controls, and it didn't increase NF-kB activity in vitro as much as other variants did. Hence, it might lie below the NF-kB-activation threshold required for disease. 26203641 PS3 n blotting of whole-cell extracts showed compQ(ARA)ble transfection efficiencies for the two constructs (Figure 2c), the analysis of soluble proteins demonstrated that the p.Asp176His change was associated with a significant decrease in the levels of free CARD14 (Figure 2d and e). This suggested that the mutant protein was forming insoluble oligomers. To validate this hypothesis, we used the anti-FLAG antibody to analyze the insoluble fraction of the cell extracts. As predicted, we found that CARD14 aggregates were significantly more abundant in the cells that had been transfected with the p.Asp176His cDNA (Figure 2f and g). Interestingly, a similar effect was noted for the constructs harboring the p.Leu156Pro and p.Glu138Ala disease alleles (Figure 2d, e, f, and g). Thus, our findings indicate that the pathogenic potential of the p. Asp176His change is compQ(ARA)ble with that of previously validated mutations. |
| 17 | 78157951 | 78157951 | G | A | exonic | CARD14 | . | Uncertain_significance | not_specified | 0.000309 | 0.0008 | 0.000599042 | 0.0005 | 0.000591 | D | B | 27.3 | nonsynonymous SNV | CARD14:NM_001257970:exon4:c.G589A:p.E197K,CARD14:NM_024110:exon4:c.G589A:p.E197K | | PS3 | | PP3 | | | | | | | | PS3,PP3 | rs200790561 | VUS | PMID: 26358359:PS3:These variants were c.349G>A (p.Gly117Ser), c.205C>T (p.Arg69Trp) and c.589G>A (p.Glu197Lys). c.589G>A (p.Glu197Lys) led to upregulation of NF-κB activity in a similar manner to that of previously descriQ(BED) psoriasis-associated mutations. |
| 17 | 78178911 | 78178911 | C | T | exonic | CARD14 | . | | . | 0 | 0.0002 | 0 | 0.0003 | 0.003739 | D | B | 15.25 | nonsynonymous SNV | CARD14:NM_024110:exon18:c.C2476T:p.R826W | | | | PP3 | | | | | | | | PP3 | rs139467141 | VUS | |
| 17 | 78179408 | 78179408 | G | A | exonic | CARD14 | . | Benign | . | 0.001081 | 0.0122 | 0.0209665 | 0.0072 | 0.001832 | T | B | 9.374 | nonsynonymous SNV | CARD14:NM_024110:exon19:c.G2648A:p.R883H | | | PM1 | | | BS1 | | | | BP4 | BP6 | PM1,BS1,BP4,BP6 | rs2289541 | Likely Benign (I) | |
| 18 | 2925284 | 2925284 | G | A | exonic | LPIN2 | . | Conflicting_interpretations_of_pathogenicity | not_specified | 0.001615 | 0.003 | 0.00259585 | 0.0027 | 0.002181 | T | B | 0.4 | nonsynonymous SNV | LPIN2:NM_014646:exon14:c.C1876T:p.P626S | | | | | | | | | | BP4 | | BP4 | rs150806357 | VUS | |
| 18 | 2925359 | 2925359 | C | T | exonic | LPIN2 | . | Benign/Likely_benign | not_specified | 0.008919 | 0.0079 | 0.00519169 | 0.0089 | 0.04826 | T | B | 22 | nonsynonymous SNV | LPIN2:NM_014646:exon14:c.G1801A:p.E601K | | | | | | | BS2 | | | BP4 | BP6 | BS2,BP4,BP6 | rs61735393 | Likely Benign (I) | |
| 18 | 2929103 | 2929103 | G | A | exonic | LPIN2 | . | Conflicting_interpretations_of_pathogenicity | Majeed_syndrome\|not_specified | 0.003385 | 0.0026 | 0.00199681 | 0.0026 | 0.002183 | D | P | 28.4 | nonsynonymous SNV | LPIN2:NM_014646:exon10:c.C1510T:p.L504F | | | PM1 | PP3 | | | | BS3 | | | | PM1,PP3,BS3 | rs104895500 | VUS | Molecular ChQ(ARA)cterization, Activity Assay Of Lipin2 Protein And Its Role In Psoriasis NCBI: BS3 :DNA analysis, SDS-PAGE and Western Blot analyses indicate that the Wild type and the six mutants are successfully sub-cloned and expressed in the expression hosts. Large scale expressions for all clones are carried out. The pure proteins were studied for their protein stability by CD spectroscopy and showed that there are no significant differences in stability. Read More: htGME(TP)://www.qscience.com/doi/abs/10.5339/qfarf.2013.BIOP-0142 he DNA analysis indicates that each fragment has been correctly cloned into the pYES2 vector. ChQ(ARA)cterization of the LPIN2 Gene and its Protein and Examination of its Role in Psoriasis BS3 The DNA analysis indicates that each fragment has been correctly cloned into the pYES2 vector.The analyses using SDS Gel Electrophoresis and Western Blot indicate that the Wild type and p.K387E are successfully expressed in S. cerevisiae while p.S734L is expressed in S. cerevisiae but at a lower level. Expression experiments are being done on the 4 remaining mutant proteins.Read More: htGME(TP)://www.qscience.com/doi/abs/10.5339/qfarf.2011.bmp63 |
| 18 | 2937699 | 2937699 | T | C | exonic | LPIN2 | . | Benign/Likely_benign | Majeed_syndrome\|not_specified | 0.000615 | 0.0043 | 0.00678914 | 0.0039 | 0.007439 | T | B | 22.8 | nonsynonymous SNV | LPIN2:NM_014646:exon7:c.A1159G:p.K387E | | | | | | | | BS3 | | BP4 | BP6 | BS3,BP4,BP6 | rs104895501 | Likely Benign (I) | Molecular ChQ(ARA)cterization, Activity Assay Of Lipin2 Protein And Its Role In Psoriasis NCBI: BS3 :DNA analysis, SDS-PAGE and Western Blot analyses indicate that the Wild type and the six mutants are successfully sub-cloned and expressed in the expression hosts. Large scale expressions for all clones are carried out. The pure proteins were studied for their protein stability by CD spectroscopy and showed that there are no significant differences in stability. Read More: htGME(TP)://www.qscience.com/doi/abs/10.5339/qfarf.2013.BIOP-0142 he DNA analysis indicates that each fragment has been correctly cloned into the pYES2 vector. ChQ(ARA)cterization of the LPIN2 Gene and its Protein and Examination of its Role in Psoriasis BS3 The DNA analysis indicates that each fragment has been correctly cloned into the pYES2 vector.The analyses using SDS Gel Electrophoresis and Western Blot indicate that the Wild type and p.K387E are successfully expressed in S. cerevisiae while p.S734L is expressed in S. cerevisiae but at a lower level. Expression experiments are being done on the 4 remaining mutant proteins.Read More: htGME(TP)://www.qscience.com/doi/abs/10.5339/qfarf.2011.bmp63 |
| 18 | 2937815 | 2937815 | G | A | exonic | LPIN2 | . | Benign/Likely_benign | Majeed_syndrome\|not_specified | 0.000461 | 0.0015 | 0.00119808 | 0.0013 | 0.002726 | D | B | 10.02 | nonsynonymous SNV | LPIN2:NM_014646:exon7:c.C1043T:p.P348L | | PS3 | | | | | | | | BP4 | BP6 | PS3,BP4,BP6 | rs34676691 | Likely Benign (II) | Molecular ChQ(ARA)cterization, Activity Assay Of Lipin2 Protein And Its Role In Psoriasis:PS3:The activity assay showed that one of the mutants (p.P348L) has higher phosphatase activity which a proximately two times more than the wild type. Read More: htGME(TP)://www.qscience.com/doi/abs/10.5339/qfarf.2013.BIOP-0142 |
| 18 | 2940693 | 2940693 | G | A | exonic | LPIN2 | . | Likely_benign | not_specified\|Majeed_syndrome | 0.000077 | 0.0043 | 0.00838658 | 0.0038 | 0.001172 | T | B | 22.3 | nonsynonymous SNV | LPIN2:NM_014646:exon5:c.C608T:p.S203F | | | | | | | | | | BP4 | BP6 | BP4,BP6 | rs144555528 | Likely Benign (II) | |
| 19 | 7708058 | 7708058 | C | T | exonic | STXBP2 | . | Benign/Likely_benign | not_specified | 0.011841 | 0.0109 | 0.00459265 | 0.0109 | 0.03831 | D | D | 26.3 | nonsynonymous SNV | STXBP2:NM_001127396:exon13:c.C1025T:p.T342M,STXBP2:NM_001272034:exon13:c.C1067T:p.T356M,STXBP2:NM_006949:exon13:c.C1034T:p.T345M | | | PM1 | PP3 | | BS1 | BS2 | | | | BP6 | PM1,PP3,BS1,BS2,BP6 | rs117761837 | Benign (II) | |
| 19 | 7710134 | 7710134 | C | T | exonic | STXBP2 | . | Benign/Likely_benign | not_specified | 0.008484 | 0.0158 | 0.00579073 | 0.0083 | 0.02898 | T | B | 15.23 | nonsynonymous SNV | STXBP2:NM_001127396:exon15:c.C1289T:p.A430V,STXBP2:NM_001272034:exon15:c.C1331T:p.A444V,STXBP2:NM_006949:exon15:c.C1298T:p.A433V | | | PM1 | | | BS1 | BS2 | | | BP4 | BP6 | PM1,BS1,BS2,BP4,BP6 | rs141309384 | Benign (II) | |
| 19 | 7712364 | 7712364 | A | G | exonic | STXBP2 | . | Benign | not_specified | 0.008931 | 0.0043 | 0.0121805 | 0.0028 | 0.005889 | D | B | 15.96 | nonsynonymous SNV | STXBP2:NM_001127396:exon18:c.A1654G:p.R552G,STXBP2:NM_001272034:exon18:c.A1696G:p.R566G,STXBP2:NM_006949:exon18:c.A1663G:p.R555G | | | PM1 | PP3 | | BS1 | | | | | BP6 | PM1,PP3,BS1,BP6 | rs61736586 | Likely Benign (I) | |
| 19 | 54299165 | 54299165 | G | A | exonic | NLRP12 | . | Likely_benign | Familial_cold_autoinflammatory_syndrome | 0.006074 | 0.0017 | 0.00419329 | 0.0012 | 0.00199 | . | . | 36 | stopgain | NLRP12:NM_001277129:exon8:c.C2875T:p.R959X,NLRP12:NM_001277126:exon9:c.C3049T:p.R1017X,NLRP12:NM_144687:exon9:c.C3046T:p.R1016X | | | PM1 | | | | | | | | BP6 | PM1,BP6 | rs35064500 | VUS | cannot mark PVS1 as NLRP12 has 9 exons and it falls in 9 exons |
| 19 | 54313859 | 54313859 | G | A | exonic | NLRP12 | . | Pathogenic | . | 0.000077 | 0.0004 | 0 | 0.0004 | 0.001172 | D | P | 24.1 | nonsynonymous SNV | NLRP12:NM_001277126:exon3:c.C1054T:p.R352C,NLRP12:NM_001277129:exon3:c.C1054T:p.R352C,NLRP12:NM_144687:exon3:c.C1054T:p.R352C | | PS3 | PM1 | PP3 | PP5 | | | | | | | PS3,PM1,PP3,PP5 | rs199881207 | Likely pathogenic (II) | PMID: 21538323:PS3:A heterozygous NLRP12 missense mutation involving a CpG site (c.1054C>T; p.Arg352Cys) was identified in exon 3, which encodes the nucleotide-binding site (NBS) of the protein, in 2 patients from different countries and carrying different NLRP12 hGME(AP)lotypes. The mutation, which does not alter the inhibitory effect of NLRP12 on NF-κB activation, increases speck formation and activates caspase 1 signaling. To define this new class of PFS, we propose the term NLRP12-associated disorders (NLRP12AD).CONCLUSION:Given the rarity of known NLRP12-associated disorders, the identification of this NLRP12 molecular defect contributes to the deliGME(NEA)tion of the clinical spectrum associated with mutations in this gene and highlights the importance of screening NLRP12 in patients presenting with unexplained PFS. This study also demonstrates, by means of functional assays, the deleterious effect of this recurrent missense mutation; the gain of function for speck formation and caspase 1 signaling associated with this NBS mutation is consistent with the inflammatory phenotype of PFS. |
| 19 | 54314003 | 54314003 | G | A | exonic | NLRP12 | . | Conflicting_interpretations_of_pathogenicity | not_specified\|Familial_cold_autoinflammatory_syndrome | 0.002691 | 0.0042 | 0.00219649 | 0.0044 | 0.007812 | D | D | 24 | nonsynonymous SNV | NLRP12:NM_001277126:exon3:c.C910T:p.H304Y,NLRP12:NM_001277129:exon3:c.C910T:p.H304Y,NLRP12:NM_144687:exon3:c.C910T:p.H304Y | | | PM1 | PP3 | | | | | | | | PM1,PP3 | rs141245482 | VUS | |
| 19 | 54314063 | 54314063 | G | A | exonic | NLRP12 | . | Pathogenic | Familial_cold_autoinflammatory_syndrome_2\|not_provided | 0.000769 | 0.0002 | 0.00239617 | 0.0002 | 0.005682 | . | . | 33 | stopgain | NLRP12:NM_001277126:exon3:c.C850T:p.R284X,NLRP12:NM_001277129:exon3:c.C850T:p.R284X,NLRP12:NM_144687:exon3:c.C850T:p.R284X | PVS1 | | PM1 | | PP5 | | | | | | | PVS1,PM1,PP5 | rs104895564 | Pathogenic 1(c) | PVS1: NLRP12 has 9 exons and it falls in 3 exons https://journals.plos.org/plosone/article?id=10.1371/journal.pone.0156981 |
| 19 | 55445884 | 55445884 | C | T | exonic | NLRP7 | . | | . | 0.002307 | 0.0007 | 0.00239617 | 0.0005 | 0.002657 | T | B | 0.157 | nonsynonymous SNV | NLRP7:NM_001127255:exon7:c.G2444A:p.R815H,NLRP7:NM_139176:exon7:c.G2360A:p.R787H,NLRP7:NM_206828:exon7:c.G2444A:p.R815H | | | | | | | | | | BP4 | | BP4 | rs150034626 | VUS | |
| 19 | 55447773 | 55447773 | G | A | exonic | NLRP7 | . | not_provided | Hydatidiform_mole | 0.000846 | 0.0012 | 0.000199681 | 0.0011 | 0.03731 | D | B | 15.89 | nonsynonymous SNV | NLRP7:NM_001127255:exon6:c.C2156T:p.A719V,NLRP7:NM_139176:exon6:c.C2072T:p.A691V,NLRP7:NM_206828:exon6:c.C2156T:p.A719V | | | | PP3 | | | BS2 | | | | | PP3,BS2 | rs104895526 | VUS | |
| 19 | 55450655 | 55450655 | T | C | exonic | NLRP7 | . | Benign | not_specified | 0.033831 | 0.0141 | 0.0247604 | 0.0131 | 0.02338 | T | B | 0.001 | nonsynonymous SNV | NLRP7:NM_001127255:exon4:c.A1532G:p.K511R,NLRP7:NM_139176:exon4:c.A1532G:p.K511R,NLRP7:NM_206828:exon4:c.A1532G:p.K511R | | | | | | BS1 | BS2 | | BS4 | BP4 | BP6 | BS1,BS2,BS4,BP4,BP6 | rs61743949 | Benign (II) | PMID:25082979:BS4: In contrast to the yeast system, the molecule to molecule interaction in HEK293T cells was clearly visible for the full-length wild-type NLRP7, the mutation R693P and the NSV K511R, while very faint for the two severe mutations, L398R and R693W. The less severe missense mutation R693P and the NSV K511R showed the same strong interaction with self-protein as the wild-type protein, |
| 19 | 55450707 | 55450707 | C | T | exonic | NLRP7 | . | | . | 0.000384 | 0.0002 | 0 | 4.88E-05 | 0.01328 | T | B | 0.033 | nonsynonymous SNV | NLRP7:NM_001127255:exon4:c.G1480A:p.A494T,NLRP7:NM_139176:exon4:c.G1480A:p.A494T,NLRP7:NM_206828:exon4:c.G1480A:p.A494T | | | | | | | BS2 | | | BP4 | | BS2,BP4 | rs61747407 | Likely Benign (I) | |
| 19 | 55450950 | 55450950 | G | A | exonic | NLRP7 | . | | . | 0.0016 | 0.0006 | 0.00179712 | 0.0004 | 0.001319 | D | D | 15.29 | nonsynonymous SNV | NLRP7:NM_001127255:exon4:c.C1237T:p.R413W,NLRP7:NM_139176:exon4:c.C1237T:p.R413W,NLRP7:NM_206828:exon4:c.C1237T:p.R413W | | | | PP3 | | | | | | | | PP3 | rs146427076 | VUS | |
| 19 | 55450991 | 55450991 | C | T | exonic | NLRP7 | . | not_provided | Hydatidiform_mole | 0 | 0.0005 | 0 | 0.0005 | 0.003063 | D | D | 22.3 | nonsynonymous SNV | NLRP7:NM_001127255:exon4:c.G1196A:p.C399Y,NLRP7:NM_139176:exon4:c.G1196A:p.C399Y,NLRP7:NM_206828:exon4:c.G1196A:p.C399Y | | | | PP3 | | | | | | | | PP3 | rs104895510 | VUS | |
| 19 | 55451018 | 55451018 | C | T | exonic | NLRP7 | . | | . | 0.001697 | 0.0005 | 0.00279553 | 0.0005 | 0.001447 | T | B | 6.418 | nonsynonymous SNV | NLRP7:NM_001127255:exon4:c.G1169A:p.R390H,NLRP7:NM_139176:exon4:c.G1169A:p.R390H,NLRP7:NM_206828:exon4:c.G1169A:p.R390H | | | | | | | | | | BP4 | | BP4 | rs141694326 | VUS | |
