## Supplementary Table 4 for "Genetic landscape of rare autoinflammatory disease variants in Qatar and Middle Eastern populations through the integration of whole-genome and exome datasets"

**Supplementary Table 4:** . Allele frequency of pathogenic and likely pathogenic variants compared with control global population (1000 genome, ExAC or gnomAD) using Fisher exact test (p<0.05). BED Bedouin, SAF Sub-Saharan African, EUR European, SOU South Asian, APY African Pygmy, ARA ARAB, PER Persian, NA not applicable]. Significant values are marked with *

| | | | | | Qatar Sub populations | | | | | | |
|---|---|---|---|---|---|---|---|---|---|---|---|
| **Pathogenic variant** | **Allele** | **Gene** | **Annotation** | **Qatar_All** | **Q(EUR)** | **Q(SOU)** | **Q(BED)** | **Q(AYG)** | **Q(ARA)** | **Q(PER)** | **Q(SAF)** |
| 2-224642579 | Ref-A | AP1S3 | | 0.994318 | NA | 1 | 1 | NA | 1 | 1 | 0.97059 |
| rs116107386 (AP1S3) | Alt-C | | | 0.005682 | NA | 0 | 0 | NA | 0 | 0 | 0.02941 |
| | AC/AN | | | 1/176 | 0/0 | 0/4 | 0/72 | 0/0 | 0/32 | 0/34 | 1/34 |
| | p-value | | Likely pathogen | 1 | 1 | 1 | 1 | 1 | 1 | 1 | 0.2362005 |
| 6-138196060 | Ref: C | TNFAIP3 | | 0.998406 | 1 | 0.99231 | 1 | 1 | 1 | 1 | 0.98413 |
| rs5029941 (TNFAIP3) | Alt: T | | | 0.001594 | 0 | 0.00769 | 0 | 0 | 0 | 0 | 0.01587 |
| | AC/AN | | | 3/1882 | 0/8 | 1/130 | 0/934 | 0/2 | 0/366 | 0/316 | 2/126 |
| | p-value | | Likely pathogen | 1 | 1 | 0.19362987 | 0.41441063 | 1 | 1 | 1 | 0.01885128* |
| 12-110034320 | Ref: G | MVK | | 0.996788 | 1 | 1 | 0.997845 | 1 | 0.997268 | 1 | 0.992063 |
| rs28934897 (MVK) | Alt: A | | | 0.003212 | 0 | 0 | 0.002155 | 0 | 0.008287 | 0 | 0.007937 |
| | AC/AN | | | 6/1868 | 0/8 | 0/130 | 2/928 | 0/2 | 3/362 | 0/312 | 1/126 |
| | p-value | | Pathogenic (IIIb | 0.07983319 | 1 | 1 | 0.66112252 | 1 | 0.02045803* | 1 | 0.18073726 |
| 15-55520906 | Ref: G | RAB27A | | 0.994155 | 1 | 1 | 0.998929 | 1 | 0.97268 | 1 | 1 |
| rs753966933 (RAB27A | Alt: A | | | 0.005845 | 0 | 0 | 0.001071 | 0 | 0.02732 | 0 | 0 |
| | AC/AN | | | 11/1882 | 0/8 | 0/130 | 1/934 | 0/2 | 10/366 | 0/316 | 0/126 |
| | p-value | | Likely pathogen | .024203E-21* | 1.00E+00 | 1.00E+00 | 0.01840619* | 1.00E+00 | .781588E-26* | 1.00E+00 | 1.00E+00 |
| 16-50750810 | Ref: A | NOD2 | | 0.998828 | 1 | 1 | 0.99768 | 1 | 1 | 1 | 1 |
| rs104895467 (NOD2) | Alt: G | | | 0.001172 | 0 | 0 | 0.00232 | 0 | 0 | 0 | 0 |
| | AC/AN | | | 2/1706 | 0/8 | 0/126 | 2/862 | 0/2 | 0/334 | 0/282 | 0/92 |
| | p-value | | Likely pathogen | 0.710876969 | 1 | 1 | 0.243883081 | 1 | 1 | 1 | 1 |
| 19-54314063 | Ref: G | NLRP12 | | 0.994318 | NA | 1 | 1 | NA | 1 | 1 | 0.97059 |
| rs104895564 (NLRP12 | Alt: A | | | 0.005682 | NA | 0 | 0 | NA | 0 | 0 | 0.02941 |
| | AC/AN | | | 1/176 | 0/0 | 0/4 | 0/72 | 0/0 | 0/32 | 0/34 | 1/34 |
| | p-value | | Pathogenic (Ib) | 0.02643622* | 1 | 1 | 1 | 1 | 1 | 1 | 0.00516364* |
| 19-54313859 | Ref: G | NLRP12 | | 0.998828 | 0.875 | 0.992063 | 1 | 1 | 1 | 1 | 1 |
| rs199881207 (NLRP12 | Alt: A | | | 0.001172 | 0.125 | 0.007937 | 0 | 0 | 0 | 0 | 0 |
| | AC/AN | | | 2/1706 | 1/8 | 1/126 | 0/862 | 0/2 | 0/334 | 0/282 | 0/92 |
| | p-value | | Likely pathogen | 0.129454087 | 0.002911628* | 0.04487728* | 1 | 1 | 1 | 1 | 1 |

| | | | GME Sub populations | | | | | | | | 1000 Genome | | | | | |
|---|---|---|---|---|---|---|---|---|---|---|---|---|---|---|---|---|
| **Pathogenic variant** | **Allele** | **GME_All** | **GME(AP)** | **GME(CA)** | **GME(ISR)** | **GME(NEA)** | **GME(NWA)** | **GME(SD)** | **GME(TP)** | **1000 Genome _all** | **AFR** | **AMR** | **EAS** | **EUR** | **SAS** | **Esp6500** |
| 2-224642579 | Ref-A | 0.99497 | 0.99415205 | 1 | 1 | 0.99592391 | 0.989899 | 1 | 0.99085366 | 0.997 | 0.998 | 0.997 | 1 | 0.99 | 0.999 | 0.990413 |
| rs116107386 (AP1S3) | Alt-C | 0.00503 | 0.00584795 | 0 | 0 | 0.00407609 | 0.010101 | 0 | 0.00914634 | 0.003 | 0.002 | 0.003 | 0 | 0.01 | 0.001 | 0.009587 |
| | AC/AN | 10/1986 | 2/342 | 0/264 | 0/20 | 3/736 | 2/198 | 0/116 | 3/328 | 15/5008 | 2/1322 | 2/694 | 0/1008 | 10/1006 | 1/978 | 115/11881 |
| | p-value | 0.1997308 | 1 | 0.2806828 | 1 | 0.3966478 | 0.6718835 | 1 | 0.747568 | | | | | | | |
| 6-138196060 | Ref: C | 0.9995 | 1 | 1 | 1 | 1 | 1 | 1 | 0.997 | 0.994 | 0.98 | 0.996 | 1 | 1 | 1 | 0.994464 |
| rs5029941 (TNFAIP3) | Alt: T | 0.0005 | 0 | 0 | 0 | 0 | 0 | 0 | 0.003 | 0.006 | 0.02 | 0.004 | 0 | 0 | 0 | 0.005536 |
| | AC/AN | 1/1986 | 0/342 | 0/264 | 0/20 | 0/736 | 0/198 | 0/116 | 1/328 | 30/5008 | 27/1322 | 3/694 | 0/1008 | 0/1006 | 0/978 | 72/12934 |
| | p-value | 0.39205057 | 1 | 1 | 1 | 0.63870017 | 1 | 1 | 0.4188855 | | | | | | | |
| 12-110034320 | Ref: G | 0.99647 | 0.9912 | 1 | 1 | 0.99728 | 1 | 1 | 0.9939 | NA | NA | NA | NA | NA | NA | 0.998308 |
| rs28934897 (MVK) | Alt: A | 0.003524 | 0.0087 | 0 | 0 | 0.00271 | 0 | 0 | 0.006 | NA | NA | NA | NA | NA | NA | 0.001692 |
| | AC/AN | 7/1986 | 3/342 | 0/264 | 0/20 | 2/736 | 0/198 | 0/116 | 2/328 | NA | NA | NA | NA | NA | NA | 22/12984 |
| | p-value | 0.04153817* | 0.01764697* | 1 | 1 | 0.32422029 | 1 | 1 | 0.09588056 | | | | | | | |
| 15-55520906 | Ref: G | NA | NA | NA | NA | NA | NA | NA | NA | NA | NA | NA | NA | NA | NA | NA |
| rs753966933 (RAB27A | Alt: A | NA | NA | NA | NA | NA | NA | NA | NA | NA | NA | NA | NA | NA | NA | NA |
| | AC/AN | NA | NA | NA | NA | NA | NA | NA | NA | NA | NA | NA | NA | NA | NA | NA |
| | p-value | NA | NA | NA | NA | NA | NA | NA | NA | | | | | | | |
| 16-50750810 | Ref: A | 0.9969 | 1 | 1 | 1 | 0.9945 | 1 | 0.9913 | 0.9969 | 0.9996 | 1 | 0.999 | 1 | 0.999 | 1 | 0.999231 |
| rs104895467 (NOD2) | Alt: G | 0.00302 | 0 | 0 | 0 | 0.0054 | 0 | 0.0086 | 0.003 | 0.0004 | 0 | 0.001 | 0 | 0.001 | 0 | 0.000769 |
| | AC/AN | 6/1986 | 0/342 | 0/264 | 0/20 | 4/736 | 0/198 | 1/116 | 1/328 | 2/5008 | 0/1322 | 1/694 | 0/1008 | 1/1006 | 0/978 | 10/12986 |
| | p-value | 0.024181757* | 1 | 1 | 1 | 0.009395715* | 1 | 0.119404012 | 0.301912508 | | | | | | | |
| 19-54314063 | Ref: G | NA | NA | NA | NA | NA | NA | NA | NA | 0.9976 | 0.991 | 1 | 1 | 1 | 1 | 0.999923 |
| rs104895564 (NLRP12 | Alt: A | NA | NA | NA | NA | NA | NA | NA | NA | 0.0023 | 0.009 | 0 | 0 | 0 | 0 | 7.7E-05 |
| | AC/AN | NA | NA | NA | NA | NA | NA | NA | NA | 12/5008 | 12/1322 | 0/694 | 0/1008 | 0/1006 | 0/978 | 10/12996 |
| | p-value | NA | NA | NA | NA | NA | NA | NA | NA | | | | | | | |
| 19-54313859 | Ref: G | 0.9994 | 1 | 1 | 1 | 1 | 1 | 1 | 0.9969 | NA | NA | NA | NA | NA | NA | 0.999231 |
| rs199881207 (NLRP12 | Alt: A | 0.0005 | 0 | 0 | 0 | 0 | 0 | 0 | 0.003 | NA | NA | NA | NA | NA | NA | 0.000769 |
| | AC/AN | 1/1982 | 0/342 | 0/264 | 0/20 | 0/736 | 0/198 | 0/116 | 1/328 | NA | NA | NA | NA | NA | NA | 1/13005 |
| | p-value | 0.513197403 | 1 | 1 | 1 | 1 | 1 | 1 | 0.112621115 | | | | | | | |

| | | Global population: gnomAD V2 | | | | | | | | |
|---|---|---|---|---|---|---|---|---|---|---|
| **Pathogenic variant** | **Allele** | **gnomAD V2 all** | **Latino** | **EURopean (Fin** | **EURopean (No** | **OTH** | **SAS** | **ASJ** | **AFR** | **EAS** |
| 2-224642579 | Ref-A | 0.992048 | 0.994596 | 0.995639 | 0.98663 | 0.99348 | 0.998231 | 0.995344 | 0.997209 | 0.99994697 |
| rs116107386 (AP1S3) | Alt-C | 0.007952 | 0.005404 | 0.004361 | 0.01337 | 0.00652 | 0.001769 | 0.004656 | 0.002791 | 5.303E-05 |
| | AC/AN | 2207 / 279700 | 184 / 35128 | 109 / 24928 | 1691 / 128180 | 57 / 7110 | 54 / 30350 | 45 / 10302 | 66 / 24180 | 1 / 19522 |
| | p-value | | | | | | | | | |
| 6-138196060 | Ref: C | 0.998395 | 0.999448 | 1 | 0.99997632 | 0.9990724 | 0.99996751 | 1 | 0.98269 | 1 |
| rs5029941 (TNFAIP3) | Alt: T | 0.001605 | 0.000552 | 0 | 2.368E-05 | 0.0009276 | 3.249E-05 | 0 | 0.01731 | 0 |
| | AC/AN | 467 / 282866 | 21 / 35440 | 0 / 25122 | 3 / 129170 | 8 / 7228 | 1 / 30616 | 0 / 10370 | 434 / 24970 | 0 / 19950 |
| | p-value | | | | | | | | | |
| 12-110034320 | Ref: G | 0.998409 | 0.998663 | 0.99862 | 0.997597 | 0.998915 | 0.9990904 | 0.9992118 | 0.9995833 | 0.9998939 |
| rs28934897 (MVK) | Alt: A | 0.001591 | 0.001337 | 0.00138 | 0.002403 | 0.001085 | 0.0009096 | 0.0007882 | 0.0004167 | 0.0001061 |
| | AC/AN | 443 / 280790 | 50 / 35436 | 33 / 23962 | 303 / 128360 | 9 / 7198 | 28 / 30616 | 8 / 10360 | 10 / 24932 | 2 /19926 |
| | p-value | | | | | | | | | |
| 15-55520906 | Ref: G | 0.9999837176 | 0.9999701564 | 1 | 0.99999101721 | 0.999816917 | 1 | 1 | 1 | 0.9999419347 |
| rs753966933 (RAB27A | Alt: A | 1.62824E-05 | 2.98436E-05 | 0 | 8.98279E-06 | 0.000183083 | 0 | 0 | 0 | 5.80653E-05 |
| | AC/AN | 4 / 250912 | 1 / 34522 | 0 / 21616 | 1 / 113396 | 1 / 6114 | 0 / 30602 | 0 / 10064 | 0 / 16234 | 1 / 18364 |
| | p-value | | | | | | | | | |
| 16-50750810 | Ref: A | 0.998911 | 0.9991575 | 1 | 0.9992425 | 0.998763 | 0.99996751 | 0.98355 | 0.99995839 | 1 |
| rs104895467 (NOD2) | Alt: G | 0.001089 | 0.0008425 | 0 | 0.0007575 | 0.001237 | 3.249E-05 | 0.01645 | 4.161E-05 | 0 |
| | AC/AN | 309 / 282898 | 29 / 35440 | 0 / 25122 | 102 / 129200 | 9 / 7228 | 1 / 30616 | 167 / 10370 | 1 / 24968 | 0 / 19954 |
| | p-value | | | | | | | | | |
| 19-54314063 | Ref: G | 0.9998411 | 0.9998547 | 1 | 1 | 0.9995355 | 1 | 1 | 0.998542 | 0.99994699 |
| rs104895564 (NLRP12 | Alt: A | 0.0001589 | 0.0001453 | 0 | 0 | 0.0004645 | 0 | 0 | 0.001458 | 5.301E-05 |
| | AC/AN | 42 / 282406 | 5 / 35412 | 0 / 25080 | 0 / 128878 | 2 / 7216 | 0 / 30612 | 0 / 10352 | 34 / 24916 | 1 / 19940 |
| | p-value | | | | | | | | | |
| 19-54313859 | Ref: G | 0.9996137 | 0.9997095 | 0.99992234 | 0.9995417 | 0.9998453 | 0.998863 | 1 | 0.99995836 | 1 |
| rs199881207 (NLRP12 | Alt: A | 0.0003863 | 2.91E-04 | 7.766E-05 | 0.0004583 | 0.0001547 | 0.001137 | 0 | 4.164E-05 | 0 |
| | AC/AN | 102 / 282638 | 10 / 35438 | 2 / 25084 | 53 / 129030 | 2 / 7224 | 34 / 30614 | 0 / 10358 | 1 / 24940 | 0 / 19950 |
| | p-value | | | | | | | | | |

| | | Global population | gnomAD V3 | | | | | | | | | |
|---|---|---|---|---|---|---|---|---|---|---|---|---|
| **Pathogenic variant** | **Allele** | **gnomAD V3 all** | **Amish** | **Latino** | **EURopean (Fin** | **EURopean (No** | **OTH** | **SAS** | **ASJ** | **AFR** | **ME** | **EAS** |
| 2-224642579 | Ref-A | 0.991353 | 0.97917 | 0.993067 | 0.993412 | 0.98596 | 0.98708 | 0.998758 | 0.995965 | 0.997152 | 0.996835 | 1 |
| rs116107386 (AP1S3) | Alt-C | 0.008647 | 0.02083 | 0.006933 | 0.006588 | 0.01404 | 0.01292 | 0.001242 | 0.004035 | 0.002848 | 0.003165 | 0 |
| | AC/AN | 1316 / 152188 | 19 / 912 | 106 / 15290 | 70 / 10626 | 955 / 68032 | 27 / 2090 | 6 / 4832 | 14 / 3470 | 118 / 41434 | 1 / 316 | 0 / 5186 |
| | p-value | | | | | | | | | | | |
| 6-138196060 | Ref: C | 0.994718 | 1 | 0.997776 | 1 | 0.99997061 | 0.99761 | 1 | 1 | 0.98159 | 1 | 1 |
| rs5029941 (TNFAIP3) | Alt: T | 0.005282 | 0 | 0.002224 | 0 | 2.939E-05 | 0.00239 | 0 | 0 | 0.01841 | 0 | 0 |
| | AC/AN | 804 / 152212 | 0 / 912 | 34 / 15286 | 0 / 10614 | 2 / 68040 | 5 / 2092 | 0 / 4830 | 0 / 3472 | 763 / 41448 | 0 / 316 | 0 / 5212 |
| | p-value | | | | | | | | | | | |
| 12-110034320 | Ref: G | 0.998535 | 1 | 0.998757 | 0.998587 | 0.997531 | 1 | 0.9997928 | 0.9991354 | 0.9995899 | 1 | 1 |
| rs28934897 (MVK) | Alt: A | 0.001465 | 0 | 0.001243 | 0.001413 | 0.002469 | 0 | 0.0002072 | 0.0008646 | 0.0004101 | 0 | 0 |
| | AC/AN | 223 / 152196 | 0 / 910 | 19 / 15280 | 15 / 10616 | 168 / 68034 | 0 / 2092 | 1 / 4826 | 3 / 3470 | 17 / 41452 | 0 / 316 | 0 / 5200 |
| | p-value | | | | | | | | | | | |
| 15-55520906 | Ref: G | NA | NA | NA | NA | NA | NA | NA | NA | NA | NA | NA |
| rs753966933 (RAB27A | Alt: A | NA | NA | NA | NA | NA | NA | NA | NA | NA | NA | NA |
| | AC/AN | NA | NA | NA | NA | NA | NA | NA | NA | NA | NA | NA |
| | p-value | | | | | | | | | | | |
| 16-50750810 | Ref: A | 0.9992381 | 1 | 0.9991494 | 1 | 0.9992798 | 0.998566 | 0.9997931 | 0.98704 | 0.99990356 | 0.996835 | 1 |
| rs104895467 (NOD2) | Alt: G | 0.0007619 | 0 | 0.0008506 | 0 | 0.0007202 | 0.001434 | 0.0002069 | 0.01296 | 9.644E-05 | 0.003165 | 0 |
| | AC/AN | 116 / 152248 | 0 / 910 | 13 / 15284 | 0 / 10624 | 49 / 68040 | 3 / 2092 | 1 / 4834 | 45 / 3472 | 4 / 41478 | 1 / 316 | 0 / 5198 |
| | p-value | | | | | | | | | | | |
| 19-54314063 | Ref: G | 0.9995597 | 1 | 0.9998032 | 1 | 0.9999853 | 0.9990449 | 0.9997929 | 1 | 0.998552 | 1 | 1 |
| rs104895564 (NLRP12 | Alt: A | 0.0004403 | 0 | 0.0001968 | 0 | 1.47E-05 | 0.0009551 | 0.0002071 | 0 | 0.001448 | 0 | 0 |
| | AC/AN | 67 / 152152 | 0 / 912 | 3 / 15244 | 0 / 10612 | 1 / 68032 | 2 / 2094 | 1 / 4828 | 0 / 3472 | 60 / 41450 | 0 / 316 | 0 / 5192 |
| | p-value | | | | | | | | | | | |
| 19-54313859 | Ref: G | 0.9997698 | 1 | 0.99993443 | 0.9998117 | 0.999647 | 0.999522 | 0.998962 | 1 | 0.99997585 | 1 | 0.9998076 |
| rs199881207 (NLRP12 | Alt: A | 0.0002302 | 0 | 6.557E-05 | 0.0001883 | 0.000353 | 0.000478 | 0.001038 | 0 | 2.415E-05 | 0 | 0.0001924 |
| | AC/AN | 35 / 152074 | 0 / 910 | 1 / 15250 | 2 / 10620 | 24 / 67992 | 1 / 2092 | 5 / 4816 | 0 / 3468 | 1 / 41412 | 0 / 316 | 1 / 5198 |
| | p-value | | | | | | | | | | | |

**Significant p-values marked with * and filled Red boxes**
