## Supplementary Data 1 for "Genetic landscape of rare autoinflammatory disease variants in Qatar and Middle Eastern populations through the integration of whole-genome and exome datasets"

**Supplementary Data 1. A detailed description of the methodology adopted for Annotation of Pathogenic /Benign Criterion.**

**Determination of PVS1**

We determine this variant as nonsense and frameshift using ANNOVAR. We checked whether the variant falls were in the last exon or last 50 base pair of the penultimate exon of the transcript by making bed files of nonsense and frameshift variants and overlapped with the last exon and last 50 bp of the penultimate exon. At last, we checked whether Loss of Function (LOF) is the main cause of disease for that gene.

**Determination of PS1 and PM5**

For determining, PS1 and PM5 we looked at the amino acid changes in clinvar with our novel variant. If the pathogenic or likely pathogenic variant in clinvar had the same amino acid change as well position, they were marked as PS1. In case, there was a different amino acid but the same position was the same, it was marked as PM5.

**Determination of PS2 and PM6**

We manually checked the literature to check whether the variant is originating de-novo by looking at the pedigree of the family. If the causative variant was absent in the parents but present in the offspring then it was marked as PS2. In case, the pedigree or the information was not sufficiently confirming the identity of the parents but was de-novo then it was regarded as PM6.

**Determination of PS3 and BS3**

These attributes were annotated using manual literature screening to check for variants for which in vivo and in vitro functional studies have been performed. If in the studies, a variant was reported to be responsible for affecting the protein function, it was marked as PS3. In case, the functional studies reported a benign effect then they were marked as BS3.

**Determination of PS4**

We manually screened literature containing case-control studies to determine the Odds Ratio. If the OR was found to be greater than 5 and the Confidence interval (CI) was more than 1, then the variant was regarded as PS4.

**Determination of PP1 and BS4**

We manually performed literature searches to analyze if the variants co-segregate with the family members. If the variant was present in all affected individuals in a family, then it is marked as PP1. If the variant was not present in any of the members of a family or present in an unaffected member of the family, then due to lack of segregation it was marked as BS4.

**Determination of PP2 and BP1**

We calculated the total number of pathogenic missense and stop gain variants for each gene using Clinvar. if the percentage of missense variant was >80% and stop-gain variant was <20% then for the nonsynonymous variant was marked as PP2. Otherwise, if the stop-gain variant was more than 80% and the nonsynonymous variant was < 20%, then the nonsynonymous variant was marked as BP1.

**Determination of PP3 and BP4**

The variants were annotated using the ljb26_all database of the ANNOVAR annotation tool to determine the PP3 and BP4 parameters. The variants were scored using the SIFT and PolyPhen2, two of the popularly used in-silico callers to score the variants for their predicted pathogenicity. While SIFT cut-offs classify the variants as deleterious or tolerated, PolyPhen2 predictions classify them into 3 categories as probably damaging, Possibly Damaging or Benign. We also considered CADD scores which contain PHRED scaled score, if the score is 10 predicts, the 10% most deleterious variants, similarly a score of 20 predicts 1% the most deleterious variant. However, we rationally considered the CADD scores above 15 to be interpreted as deleterious in our analysis. The variants classified as deleterious/damaging by at least two of the three in-silico callers were marked as PP3 (pathogenic). Similarly, variants were marked as BP4 if the majority of the in-silico tools predicted them to be benign/tolerated.

**Determination of PP4**

We used the OMIM database which is a comprehensive database of human genes and genetic phenotype whose main focus on the relationship of phenotype and genotype in Mendelian disorder and 15,000 genes. It is used to determine whether the disease was associated with just a single gene etiology. The variant was consistent means inherited in the family. The total number of benign should be less than 50% in a gene. All the variants fulfilling these criteria were marked as PP4.

**Determination of PP5 and BP6**

Variants were marked as PP5 and BP6 using publicly available database Clinvar. Those variants which had non-conflicting pathogenic/likely pathogenic calls from reputable laboratories were marked as PP5. Similarly, variants having non-conflicting benign/likely benign calls were marked as BP6.

**Determination of PM1**

We took the protein domains and their corresponding coordinates from the PFAM in UCSC gene track present in the UCSC browser and intersected the variant coordinates upon them. If the mutation hotspot was within these domains, then the variant was marked as PM1.

**Determination of PM2, BA1, BS1, and BS2**

Population datasets like 1000 Genome Project (ALL.sites.2015_08), Exome Sequencing Project (esp6500siv2_all), Exome Aggregation Consortium (exac03) and gnomAD derived from ANNOVAR tool databases were used to annotate the variants as BA1, BS1, and PM2. All those variants which had MAF more than 5% in any of the four population datasets were marked as BA1 whilst variants having MAF between 1 and 5% were considered as strong evidence to be benign for Mendelian disorder were marked as BS1. In case the variant is absent from all of the control population datasets or is at extremely low frequency in autosomal recessive i.e. < 0.01%, it was classified as moderate evidence to be pathogenic (PM2). All variants that occurred in genes at greater than 1% frequency in Qatar population dataset, regardless of population allele frequency, were marked as BS2.

**Determination of PM3 and BP2**

We first determined the mode of inheritance using OMIM. Using literature screening, we checked whether there were two heterozygous variants and the disorder was autosomal recessive. If both the mutations were found to be in trans they were marked as PM3. Similarly, if these were in cis they were considered to be BP2.

**Determination of PM4 and BP3**

For annotating in-frame insertions or deletions by overlapping them with the repeated region in the human genome using repeat masker. If the in-frame insertions or deletions fall in the repeated they were marked as BP3, otherwise, they were marked as PM4

**Determination of BP7**

Synonymous variant not in splice site was marked as BP7.
